# TWINGEN – protocol for an observational clinical biobank recall and biomarker study to identify individuals with high risk of Alzheimer’s disease

**DOI:** 10.1101/2023.11.03.23298018

**Authors:** Eero Vuoksimaa, Toni T. Saari, Aino Aaltonen, Sari Aaltonen, Sanna-Kaisa Herukka, Paula Iso-Markku, Tarja Kokkola, Aija Kyttälä, Sari Kärkkäinen, Hilkka Liedes, Miina Ollikainen, Teemu Palviainen, Ilona Ruotsalainen, Auli Toivola, Mia Urjansson, Tommi Vasankari, Henri Vähä-Ypyä, Markus M. Forsberg, Mikko Hiltunen, Anu Jalanko, Reetta Kälviäinen, Teijo Kuopio, Jaakko Lähteenmäki, Pia Nyberg, Minna Männikkö, Raisa Serpi, Sanna Siltanen, FinnGen, Aarno Palotie, Jaakko Kaprio, Heiko Runz, Valtteri Julkunen

## Abstract

**Introduction:** A better understanding of the earliest stages of Alzheimer’s disease (AD) could expedite the development or administration of treatments. Large population biobanks hold the promise to identify individuals at an elevated risk of AD and related dementias based on health registry information. Here, we establish the protocol for an observational clinical recall and biomarker study called TWINGEN with the aim to identify individuals at high risk of AD by assessing cognition, health and AD-related biomarkers. Suitable candidates were identified and invited to participate in the new study among Finnish biobank donors according to TWINGEN study criteria.

**Methods and analysis:** A multi-center study (n=800) to obtain blood-based biomarkers, telephone-administered and web-based memory and cognitive parameters, questionnaire information on lifestyle, health and psychological factors, and accelerometer data for measures of physical activity, sedentary behavior and sleep. A sub-cohort are being asked to participate in an in-person neuropsychological assessment (n=200) and wear an Oura ring (n=50). All participants in the TWINGEN study have genome-wide genotyping data and up to 48 years of follow-up data from the population-based older Finnish Twin Cohort (FTC) study of the University of Helsinki. TWINGEN data will be transferred to Finnish Institute of Health and Welfare (THL) biobank and we aim to further to transfer it to the FinnGen study where it will be combined with health registry data for prediction of AD.

**Ethics and dissemination:** This recall study consists of FTC/THL/FinnGen participants whose data were acquired in accordance with the Finnish Biobank Act. The recruitment protocols followed the biobank protocols approved by Finnish Medicines Agency. The TWINGEN study plan was approved by the Ethics Committee of Hospital District of Helsinki and Uusimaa (number 16831/2022). THL Biobank approved the research plan with the permission no: THLBB2022_83.

**Article summary:** *Strengths and limitations of this study:*

- A large sample of individuals is recruited from a representative biobank database
- Using health registry information, we exclude those with documented AD or other neurological or psychiatric diseases that can affect cognition. Pre-screening limits the sending of unnecessary invitations and saves costs
- Participants have up to 48 years of follow-up questionnaire and clinical data from the Finnish Twin Cohort study and these data can be combined with multifaceted Finnish health registry information. Previous genotype data is available in the biobank from all TWINGEN study participants.
- We assess the feasibility of remote cognitive testing and blood samples in large-scale screening of AD risk, translating to the requirements of intervention trials and clinical practice
- Limitations of the study are a lack of gold standard biomarkers (cerebrospinal fluid, positron emission tomography imaging) and neurological examinations

## Introduction

Alzheimer’s disease (AD) – the most common cause of dementia – is characterized by pathological accumulation of beta-amyloid (Aβ) and tau in the brain.^1^ As populations age, the prevalence of dementia is projected to nearly double every two decades^1,2^ and AD and related dementias are becoming one of the most common causes of death in many countries (20% of deaths in Finland ranking it as the third most common cause of death).^3^ The AD disease process starts up to 20-30 years before the diagnosis, so intervention trials targeted at preclinical or prodromal stages of AD are of high priority, but time-consuming and costly with screen-failure rates of 78-88%.^4^

In clinical practice, AD is diagnosed mainly based on the clinical phenotype, episodic memory impairment being the cognitive hallmark, while in research there has been a shift from clinical diagnosis to biological classification independent of the cognitive status.^5^ Evidence of diagnostic properties of different blood-based biomarkers is rapidly accumulating,^6^ but population-based studies are still scarce. In one population-based study, 11% of older adults (median age 74 years) without dementia were found to have Aβ pathology using blood-based biomarkers.^7^

Our earlier pilot study explored the feasibility of FinnGen, a nationwide Finnish biobank study^8^ in recruiting individuals with AD to an observational study via the biobank.^9^ Our protocol included blood-based biomarkers and remote cognitive assessment, approaches that are suggested to improve the recruitment of participants for AD trials.^10–12^ Another FinnGen study showed that biobank participants could be recontacted for additional data collection in a larger scale as well.^13^ The full potential of population biobank datasets lies in the large cohorts of undiagnosed individuals who are at the risk of developing AD in near future and might be suitable for targeted screening for early diagnostics and interventional trials encompassing both pharmacological treatments and lifestyle interventions.

After showing in our pilot study that recall of biobank participants with AD to a clinical study including multimodal (remote and in-person) cognitive assessment and blood-based biomarker analyses is feasible,^9^ we modified the protocol of the pilot study to target cognitively unimpaired older adults (based on health registry data) in the current study. Additionally, we augmented the assessment battery with passive technology for measuring physical activity, sedentary behavior, and sleep.

This protocol paper describes TWINGEN, a population-based follow-up study investigating the utility of easily implementable methods for assessing the risk of AD. We aim to conduct a proof-of-principle study for using biobank registries as a platform for recruiting participants suitable for clinical trials, particularly in diseases where recruitment and screening has generally been challenging. We also focus on the remote cognitive assessment methods and blood-based biomarkers of AD. The study also aims to enrich existing biobank data derived from a long-standing prospective twin study with cognitive and lifestyle measures. The research setting is unique as it utilizes the pre-screening and recall option based on the data of the biobank in combination with long preceding population-based follow-up data from the twin study.

## Methods and analysis

### Study participant selection

The target group of the TWINGEN study are individuals who have participated in the older Finnish Twin Cohort (FTC) study of the University of Helsinki (UH), and whose samples and data have been transferred to the Finnish Institute of Health and Welfare (THL) biobank in 2018. The main selection criteria in the biobank were previous participation in FTC, age (66-85), place of current residence in Finland, Finnish as the first language, and no known diagnosis affecting cognition in biobank records. Selection also included participation in the FinnGen biobank study.8 This criterion was important so that the TWINGEN data to be returned to THL Biobank can later be requested from the biobank for the FinnGen study and combined with its extensive gene and health register data. Below, we describe each of the data sources and the study protocol.

### The older FTC study

The older FTC study from the University of Helsinki is a population-based study that includes all Finnish same-sex twins born before 1958 and living in Finland at the start of the study in 1974 (Figure 1).^14^ The baseline survey was conducted in 1975 via postal questionnaire and 27750 individuals participated with an 89% participation rate. Follow-up questionnaires were sent in 1981 (n=24684 with a 84% response rate), 1990 for those born in 1930 or later (n=12502; 77%), and in 2011–2012 for those born in 1945–1957 (n=8410; 72%).^15^ Those born in 1938–1944 have also participated in MEMTWIN II study (n=1772) that utilized telephone interview to assess cognition.^16^ Some of the twins born in 1945–1957 have participated in Essential Hypertension EPIgenetics study (EH-EPI, n=445).^17^ The MEMTWIN II and EH-EPI study participants are the primary groups of interest in TWINGEN because they have either earlier cognitive (MEMTWIN II; Table 1) or multi-omics data (EH-EPI; Supplementary Table 1) available.

**Figure 1.**
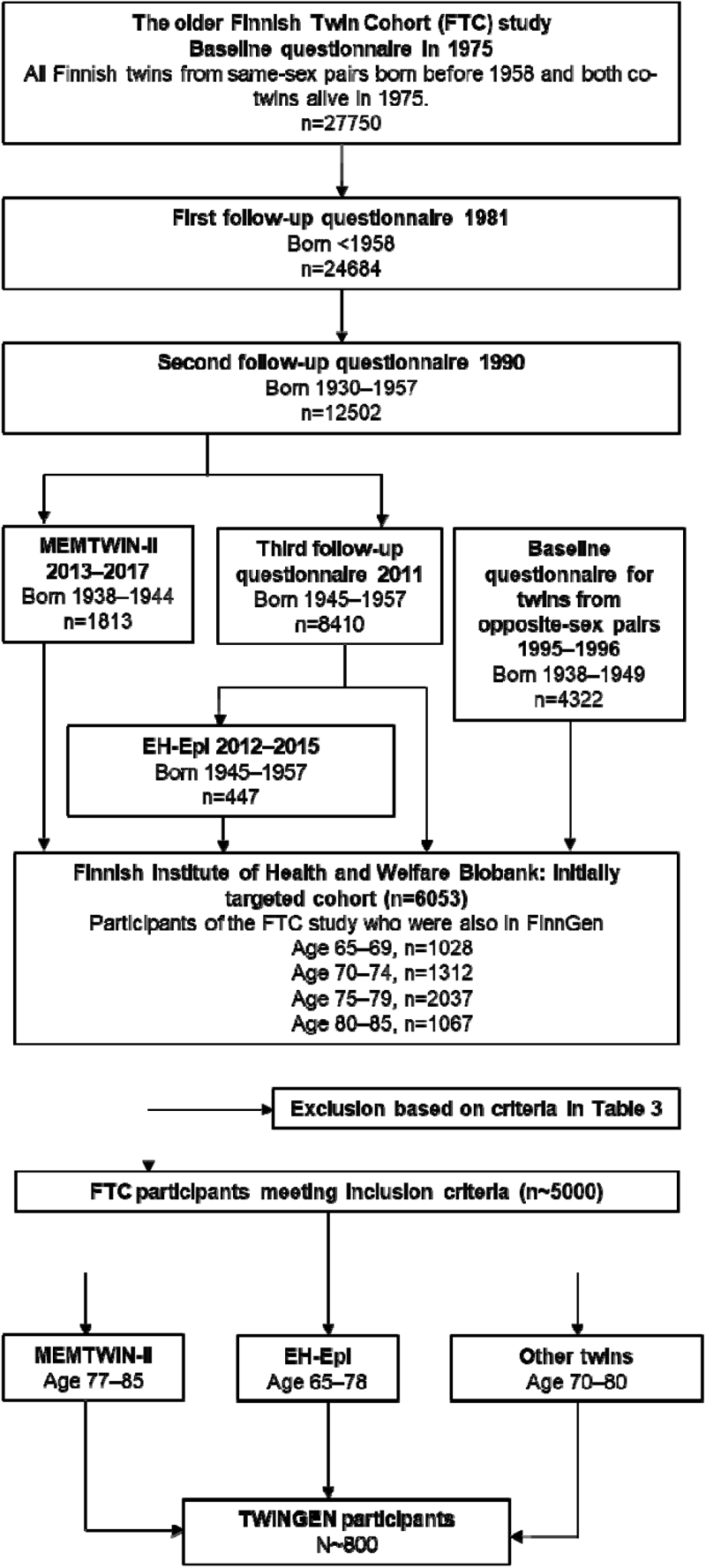
Flow-chart of study participant selection.

**Table 1.** Cognitive measures used in the TWINGEN study.

| In-person neuropsychological assessment | Telephone (TELE/TICS-m3) | Web-based computerized assessment (cCOG) |
| --- | --- | --- |
| **Memory and learning** |  |  |
| CERAD Word List Learning | Word List Learning | Episodic Memory Learning |
| CERAD Word List Recall | Word List Recall | Episodic Memory Recall |
| CERAD Word List Recognition |  | Episodic Memory Recognition |
| CERAD Constructional Praxis Recall |  |  |
| WMS-III Logical Memory Story A |  |  |
| WMS-III Logical Memory Story A Recall |  |  |
| **Executive function** |  |  |
| Trail Making Test-B |  | Modified Trail Making Test-B |
| Stroop Interference |  |  |
| Stroop Set-Shifting |  |  |
| **Visuospatial skills & visuoconstruction** |  |  |
| CERAD Constructional Praxis |  | Fragmented Letters |
| CERAD Clock Drawing Test |  |  |
| **Language skills & fluency** |  |  |
| CERAD Naming Test | Similarities |  |
| CERAD Semantic Fluency | Semantic Fluency |  |
| **Processing speed** |  |  |
| Trail Making Test-A |  | Modified Trail Making Test-A |
| Stroop Word Reading |  |  |
| Stroop Color Naming |  |  |
| **Global cognition** |  |  |
| CERAD MMSE | TELE Global Score | cCOG Global Score |
|  | TICS Global Score |  |
|  | TICS-m3 Global Score |  |

MEMTWIN-II participants have prior TELE/TICS-m data. CERAD-nb = Consortium to Establish a Registry for Alzheimer's Disease – neuropsychological battery; MMSE = Mini-Mental State Examination; TELE = telephone assessment for dementia; TICS = Telephone Interview for Cognitive Status; TICS-m = modified Telephone Interview for Cognitive Status; TICS-m3 = TICS-m including three learning trials in the Word List Learning; WMS-III = Wechsler Memory Scale 3rd edition

To achieve the target number of 800 participants, the biobank selection was expanded to the twins born in 1945–1952 who had not participated in MEMTWIN II or EH-EPI. Most of the invited twins were from same-sex pairs, but we also invited twins from opposite-sex pairs included in the older FTC study in year 1995–96, when they replied to a brief health questionnaire (Figure 1). We prioritized invitations to participants who lived closest to one of the six study sites. All TWINGEN participants from same-sex twin pairs have longitudinal questionnaire data on health and health-related behaviors from years 1975, 1981 and 1990, and those born in 1945–1957 have data also from year 2011–2012. An overview of longitudinal data available for the MEMTWIN II and EH-EPI and other TWINGEN participants are presented in Table 2. Necessary inclusion criteria for TWINGEN were available DNA sample in THL Biobank and not having any of the exclusion criteria (Table 3). Additional references for studies using the older FTC data are found in Supplementary Table 2.

**Table 2.**
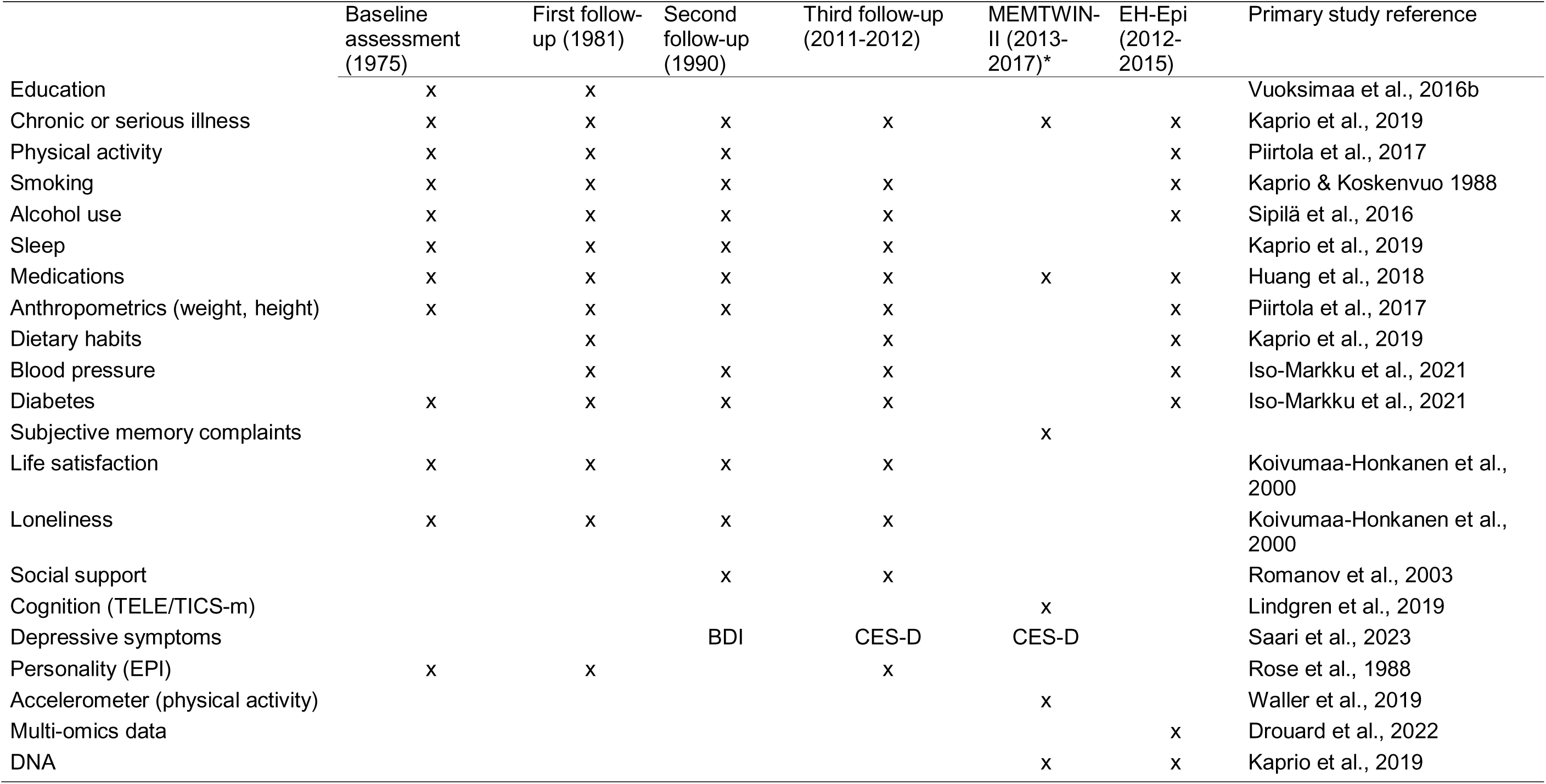
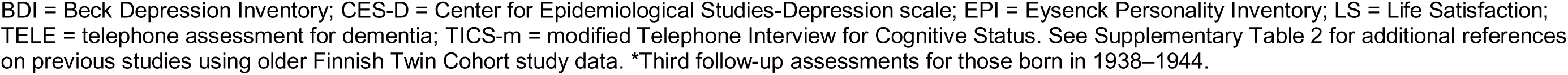
Previous data from the participants recruited in TWINGEN.

|  | Baseline<br>assessment<br>(1975) | First follow-up (1981) | Second<br>follow-up<br>(1990) | Third follow-up<br>(2011-2012) | MENTWIN-<br>II (2013-2017)* | EH-Epi<br>(2012-2015) | Primary study reference |
| --- | --- | --- | --- | --- | --- | --- | --- |
| Education | x | x |  |  |  |  | Vuoksima et al., 2016b |
| Chronic or serious illness | x | x | x | x | x | x | Kaprio et al., 2019 |
| Physical activity | x | x | x |  |  | x | Piirtola et al., 2017 |
| Smoking | x | x | x | x |  | x | Kaprio & Koskenvuo 1988 |
| Alcohol use | x | x | x | x |  | x | Sipilä et al., 2016 |
| Sleep | x | x | x | x |  |  | Kaprio et al., 2019 |
| Medications | x | x | x | x | x | x | Huang et al., 2018 |
| Anthropometrics (weight, height) | x | x | x | x |  | x | Piirtola et al., 2017 |
| Dietary habits |  | x |  | x |  | x | Kaprio et al., 2019 |
| Blood pressure |  | x | x | x |  | x | Iso-Markku et al., 2021 |
| Diabetes | x | x | x | x |  | x | Iso-Markku et al., 2021 |
| Subjective memory complaints |  |  |  |  | x |  |  |
| Life satisfaction | x | x | x | x |  |  | Koivumaa-Honkanen et al., 2000 |
| Loneliness | x | x | x | x |  |  | Koivumaa-Honkanen et al., 2000 |
| Social support |  |  | x | x |  |  | Romanov et al., 2003 |
| Cognition (TELE/TICS-m) |  |  |  |  | x |  | Lindgren et al., 2019 |
| Depressive symptoms |  |  | BDI | CES-D | CES-D |  | Saari et al., 2023 |
| Personality (EPI) | x | x |  | x |  |  | Rose et al., 1988 |
| Accelerometer (physical activity) |  |  |  |  | x |  | Waller et al., 2019 |
| Multi-omics data |  |  |  |  |  | x | Drouard et al., 2022 |
| DNA |  |  |  |  | x | x | Kaprio et al., 2019 |

BDI = Beck Depression Inventory; CES-D = Center for Epidemiological Studies-Depression scale; EPI = Eysenck Personality Inventory; LS = Life Satisfaction; TELE = telephone assessment for dementia; TICS-m = modified Telephone Interview for Cognitive Status. See Supplementary Table 2 for additional references on previous studies using older Finnish Twin Cohort study data. \*Third follow-up assessments for those born in 1938–1944.

**Table 3.** Exclusion criteria for the TWINGEN study.

| ICD-10 code | Explanation |
| --- | --- |
| G30 and F00 | Any variant of Alzheimer's disease or any dementia relating to Alzheimer's disease, dementia with Lewy bodies, frontotemporal dementia and mixed dementia |
| F01-F03 | Dementia, any etiology |
| G20 | Parkinson's disease |
| G35 | Multiple sclerosis |
| I60, I61 and I63 | Intracerebral hemorrhage, subarachnoid hemorrhage or ischemic stroke and their subcategories |
| S06.1-S06.7 | Traumatic brain injuries other than concussion (S06.0) |
| F20 | Schizophrenia |
| F31 | Bipolar disorder |
| F33, F32.1-F32.3, F34 | Recurrent depression, moderate or severe depression and long-lasting mood disorders |
| F60 | Personality disorders |
| F10 | All diagnoses relating to excessive usage of alcohol |
| F11-F19 | Intoxication because of opioids, cannabinoids, sedative medication, cocaine or hallucinogens |
| F70-73 | Different stages of intellectual disabilities |

### THL Biobank and the FinnGen study

DNA samples of FTC participants were transferred to the FinnGen project, a nationwide biobank research study including sample donors with DNA samples from Finnish biobanks. In TWINGEN, seven biobanks were included: THL Biobank, Arctic Biobank, Biobank Borealis, Biobank of Eastern Finland, Central Finland Biobank, Finnish Clinical Biobank Tampere and Helsinki Biobank. FinnGen has combined all biobank samples and linked these to registry data including e.g., medication history, hospital discharge, and mortality.^8^ The biobank sample donors included in FinnGen provide a good representation of the Finnish population (risteys.finregistry.fi).

### Recall procedure

The results of a THL Biobank feasibility assessment indicated that there were 6053 individuals aged 65–85 who have participated in the FTC of UH and have data in the THL Biobank. Approximately 1000 individuals were excluded due to our exclusion criteria of Alzheimer’s disease and other neurological or psychiatric diseases that can affect cognition (Figure 1; Table 3). Furthermore, the genotype data of approximately 100 individuals have been verified by FinnGen, but these data have not been returned to their respective biobanks at the time of the feasibility assessment; thus, these individuals are excluded from the pool of potentially eligible participants. The target sample size of 800 participants was chosen as it fits the timeframe and resources of the study and is sufficiently large for stratifying participants at varying risks for AD.^7^

As a biobank recall study, TWINGEN participants are contacted by the THL Biobank by an invitation letter. The invitation letter includes information about the individuals’ prior participation to the older FTC study and the transfer to and storage of samples and data at the THL Biobank, information about the participation in the FinnGen study via the biobank, and information about the new TWINGEN study. The invitation letter also contains two separate consent forms: one for the participation in the TWINGEN study and the biobank consent for THL Biobank. The consent for THL Biobank is needed to store the new samples and data obtained in TWINGEN to THL Biobank and to confirm the biobank participation with a written biobank consent, which is the primary basis for storing samples and data into a biobank. After receiving consent forms, UH research staff contacts the potential participants to verify that the individuals have understood the purpose and procedures of the study. Additionally, as the health registry data available in the biobank is not up to date but reflects status at the end of year 2022, research staff verifies (via telephone) that the exclusion criteria are not met. Eligibility of each individual is assessed independently of their co-twin’s vital status or eligibility.

### Data collection

Data collection started on March 29^th^ 2023 and is planned to be completed by the end of 2023. Data collection is conducted in six locations across Finland: at the Institute for Molecular Medicine Finland (FIMM), University of Helsinki in Helsinki, biobanks in four locations across Finland: in Jyväskylä (Central Finland biobank), Kuopio (Biobank of Eastern Finland), Oulu (Arctic Biobank and Biobank Borealis of Northern Finland) and Tampere (Finnish Clinical Biobank Tampere) and at the clinical laboratory of Turku University of Applied Sciences. Protocol for all participants includes telephone- and computer-administered cognitive testing, blood-draw and self-report questionnaire. Additionally, a waist-worn accelerometer will be given to participants who are willing to complete one week at-home measurement. Furthermore, individuals living in greater Helsinki area are invited to participate in in-person neuropsychological testing (target sample size n=200) and measurement of weight, height, waist circumference and blood pressure. In Helsinki, we also provide Oura rings for 50 participants who are willing and able to wear the ring and use the associated mobile app. After participating in the study, participants receive a report based on their performance in telephone interview-based and computerized tests of cognition and about their physical activity. A detailed description of the data collections is provided below.

### Telephone interview for assessing cognitive status and function

Two validated telephone-administered cognitive screening instruments are used: a telephone assessment for dementia (TELE)^18^ and the modified Telephone Interview for Cognitive Status (TICS-m).^19^ TELE and TICS have been translated to Finnish and adapted to Finnish culture^20^ and both have been used in the older FTC study. TICS-m, a modified version of TICS includes an additional delayed free recall of the 10-word list and has been used in the MEMTWIN II sub-study.^21^ We further modified the TICS-m by including three learning trials of the 10-word list and this instrument is later referred to as TICS-m3.^9^ We also included semantic fluency. Three trial word list task yields immediate and delayed free recall measures of episodic memory. Semantic fluency score is the number of animals named in 1 minute (Table 1). In the telephone interview, the participants are also asked about their functional abilities regarding household maintenance, ambulation, shopping, dressing and undressing, use of mobility aids, memory problems and possible visits to the doctor regarding memory problems. Telephone interviews were conducted by trained study nurses or psychologists.

### Computerized web-based cognitive testing

Web-based cCOG tool created by Combinostics is used for computerized cognitive testing.^22^ It includes six subtests: Episodic Memory (learning & recall), Reaction Time, Modified Trail Making A and B and Distorted Letters. The tasks measure visual processing, memory, processing speed, attention, and executive function (Table 1). In addition to cognitive tests, cCOG includes background questions (education) and a seven-item questionnaire designed to assess probable dementia with Lewy bodies.^23^ The test battery takes about 25 minutes to complete and is performed via a keyboard and a mouse or a touchscreen device.

### In-person neuropsychological tests

The participants in Helsinki study site undergo an in-person neuropsychological assessment, with a target sample size of 200 individuals. The minimum sample size to detect medium correlations (r = .3) between cognitive measures with a significance level = .05 and power = .8 is n = 85, thus, the target sample size of 200 individuals with in-person cognitive assessment is adequate for examining correlations between in-person and remote cognitive measures. The larger sample size is expected to be adequate for factor analyses of the neuropsychological battery.

The in-person neuropsychological assessment consists of the Consortium to Establish a Registry for Alzheimer’s Disease neuropsychological battery (CERAD-nb)^24^ and tests measuring executive functions, processing speed and episodic memory. The CERAD-nb includes Mini-Mental State Examination and Semantic Fluency, abbreviated Boston Naming Test, Word List Learning, Recall and Recognition, and Constructional Praxis (Copy and Recall). The Finnish version of the CERAD-nb also includes Clock Drawing test.^25^ Finnish education adjusted cut-offs are available for total score and for each sub-test.^26^ In addition to the CERAD-nb, we include the following tests: Logical Memory story A from the Wechsler Memory Scale 3rd edition,^27^ Trail Making Test A and B,^28^ and Stroop test.^29^ The version of Stroop used in this study is the 40-item version used in the FINGER study^30^ with an additional fourth “set-shifting” condition whereby the task is to name the color of ink (as in the classical Stroop condition) or to read out the color-word when the word is inside a rectangle. Neuropsychological tests are administered by trained psychologists. A summary of key cognitive measures of all three modalities are presented in Table 1.

### Blood sample

A venous blood sample is drawn from the inside of the elbow or alternatively from the back of the hand. A total of six tubes are collected: three BD Vacutainer K2EDTA (10/10ml) tubes, two BD Vacutainer SSTII Advance-serum gel tubes (10/8.5ml), and one BD PAXgene Blood RNA (7/2.5ml) tube (Supplementary Figure 1). Processing will be done immediately after the samples have been taken. Serum tubes are allowed to clot at least 30 minutes (max. 60’) before separating. EDTA-plasma tubes do not clot but two EDTA tubes are also let sit for the same time as serum because of the easier workflow. Serum and two EDTA-plasma tubes are centrifuged (1500G) for 10’ and the supernatants are pooled within serum and plasma. Samples are apportioned into 0.5 ml aliquots and stored at −20°C. One EDTA tube and PAXgene RNA-tube will be stored as a whole blood at −20°C. Serum, plasma and RNA samples final storage temperature is at −80°C. One of the 0.5 ml EDTA-plasma aliquot will be sent to University of Eastern Finland for biomarker analysis. RNA, EDTA whole blood samples, and half of the serum and plasma aliquots are dedicated to the Twin Study, while the other half will be available for research via THL Biobank.

### Blood-based AD biomarkers

The primary blood-based biomarkers include phosphorylated-tau181 (p-tau181), phosphorylated-tau217 (p-tau217), Aβ1-42/40, glial fibrillary acidic protein (GFAP) and neurofilament light chain (NfL); all measured using Simoa HD-X analyzer (Quanterix, Billerica, Massachusetts, USA). Plasma p-tau181 levels are quantified using Simoa p-tau181 Advantage V2.1 Kit(Ref# 104111, Quanterix),^31^ Aβ1-40, Aβ1-42, GFAP and NfL levels using Simoa Neurology 4-Plex E Advantage Kit (Ref# 103670, Quanterix),^32^ and p-tau217 levels using ALZpath Simoa pTau-217 v2 Assay Kit (Ref# 104371, Quanterix).^33^ Prior to analyses, EDTA plasma samples are thawed, mixed and centrifuged (10,000xg, 5 min, +20 °C). These biomarkers are determined at the Biomarker Laboratory of University of Eastern Finland.

### Apolipoprotein E (APOE) genotype and polygenic risk scores

APOE status (ε4-carrier vs. non-carrier and number of ε4-alleles) is defined by two single-nucleotide polymorphisms, rs429358 and rs7412, in chromosome 19^34^ and polygenic risk score (PRS; with and without APOE) of AD is based on the Bellenguez et al.^35^ or newer meta-analysis if available. Genetic data will be used to calculate also PRS’s for diseases and traits (such as cardiovascular disease and educational attainment) that are related to risk and protective factors of dementia.^36^

### Accelerometer-measured physical activity, sedentary behavior, and sleep

A tri-axial accelerometer (UKK RM42, UKK Terveyspalvelut Oy, Tampere, Finland) is used to monitor participants’ daily physical activity, sedentary behavior, and sleep for seven consecutive days (Table 4).^37^ In addition to total time spent in physical activity of different intensities and sedentary behavior, we will also measure number of bouts and length of the bouts of physical activity and sedentary behavior. The participants receive the devices during their in-person visit at FIMM or by mail if participating in other location. Participants are asked to wear the accelerometer on the hip during waking hours and on the wrist during sleep. At least a 4-day monitoring period with a minimum of 10 hours wear-time a day will be required for the adequate accelerometer data collection.^38^

**Table 4.** Core UKK RM42 accelerometer and Oura ring parameters.

|  |  |
| --- | --- |
| The UKK RM42 accelerometer | Oura ring |
| Target n = 800 | Target n = 50 |
| Participants from all sites | Participants from Helsinki site |
| Wearing time = 1 week | Wearing time = 2 weeks |
| **Physical activity parameters** |  |
| Light physical activity | Low intensity activity |
| Moderate physical activity | Medium intensity activity |
| Vigorous physical activity | High intensity activity |
| Total physical activity | Total physical activity |
| Number of steps | Number of steps |
| Standing time |  |
| **Sedentary behavior parameters** |  |
| Lying time | Inactive time |
| Reclining time | Resting time |
| Sitting time |  |
| Number of breaks during sedentary time |  |
| **Sleep parameters** |  |
| Total sleep time | Total sleep time |
| Restless sleep time | Restless sleep percentage |
| Restful sleep time | Total amount of deep sleep |
|  | Total amount of REM sleep |
|  | Total amount of light sleep |

The UKK RM42 device and its closely related counterpart Hookie AM 20 accelerometer have been used in samples with over 18,000 Finnish 18–85-year-old adults.^37,39^ Thus, the UKK RM42 accelerometer is usable in the TWINGEN sample of 63–85-year-olds, and we will get an opportunity to compare the measurements against normative data of Finnish adults. The analyses of raw acceleration data of the UKK RM42 are based on validated algorithms; the technical details related to the recording and analyzing of raw acceleration data are given elsewhere.^40–42^

### Oura-measured physical activity, sedentary behavior and sleep

Oura ring (Gen3 Heritage, Ōura Health Ltd., Oulu, Finland) will be given to 50 participants in Helsinki study collection site. First, participants use a ring-size kit to determine optimal ring size and then they receive the ring either at their in-person visit or by mail. Participants are asked to wear the ring (width: 7.9mm, thickness: 2.55 mm, weight: 4–6g) in any finger for two weeks during day and night, except when charging the ring every 4–6 days (20–80 minutes to fully charge). To monitor participant’s sleep, sedentary behavior and physical activity (Table 4), the Oura ring uses infrared photoplethysmography sensors, negative temperature coefficient sensor and 3D accelerometer. Participants receive written instructions on using the Oura ring and downloading the Oura mobile application, through which they can access their own data. If necessary, the research staff provides phone guidance for both using the ring and installing the application. The data from Oura ring is transferred to participant’s Oura application when opening the application and to a cloud server. Data collection will be monitored from Oura cloud server and participants’ will be sent a reminder if there are no data from previous 2 days.

The Oura ring’s sleep stage detection algorithm (wake, light non-rapid eye movement [NREM] sleep, deep NREM sleep, rapid eye movement [REM] sleep) has been validated against polysomnography and it showed 80–96% accuracy, 74–82% sensitivity and 79–98% specificity.^43^ Furthermore, moderate-to-vigorous intensity physical activity and step count of Oura ring has shown strong correlations with accelerometer-measured corresponding values.^44^

### Questionnaire

Participants are given a 16-page self-report questionnaire that includes many of the same measures as in previous questionnaires in years, 1975, 1981, 1990 and 2011 (Table 2). Questions cover anthropometrics, demographics, social relationships, chronotype, health (general, cardiovascular, dementia, memory, medications, vision, hearing, balance and mobility) and health-related behavior including sleep, physical activity, smoking and alcohol use. Psychological wellbeing scales included in the questionnaire are: 8-item Center for Epidemiologic Studies Depression;^45,46^ 7-item Purpose in Life subscale from Ryff’s Scales of Psychological Well-Being;^47,48^ Extraversion (9 items) and Neuroticism (10 items) from the short version of Eysenck Personality Inventory;^49,50^ and four-item life satisfaction scale derived from questionnaires by Allardt.^51,52^

### Patient and public involvement statement

None.

### Aims, data analysis and future directions

In addition to the overarching aim of assessing the feasibility of biobank recall in the context of preclinical AD, we also have more focused research questions. By combining all data, we aim to stratify our participants in sub-groups of low, intermediate and high AD risk based on genetic, biomarker, cognitive, lifestyle and symptom data. This information is potentially useful for improving participant selection for AD drug and intervention trials. These methods would also be valuable in clinical settings where non-invasive and widely available tools for evaluating the presence of AD pathology underlying cognitive symptoms is important, especially once disease-modifying treatments become available. TWINGEN will also establish a baseline cohort that can be used in follow-up studies with neuroimaging and cerebrospinal fluid biomarkers. Through FinnGen, we will be able to follow these individuals based on their health records and thus will be ultimately able to predict progression to AD.

We aim to assess the comparability of in-person, telephone-based and computerized cognitive assessments using correlation analysis. We will also explore the distributions and correlations among blood-based biomarkers of AD-related pathologies and investigate the associations between cognition and biomarkers. Cognition will be treated both categorically (cognitive status) and as continuous outcomes for domain-specific measures (e.g., episodic memory, executive function). For the domain-specific cognitive composites, factor scores will be calculated similar to previous studies on preclinical AD and mild cognitive impairment.^53,54^

Additionally, the effect of genetic and lifestyle (with up to 48 years of follow-up) factors can be used in predicting cognitive and biomarker status. The measures of physical activity, sedentary behavior and sleep (Table 4) allow us to explore the relationships between physical activity and sleep with cognition and biomarkers. Additional parameters of physical activity and sleep are also available for detailed analyses. Although inclusion in the study was not dependent on the co-twin’s participation, it is expected that full twin pairs will also participate. This will allow studying if the between-family associations are also evident in within-family comparisons and we can identify twin pairs who are discordant for cognition or biomarkers^55^

### Ethics and dissemination

According to the Finnish Biobank Act, research collections, which have been collected, or whose collection started before the Biobank Act became into force on 1 September 2013, can be transferred to a biobank by a specific procedure, which includes an ethical evaluation and informing the sample donors either personally or by public announcement. Accordingly, biological samples of the FTC, including DNA and associated data were transferred to THL biobank in December 2018 to facilitate biobank research. The action of transferring FTC data to THL Biobank was publicly announced in major newspapers.

Recruitment protocols followed the biobank protocols approved by Fimea. The FinnGen study was approved by the Coordinating Ethics Committee of Hospital District of Helsinki and Uusimaa (HUS; statement number HUS/990/2017). The permit numbers of the decisions made by Finnish Institute for Health and Welfare and the Biobank Access Decisions for FinnGen samples and data utilized in FinnGen Data Freeze 9 are presented in Acknowledgements.

Before entering the TWINGEN study all potential study participants received detailed information regarding this study by a formal information letter. All study participants were asked for and provided written informed consent. The recall study was reviewed and approved by the ethics committee of HUS (approval number 16831/2022). THL Biobank approved the research plan with the permission no: THLBB2022_83.

The data acquired in this study are managed and initially stored by UH. Keeping with the Biobank Act and the consent given by the participants, the data are also transferred to the THL Biobank for later research use. FinnGen request to use the TWINGEN data from THL Biobank, after which the data acquired in this study are linked with existing genetic and register data in a controlled FinnGen sandbox environment.

The data acquired in this study is subject to conditions of the IRB protocols and the policies of the Finnish biobank legislation and therefore unavailable for unsupervised usage. The data is stored in FIMM and THL Biobank, where approved researchers can access the data. Eventually the data will also be transferred to the secure FinnGen sandbox environment and linked to the registers available in FinnGen. The results of the study will be published in peer-reviewed journals and presented at scientific conferences.

## Discussion

The TWINGEN study uses biobank registries for a recall study with the aim of identifying individuals at risk of AD. The collected data comprise many known risk factors^36^ and scalable screening methods of AD. While these data are expected to yield novel insights even in isolation, the unique potential for discovery comes from combining the existing follow-up and newly collected data of TWINGEN participants with the existing data.

Since its inception, the FinnGen project has created new expansion areas with the aim of enriching phenotype information. The TWINGEN study addresses this goal by collecting cognitive, physical activity, lifestyle and biological data that are returned to FinnGen via THL biobank, whereas FinnGen would allow for a registry-based follow-up of the TWINGEN study participants.

## Author contributions

TTS: writing the original draft, revision and data collection. EV: writing the original draft, revision, resources, concept, design and supervision. AA: writing, revision and data collection. IR, HL, S-KH, SK, TK, SA: writing. MO, P-IM: revision. AK: design, administration and revision. AT: study coordination and revision. TV, HV-Y, MM, RS, PN, SS: resources and revision. V-MK, TK, EP, MR: resources. JK: concept, design, revision and resources. MU: study coordination, data collection. VJ: concept, design, revision, supervision. MMF, MH, JL, RK: consultation in the design of the study. HR: concept, design, revision. AP: concept, resources, revision. TP: data management. AJ: administration. TS and EV contributed equally to this paper.

## Data Availability

Not applicable

## Acknowledgements

We thank THL Biobank and biobank directors Eero Punkka (Helsinki Biobank) and Veli-Matti Kosma (Biobank of Eastern Finland) for providing resources to carry out this study. Biobank personnel who have collected the data and processed the samples: Sabrina Belgasem (Helsinki Biobank), Henna Palin, Minttu Virolainen and Anna-Kaisa Pohjonen (Finnish Clinical Biobank Tampere), Anu Outinen-Tuuponen, Marja-Leena Kytökangas, Riikka-Mari Siiro-Virtanen (Arctic Biobank and Biobank Borealis of Northern Finland), Senni Lipponen (Central Finland Biobank), Nina Hurula (Biobank of Eastern Finland) and Heidi Kalve, Anniina Friman and biomedical laboratory scientist students from the Turku University of Applied Sciences’ clinical laboratory. The participants of the TWINGEN study were recruited through THL Biobank (study number THLBB2022_83). We thank Jyrki Tammerluoto and Steffi Besselink for legal services and Huei-Yi Shen for administrative work. We thank the participants of the Finnish Twin Cohort study for their participation to TWINGEN and all previous data collection waves.

## FinnGen Collaborators

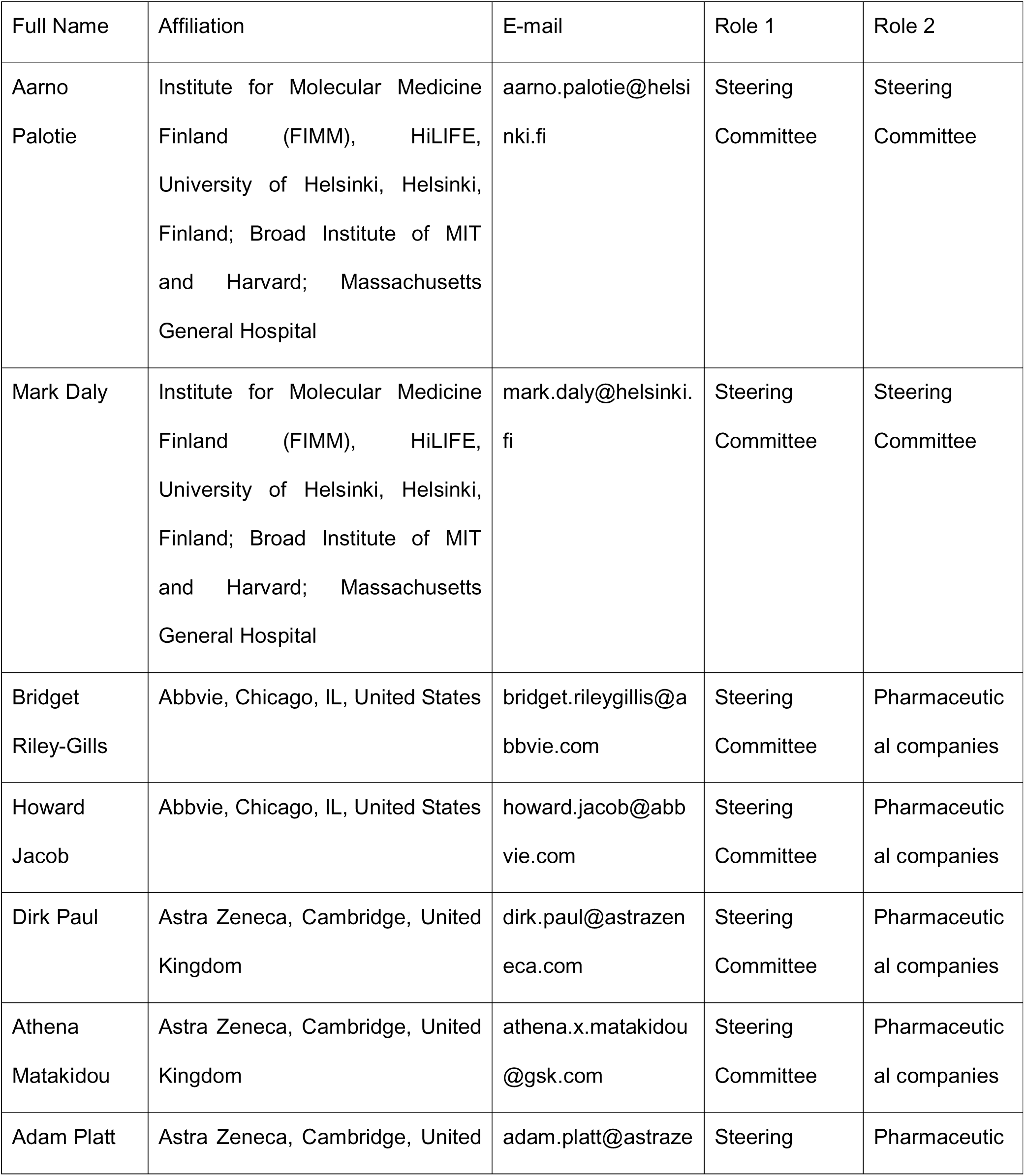
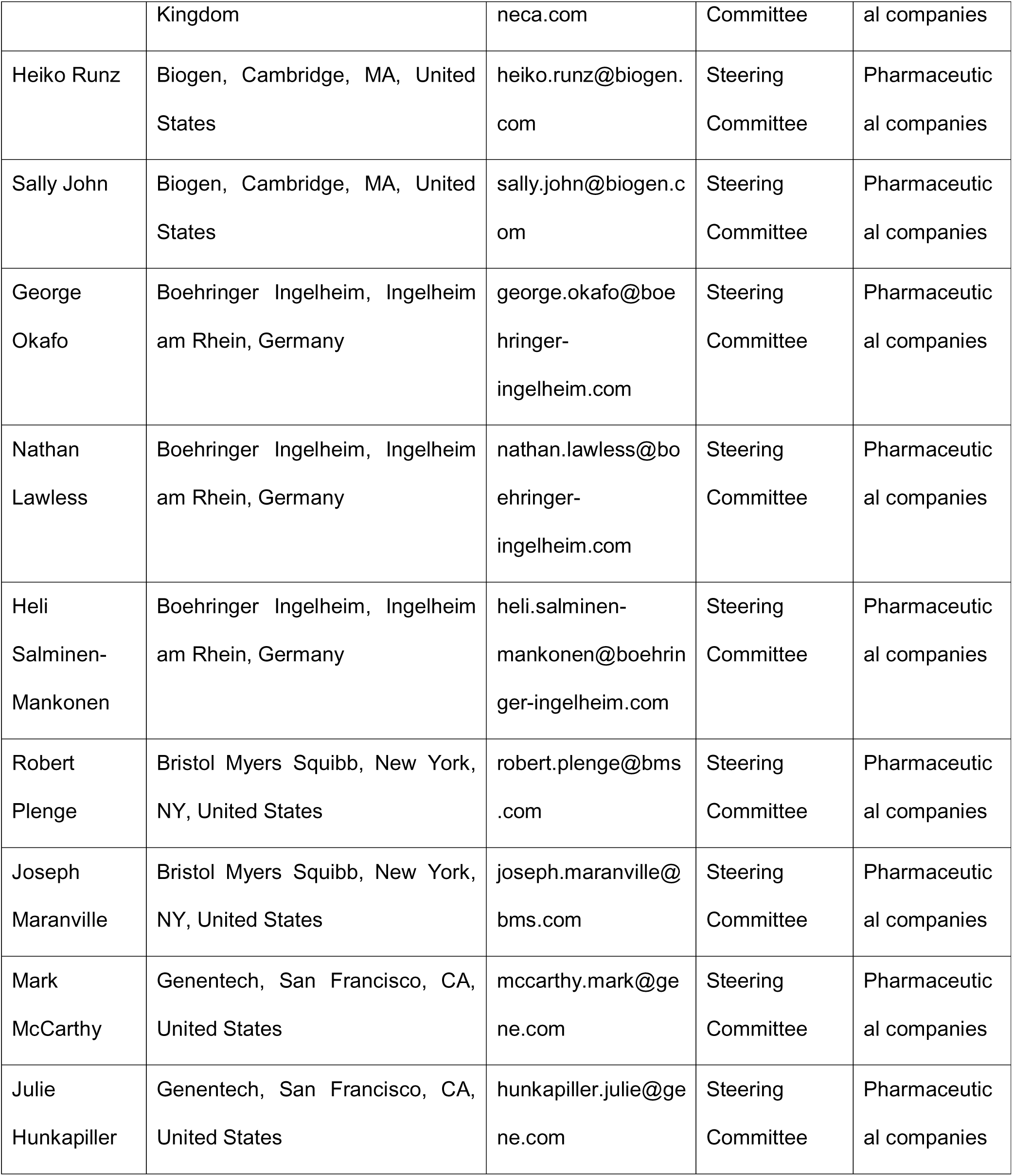
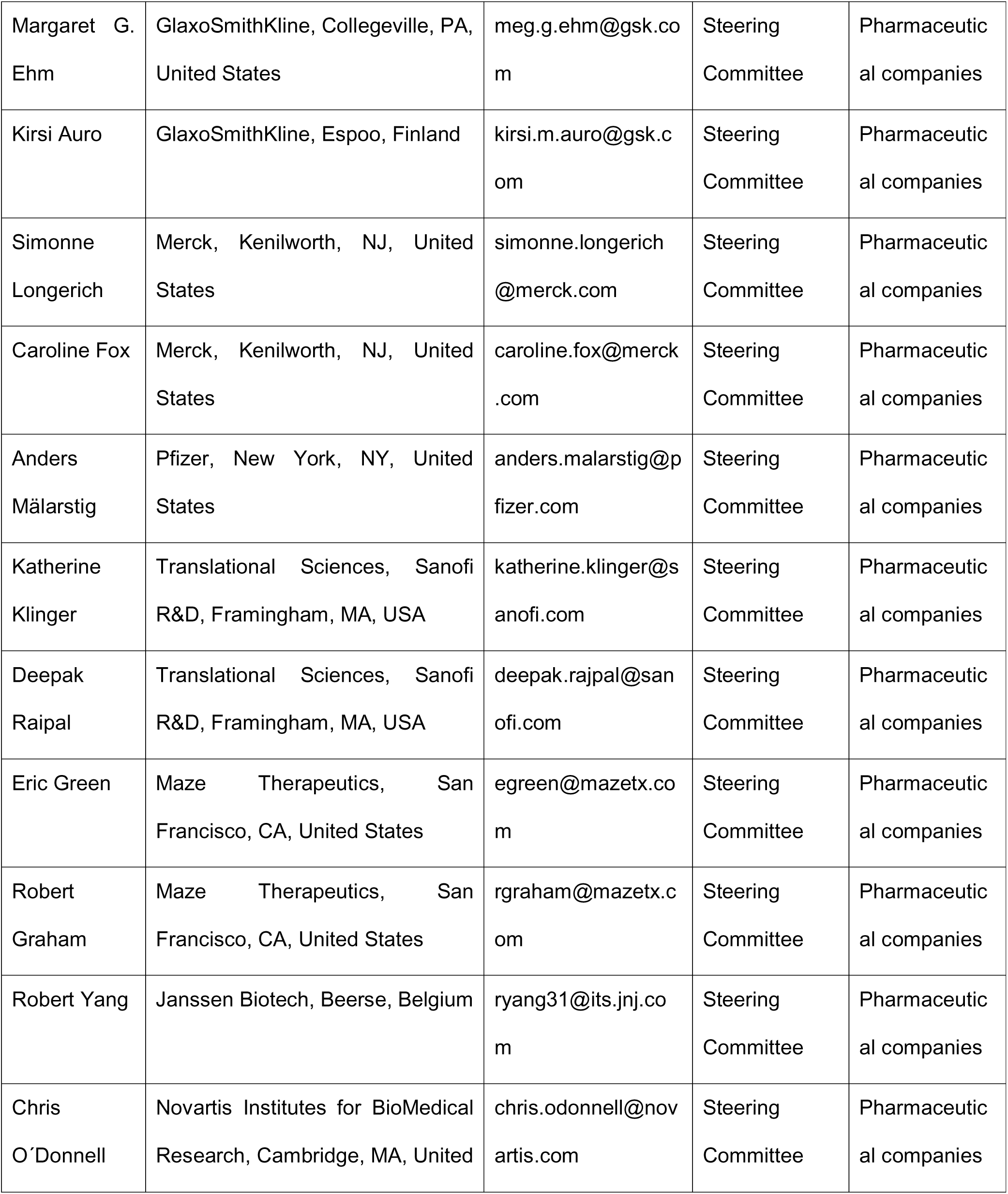
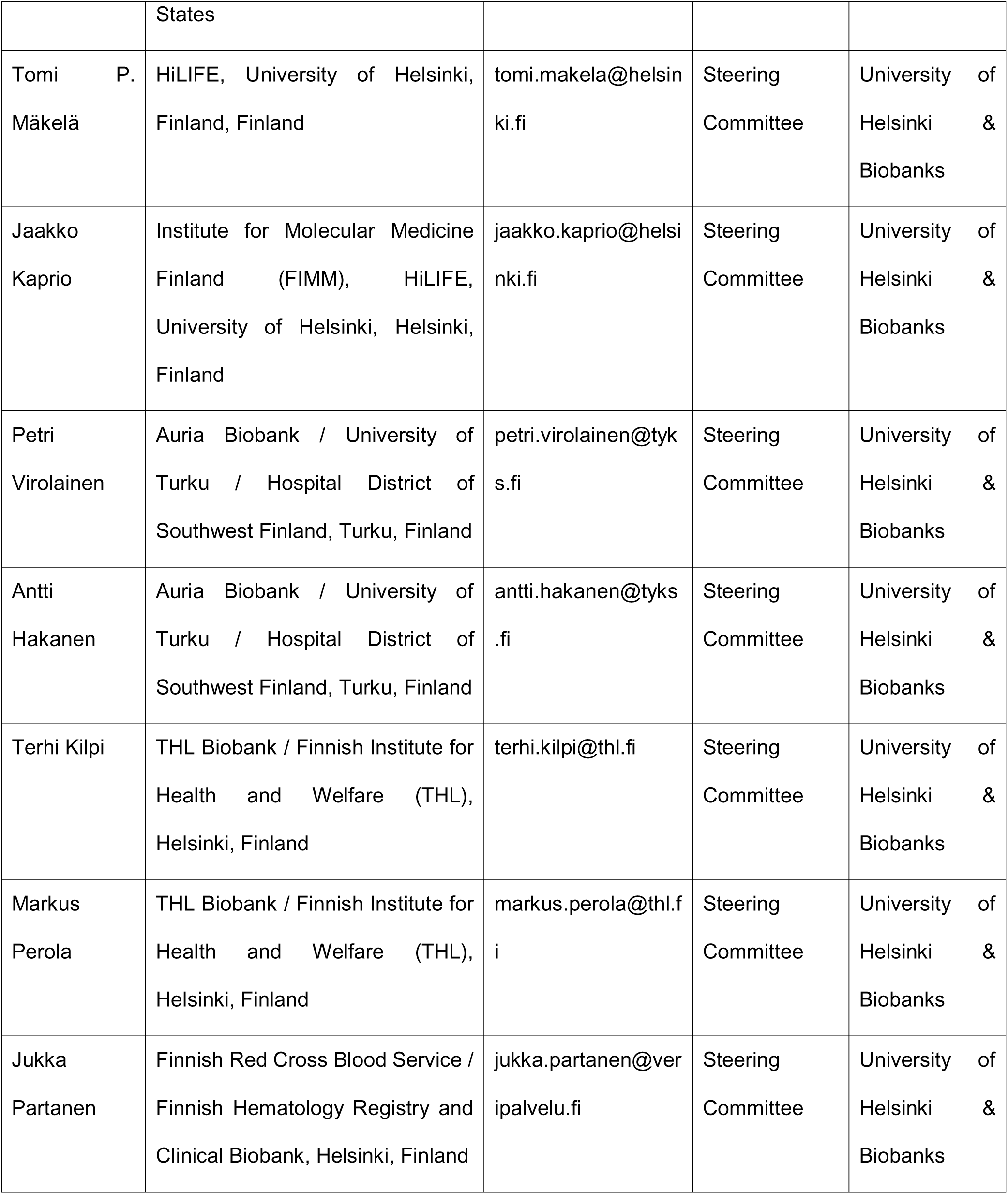
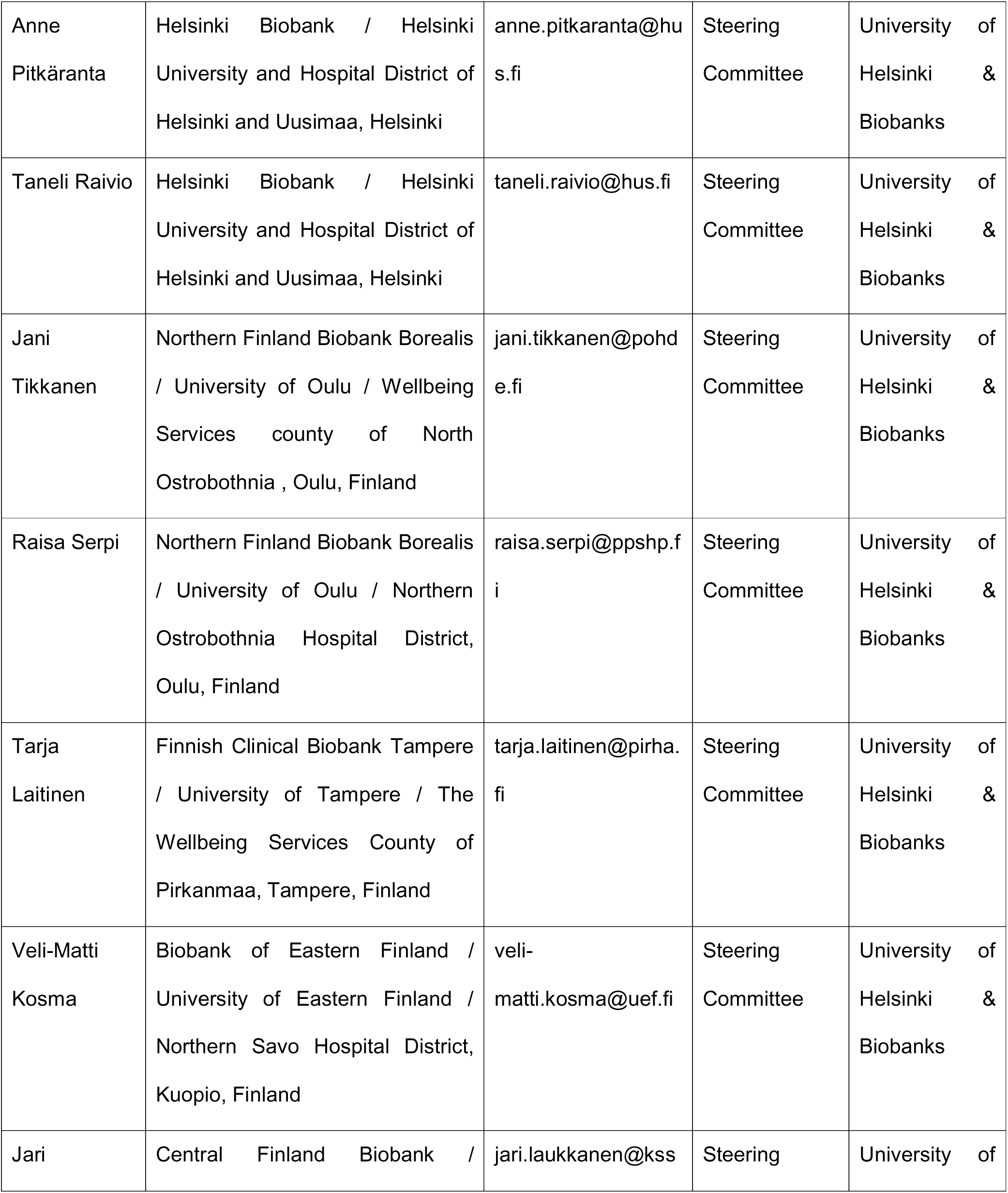
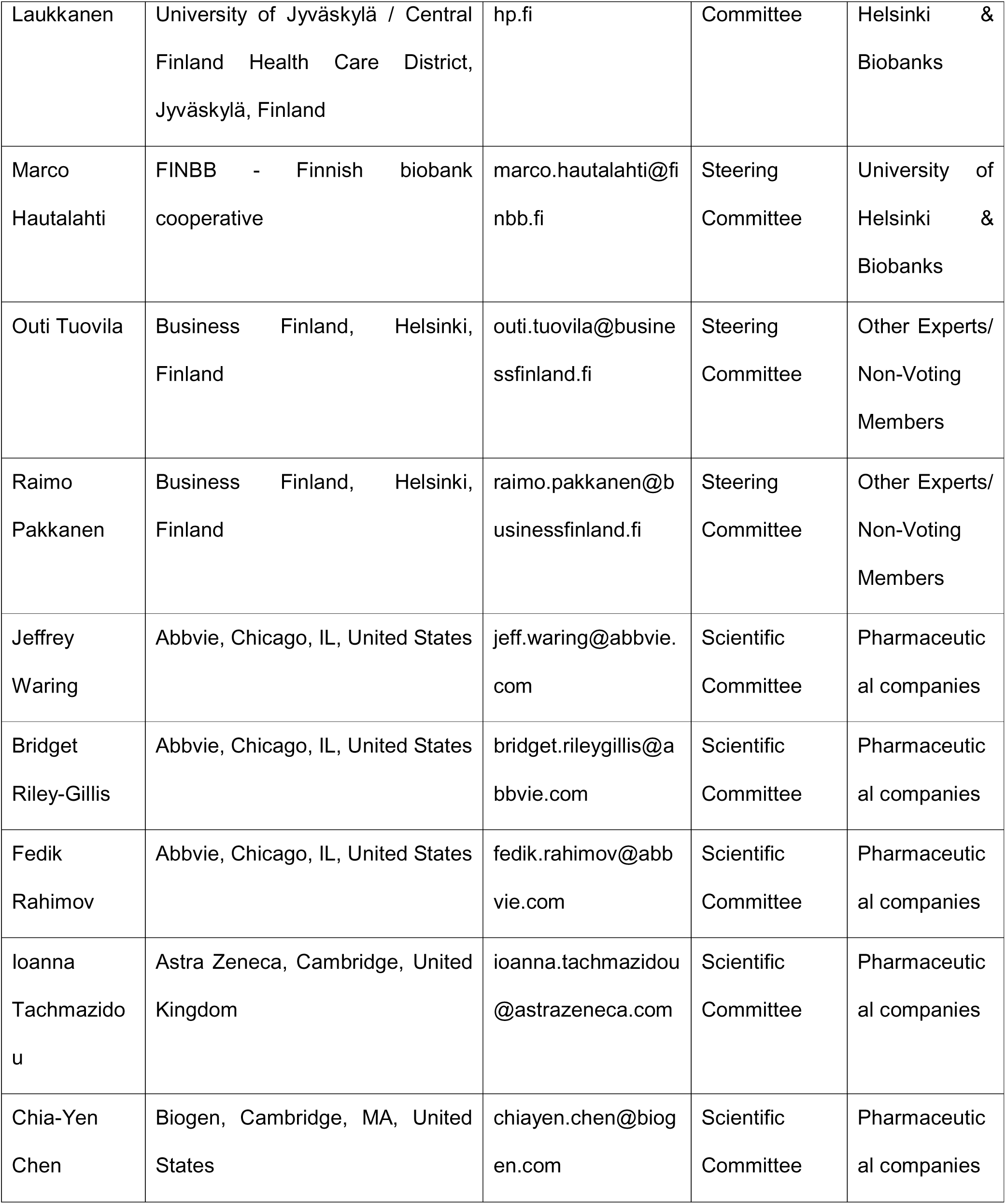
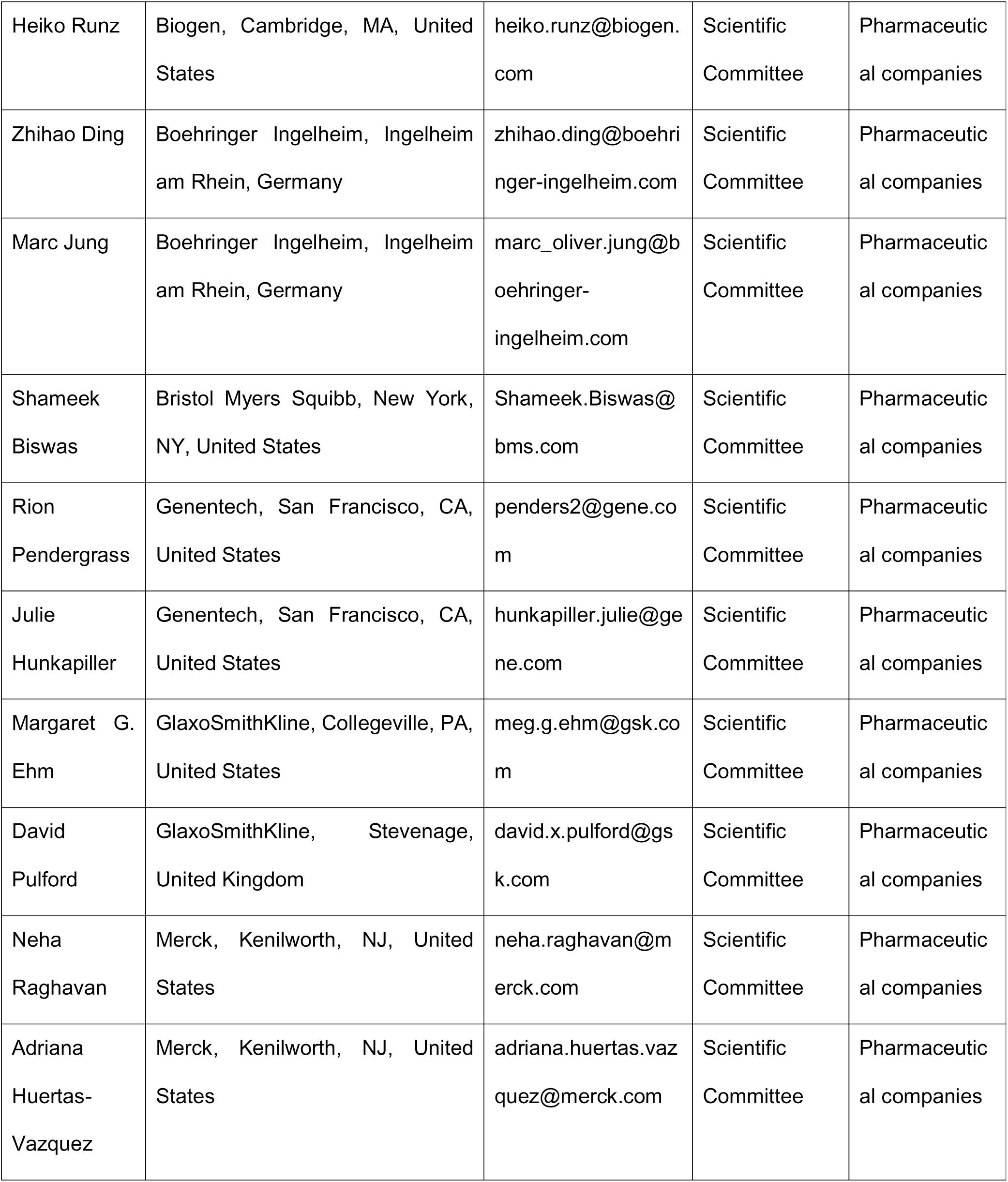
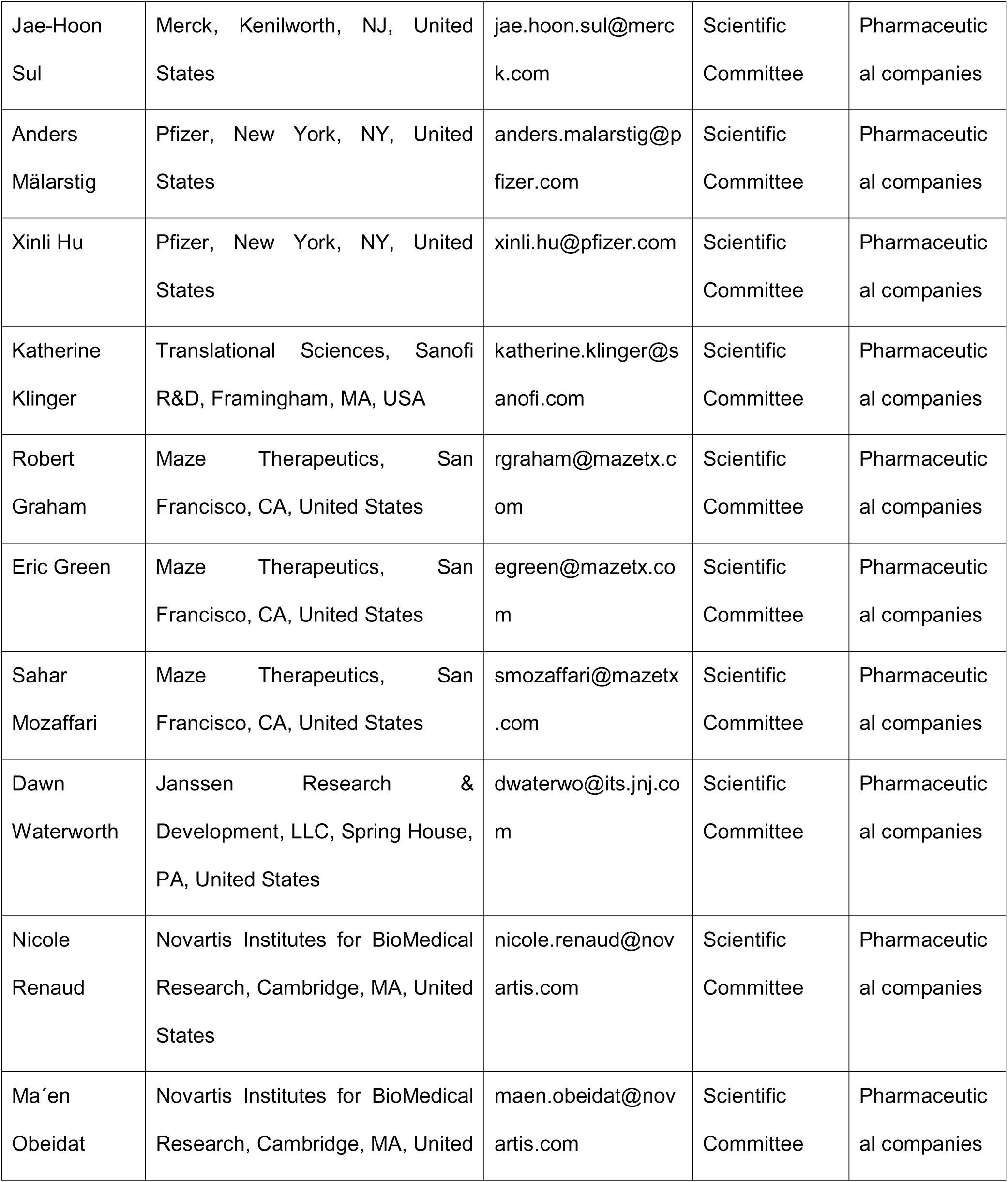
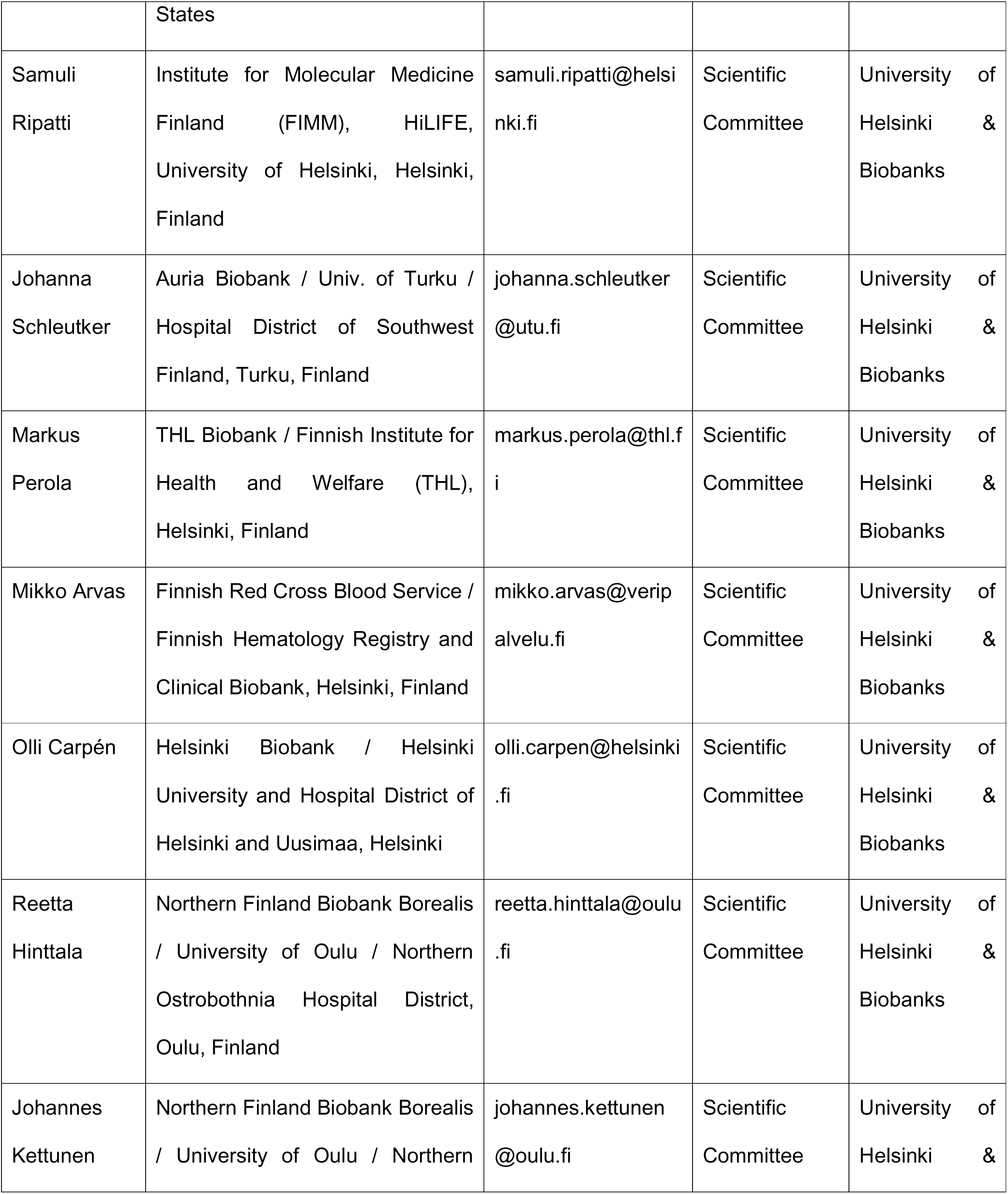
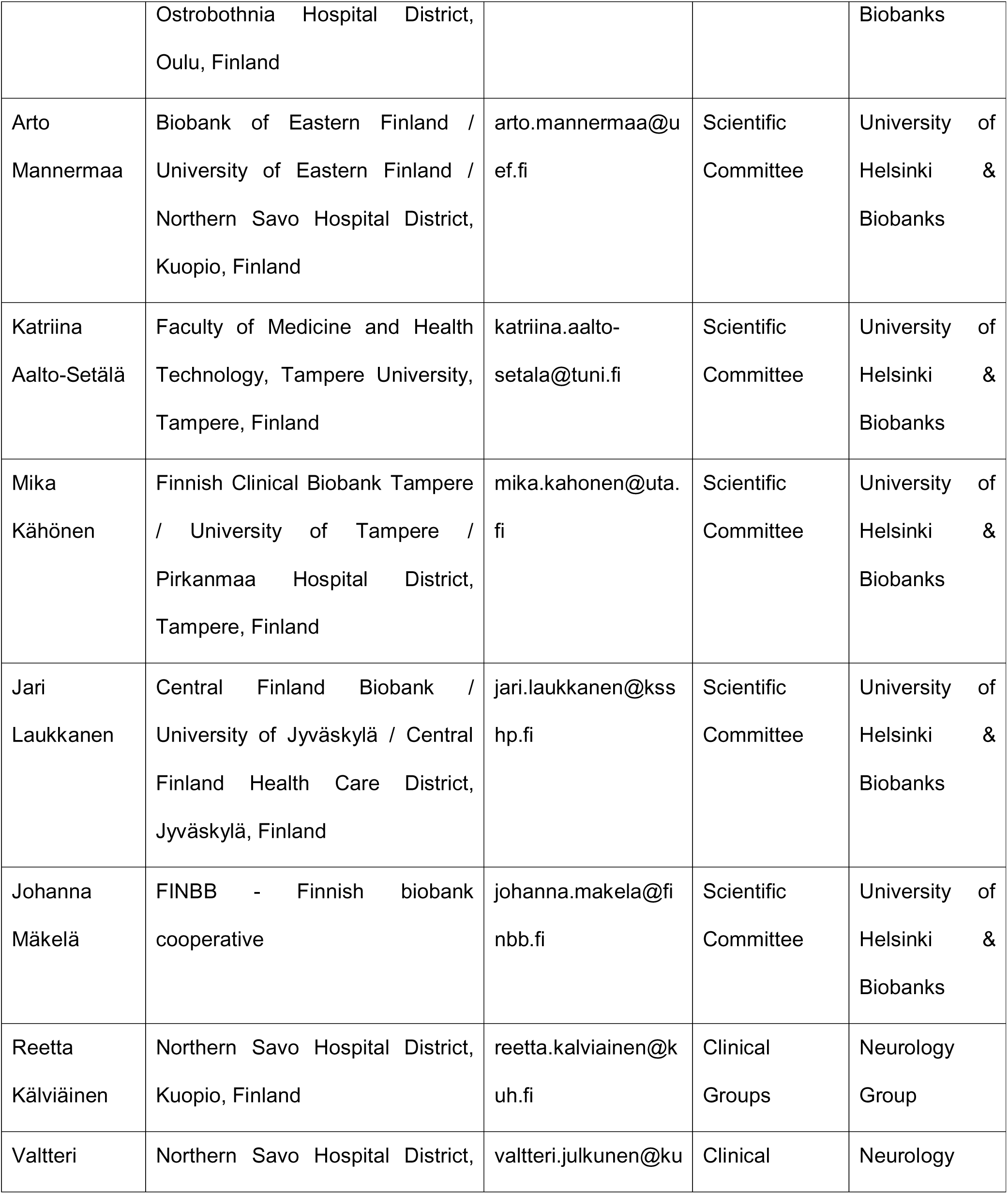
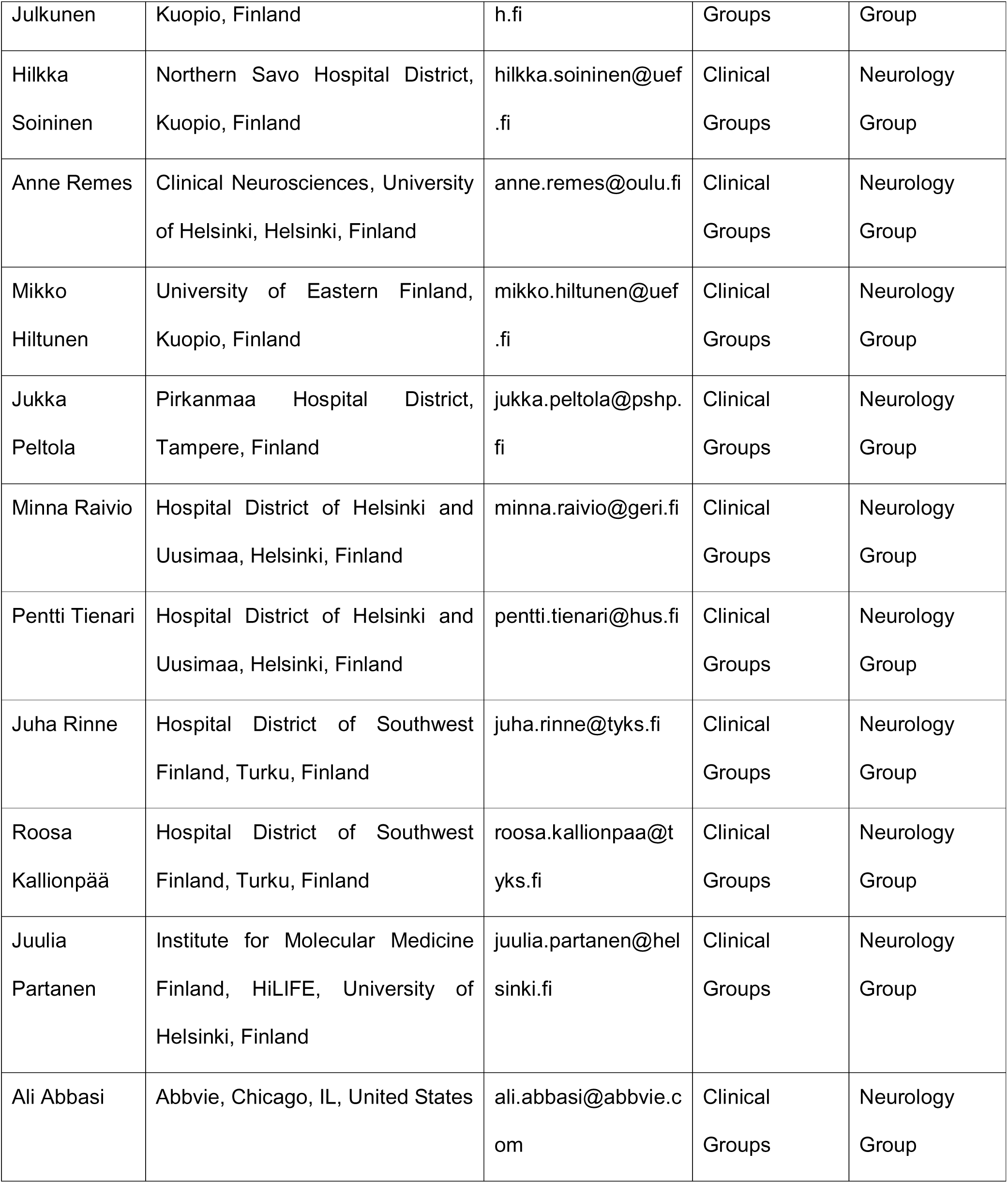
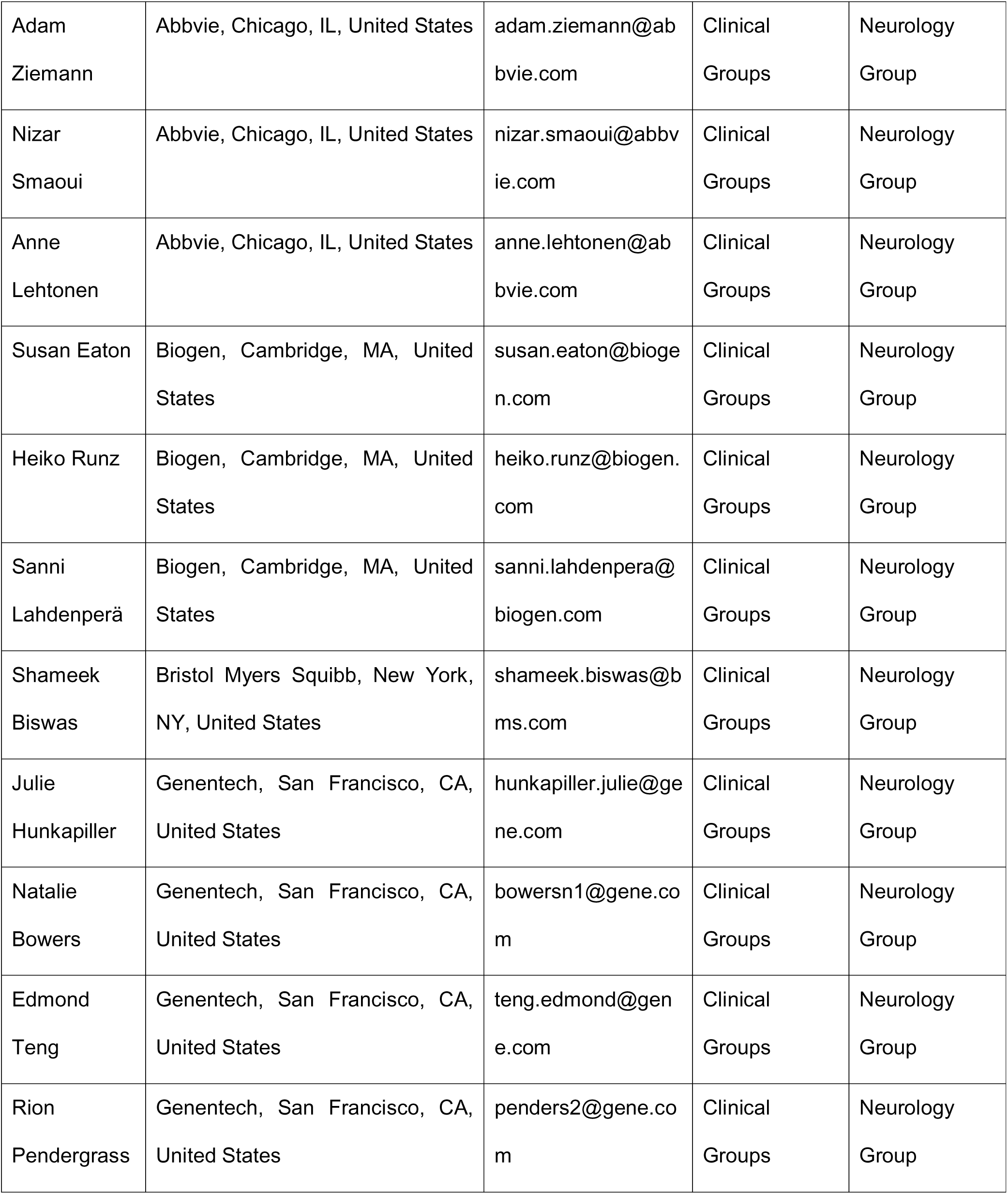
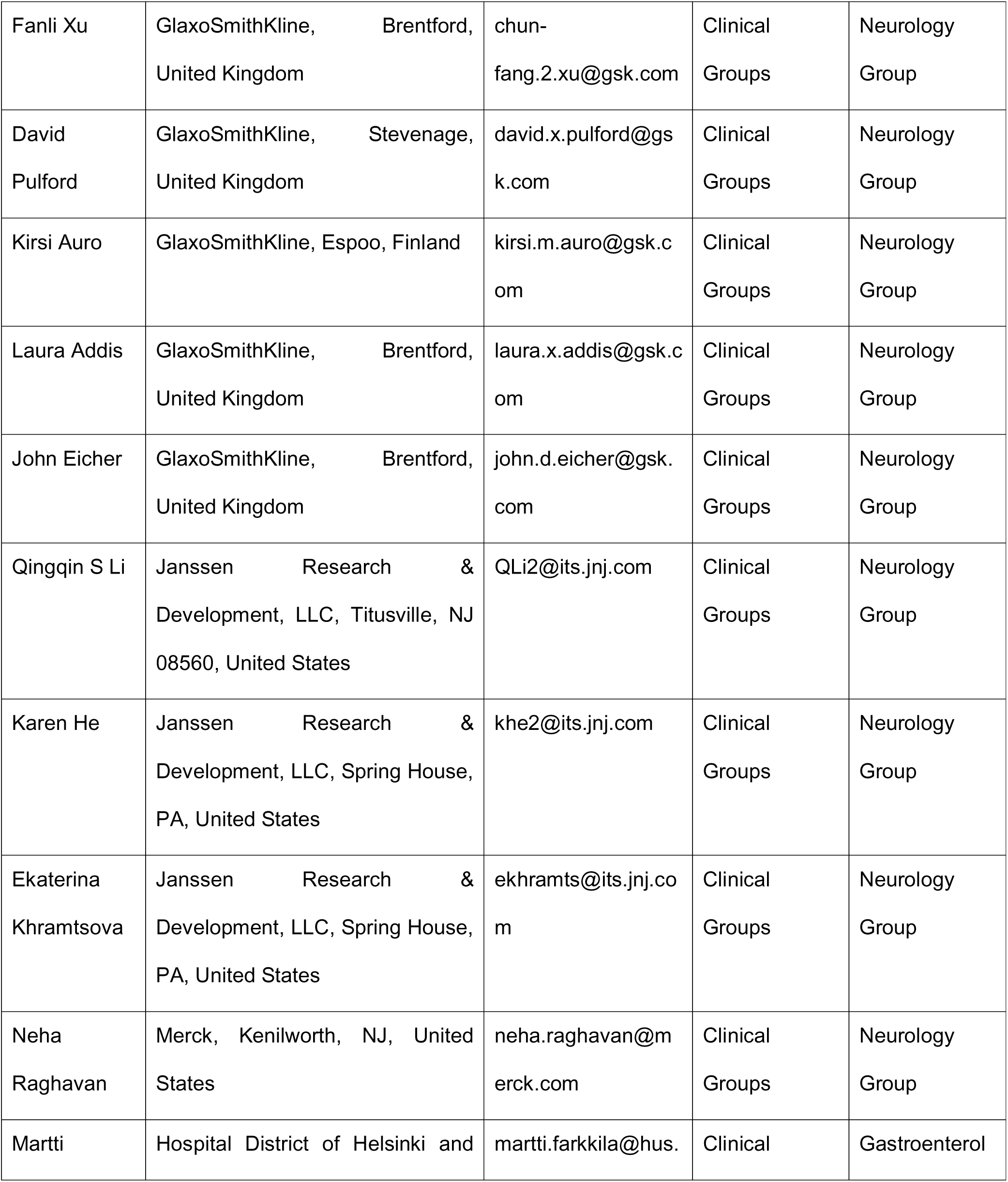
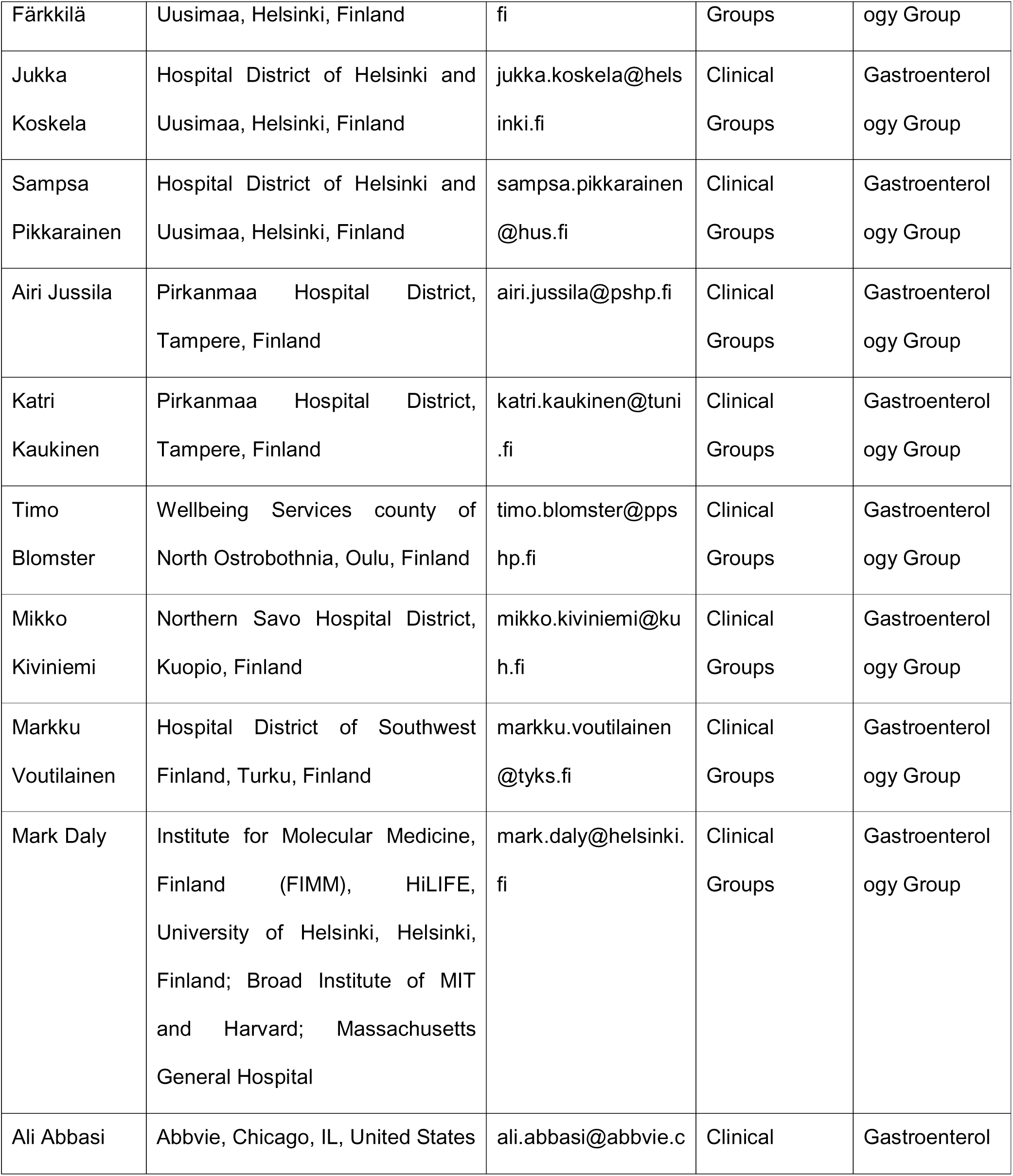
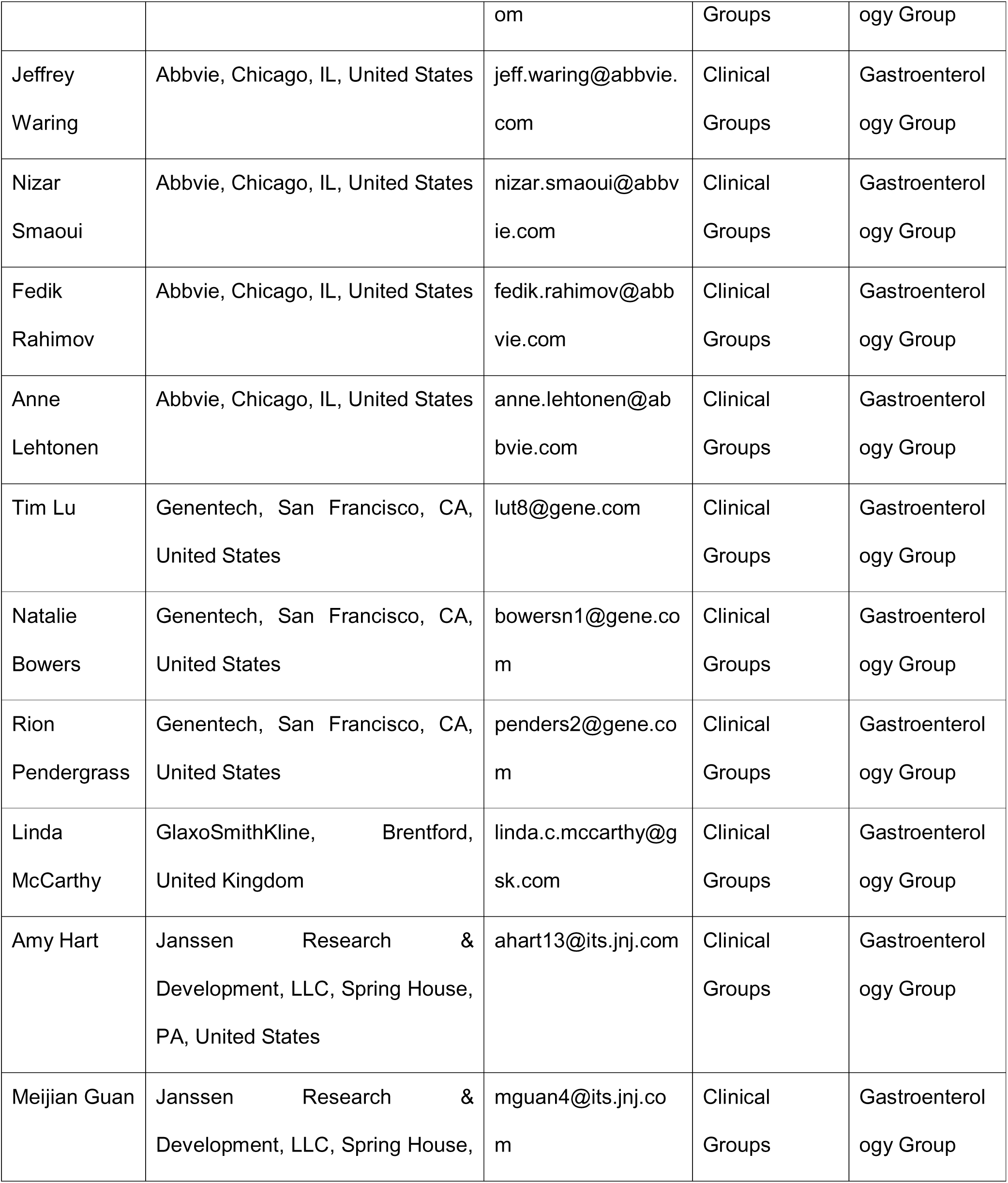
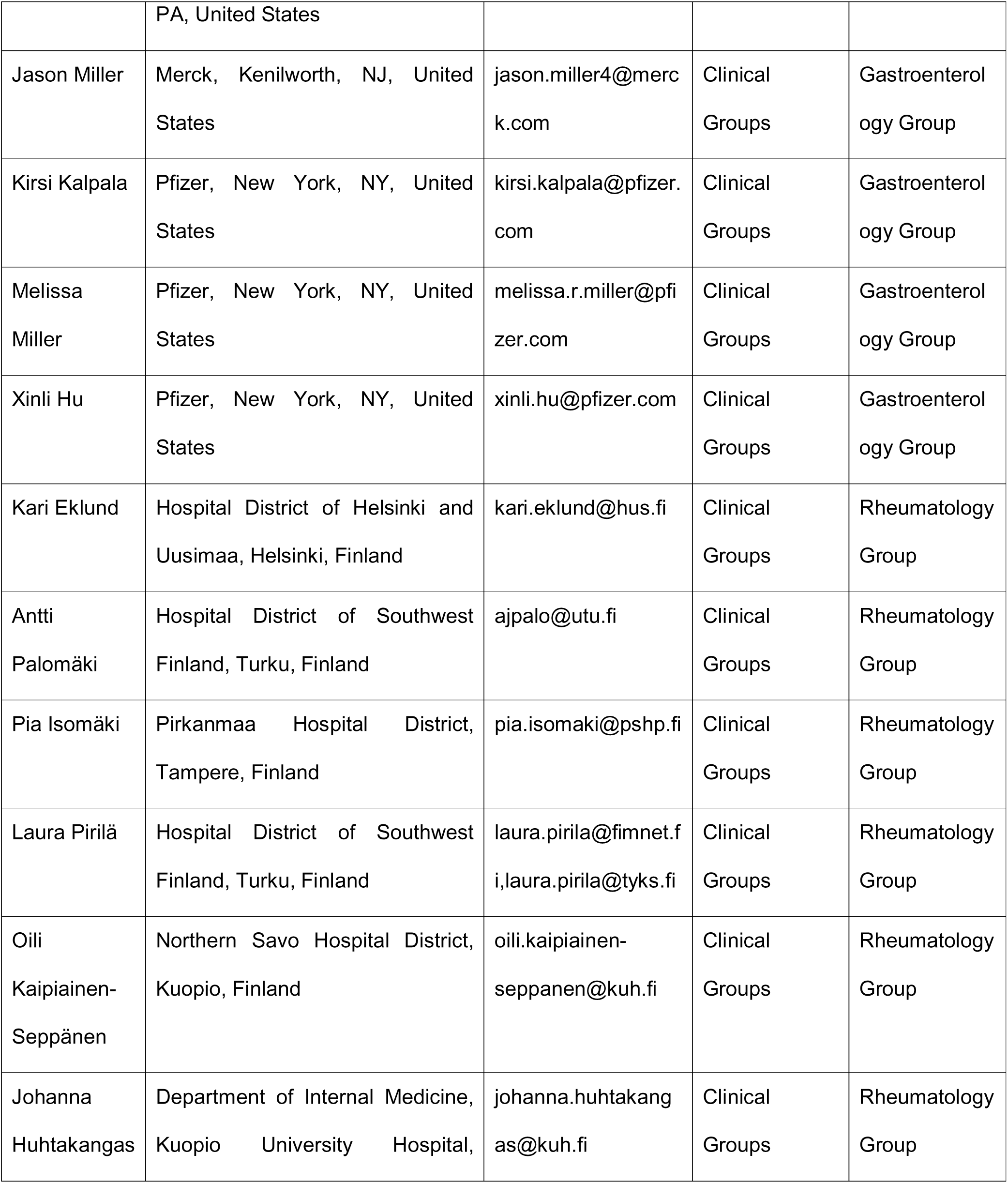
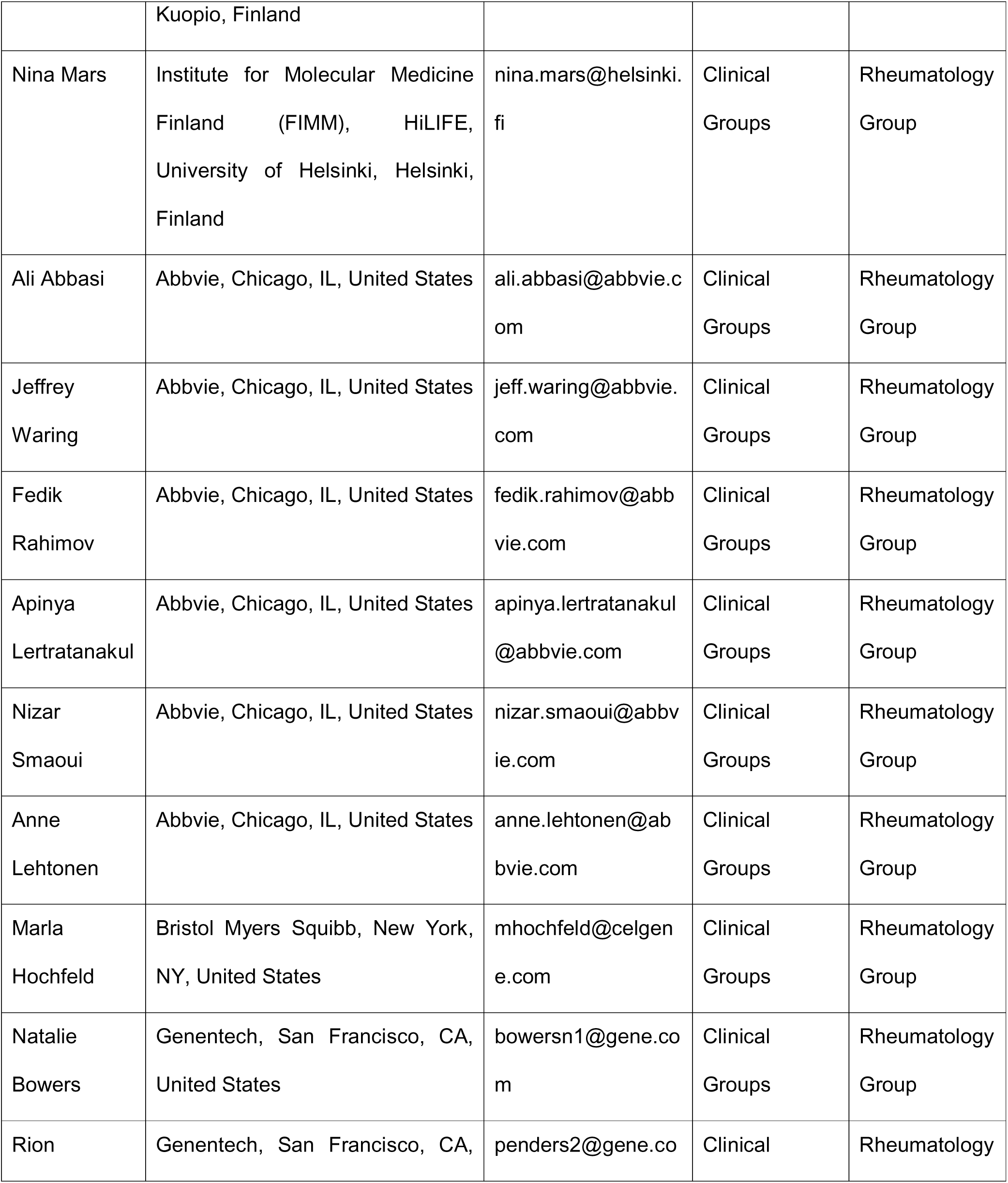
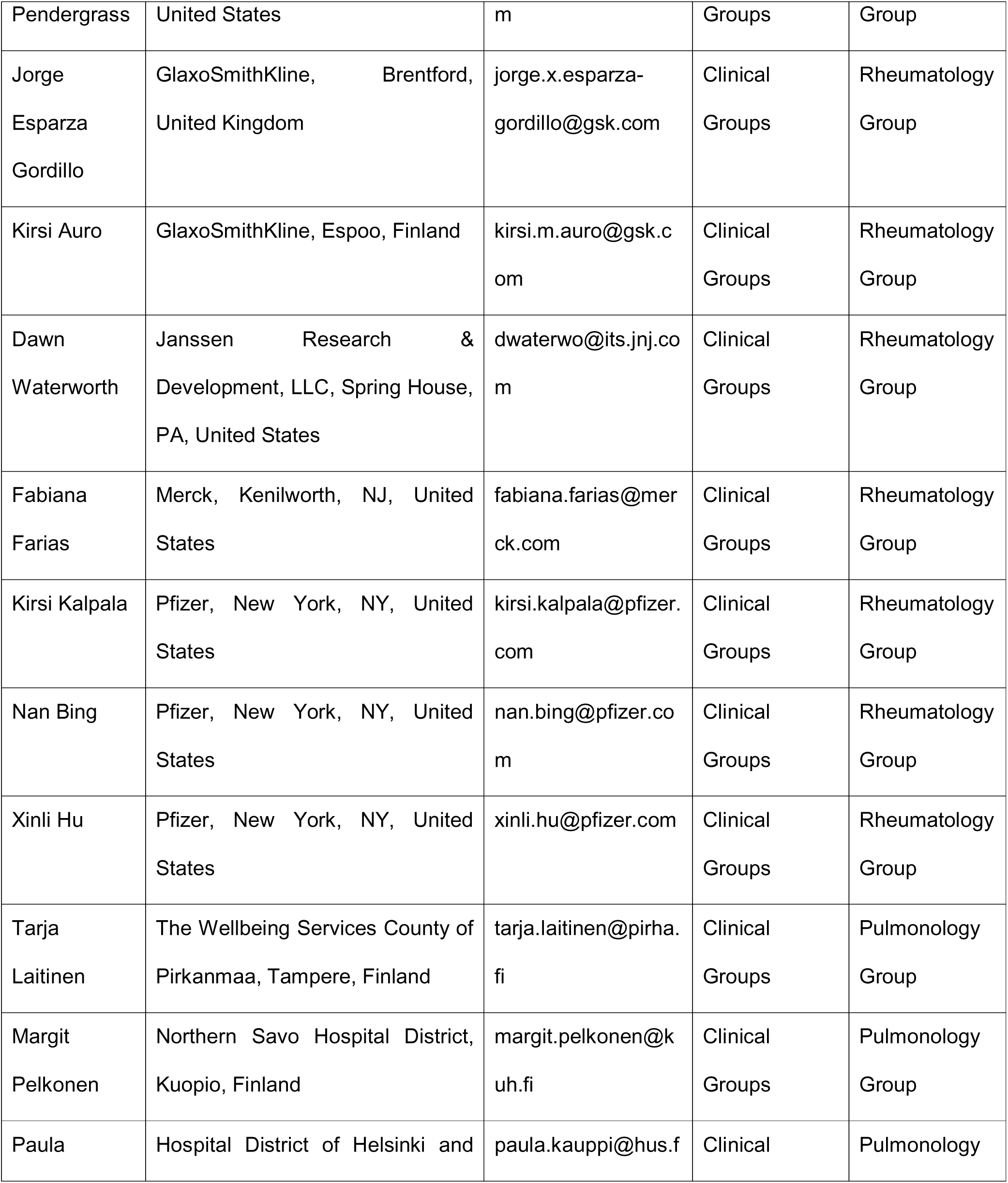
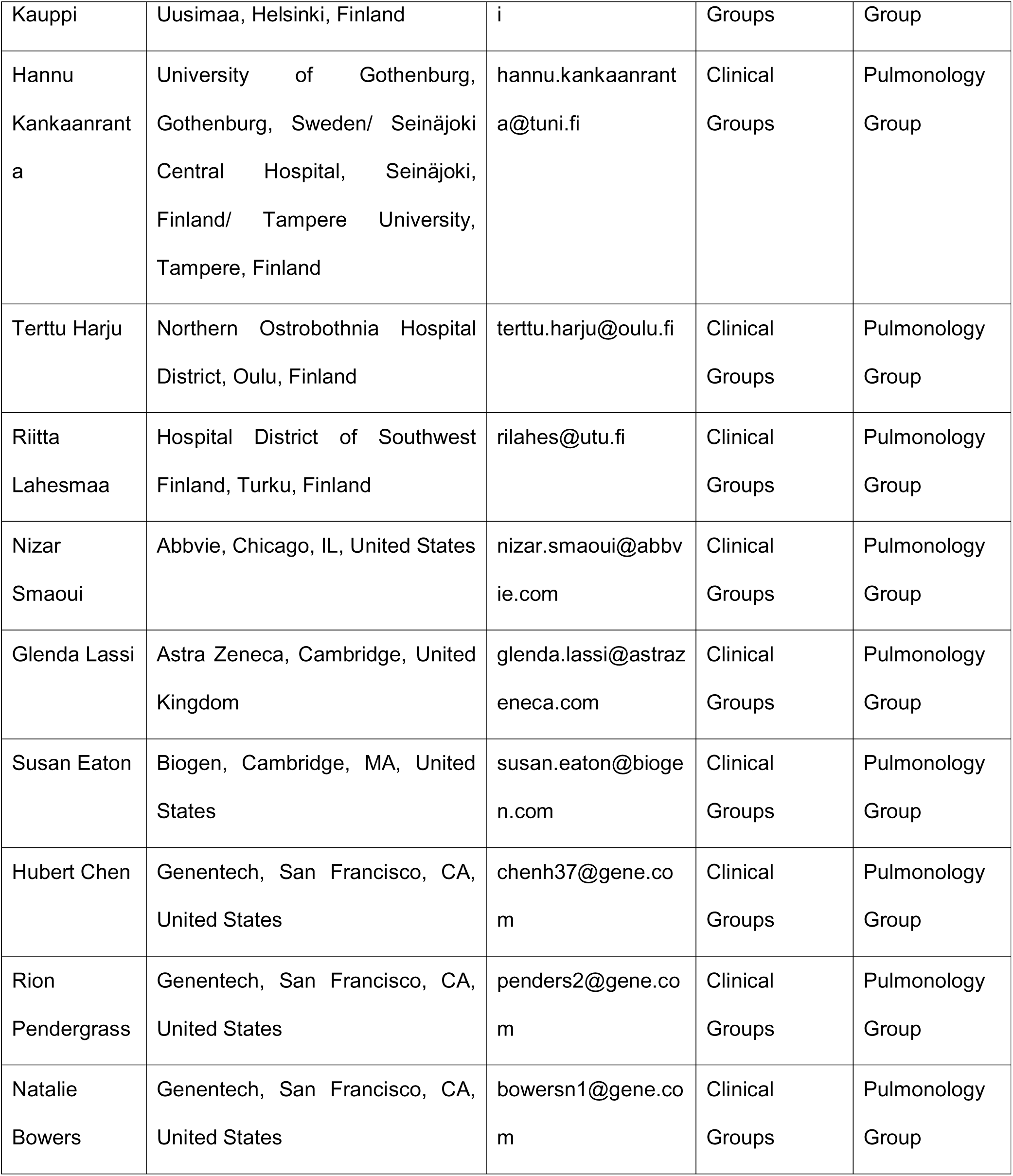
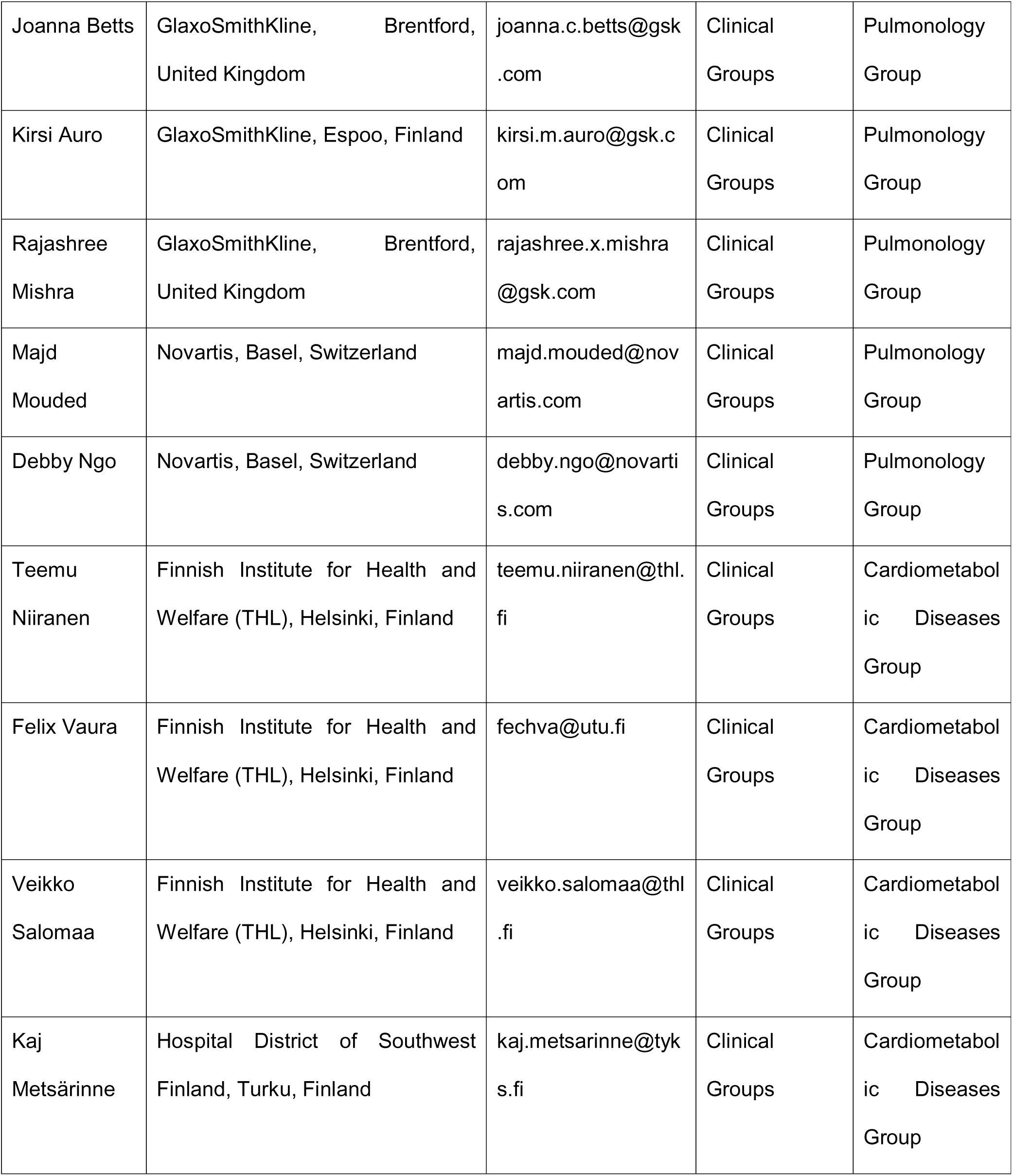
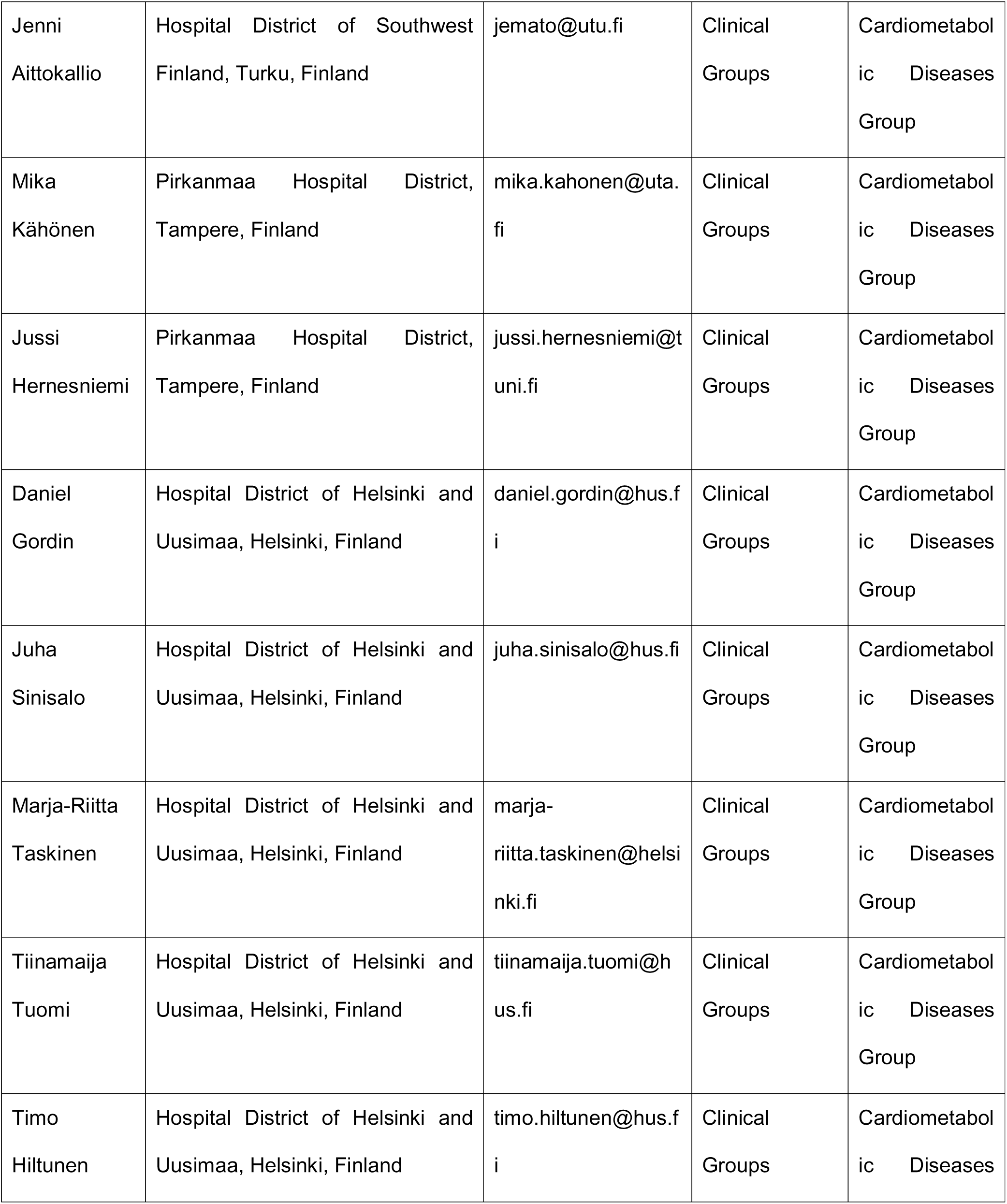
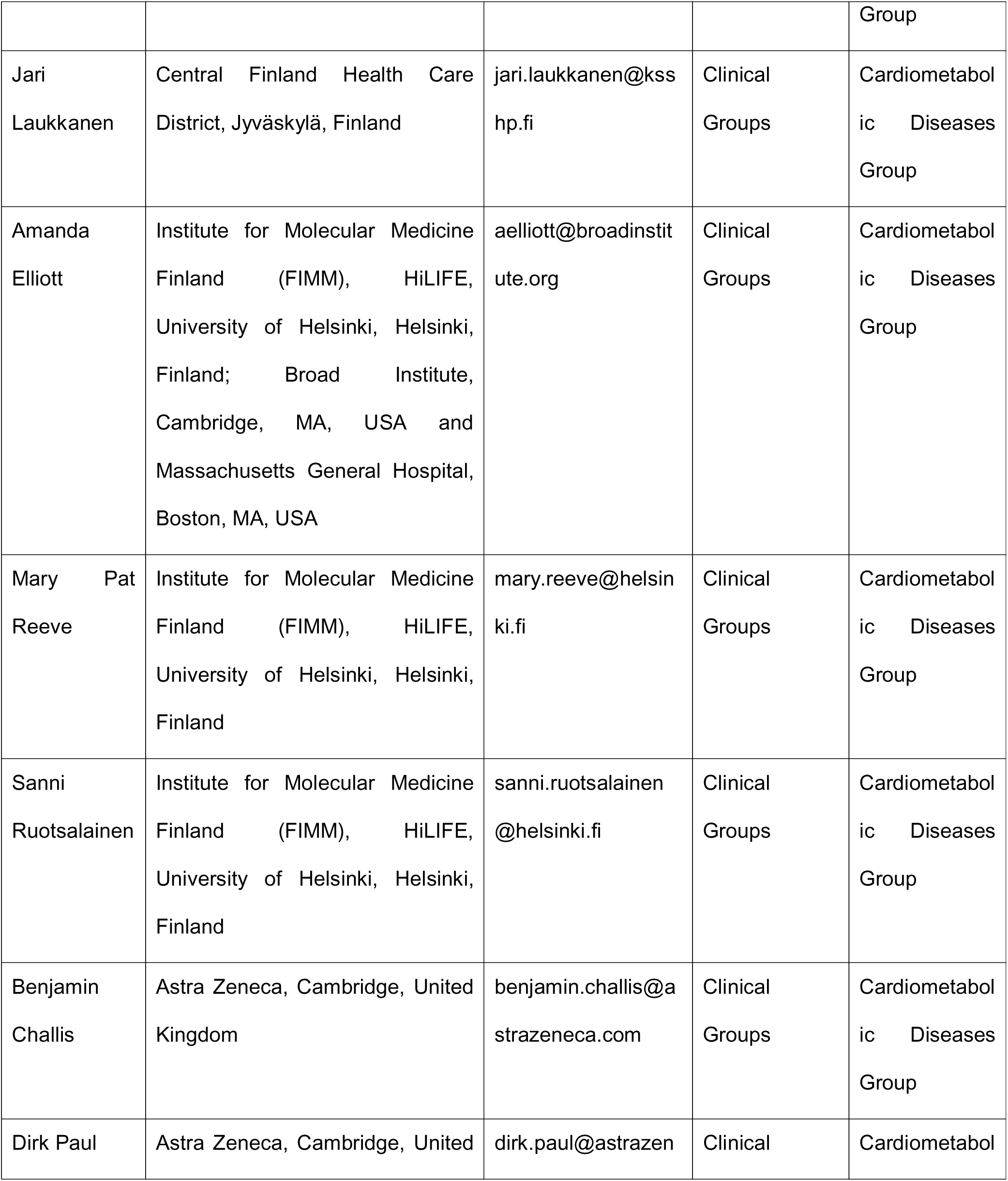
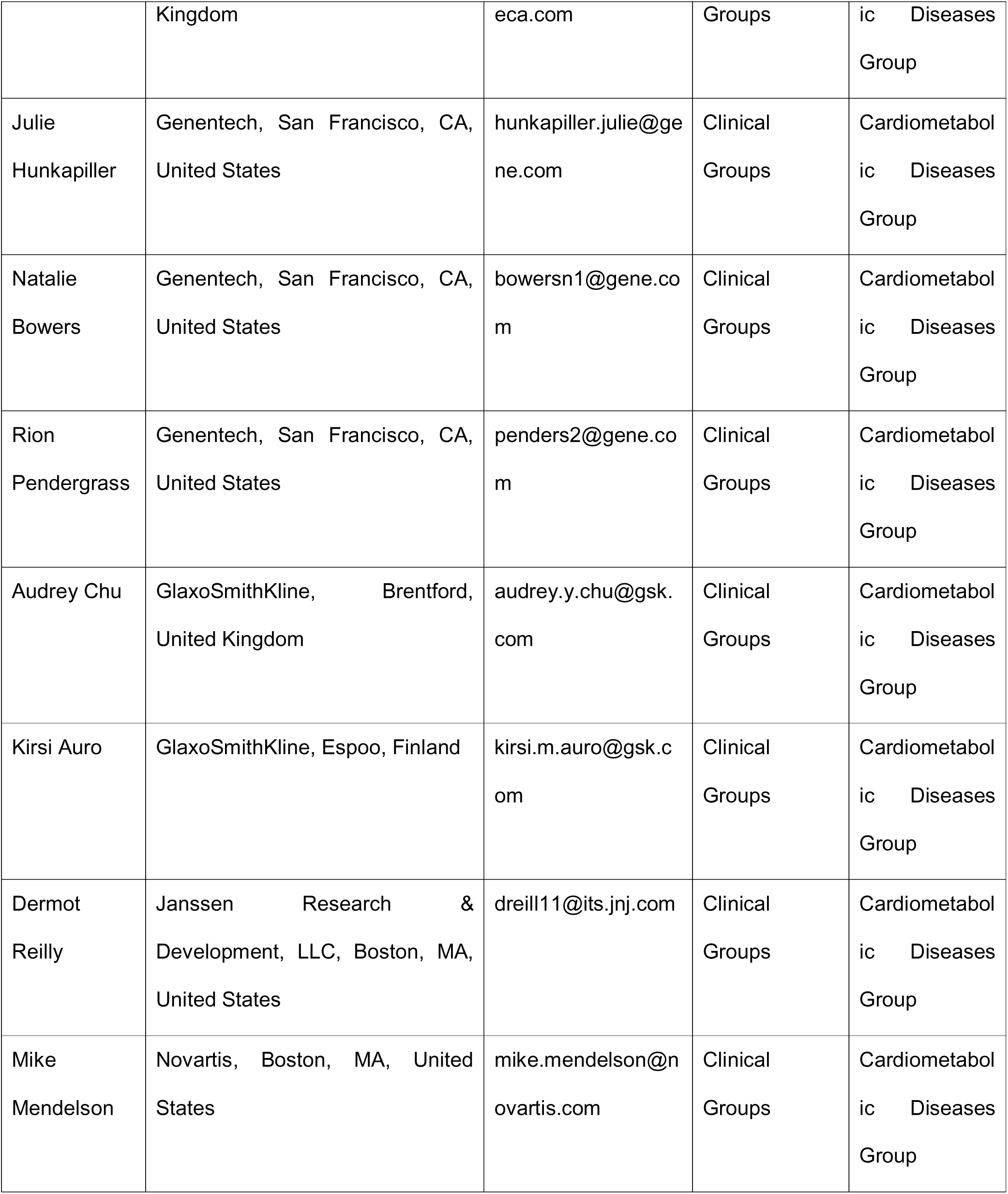
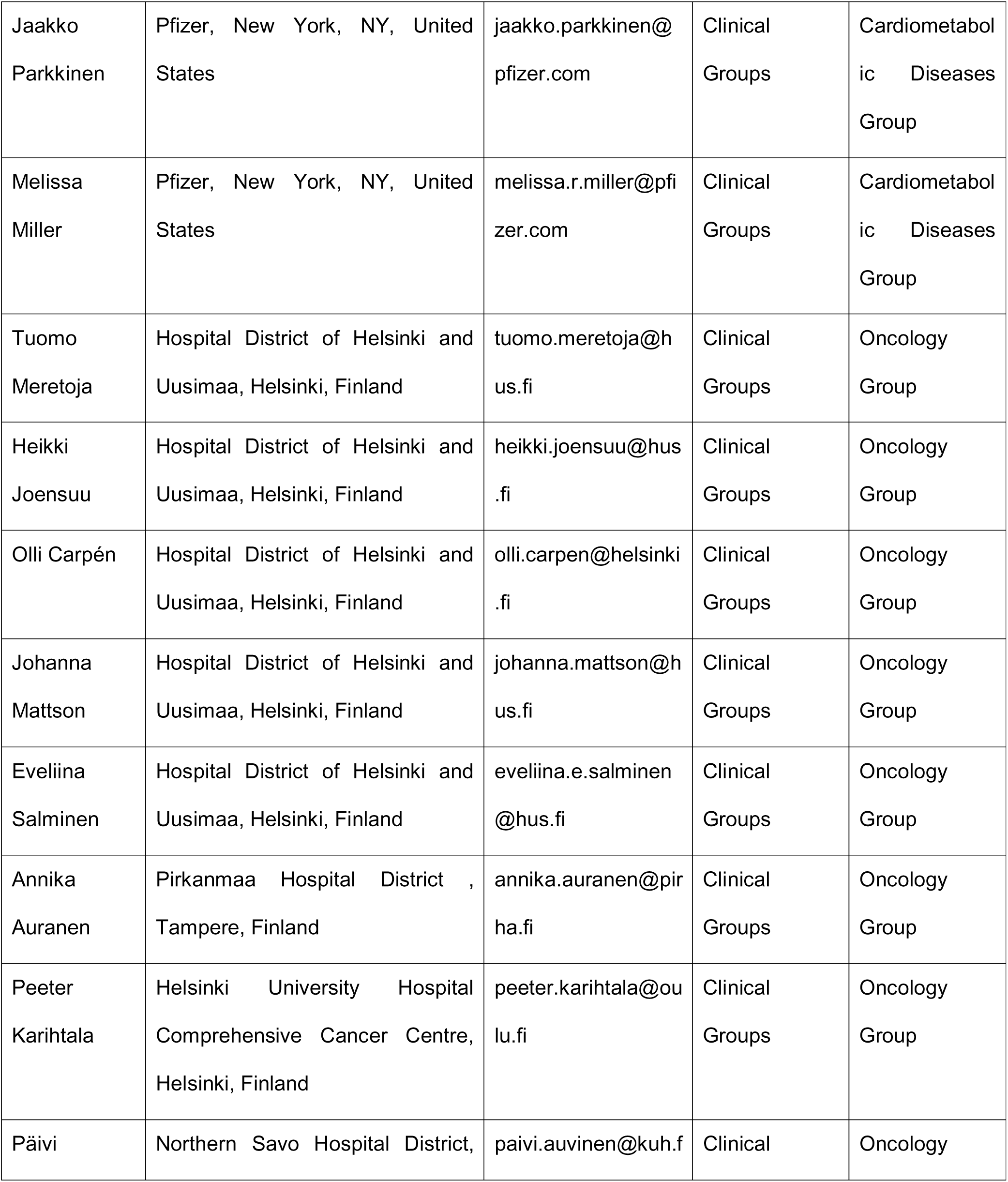
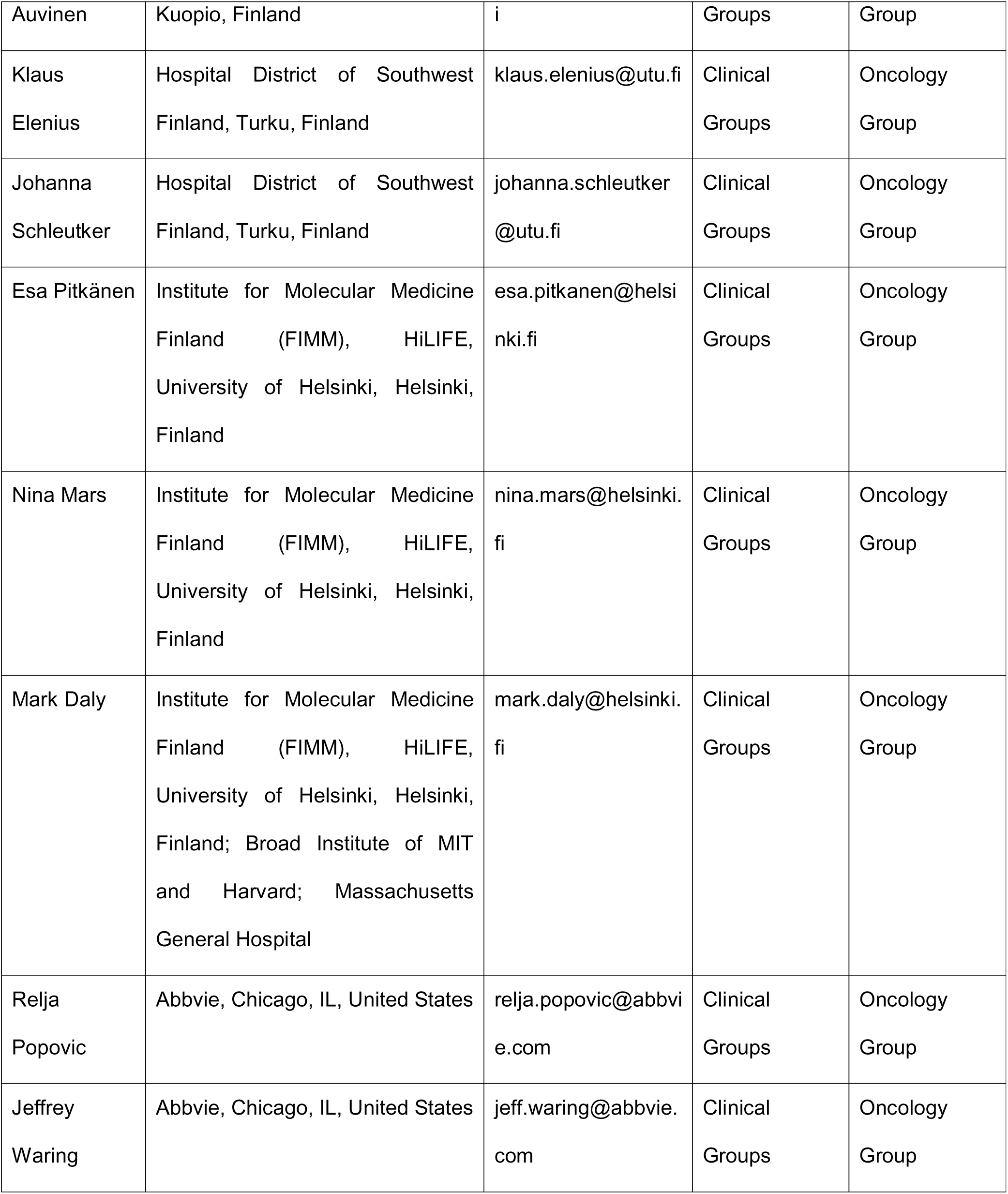
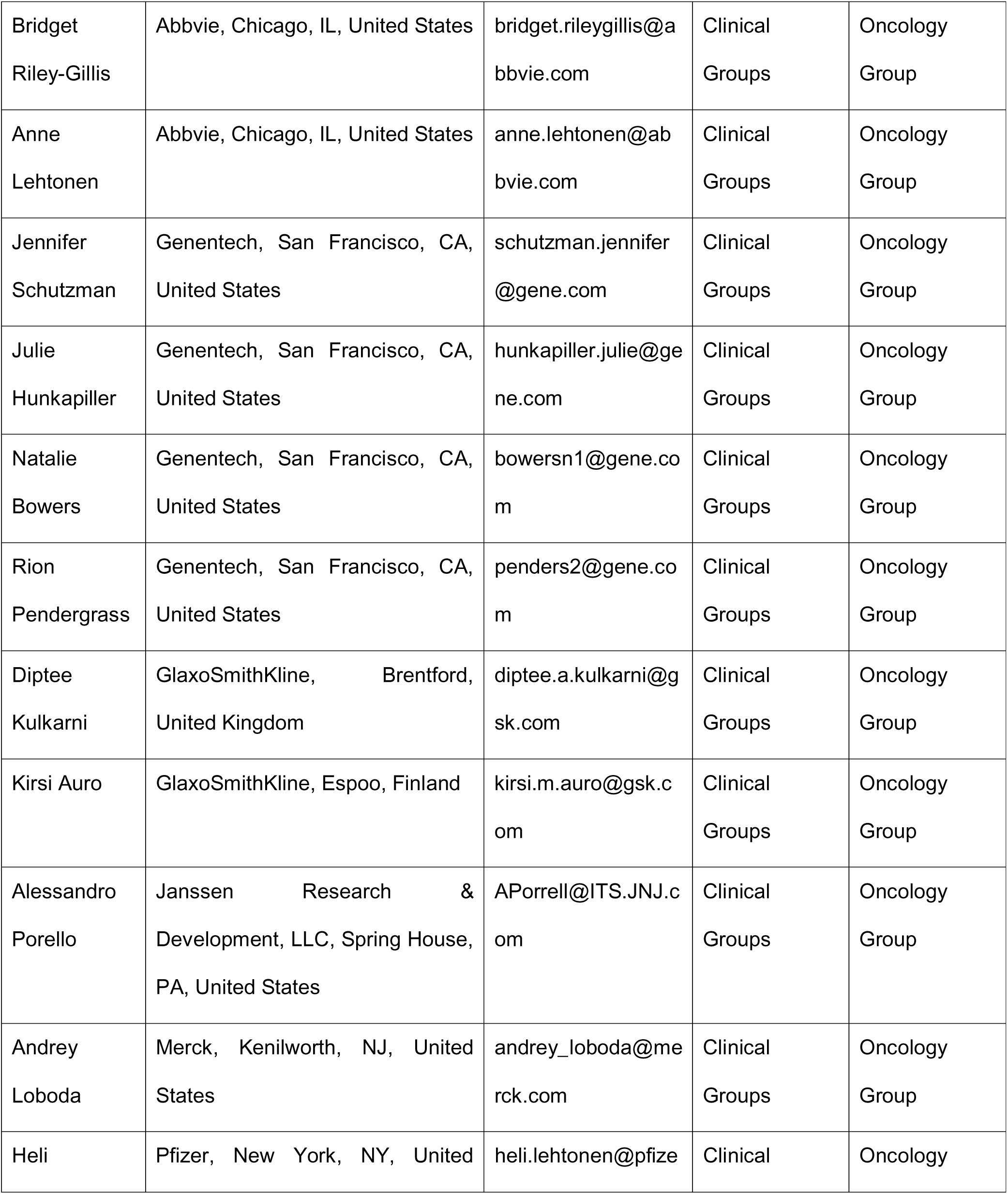
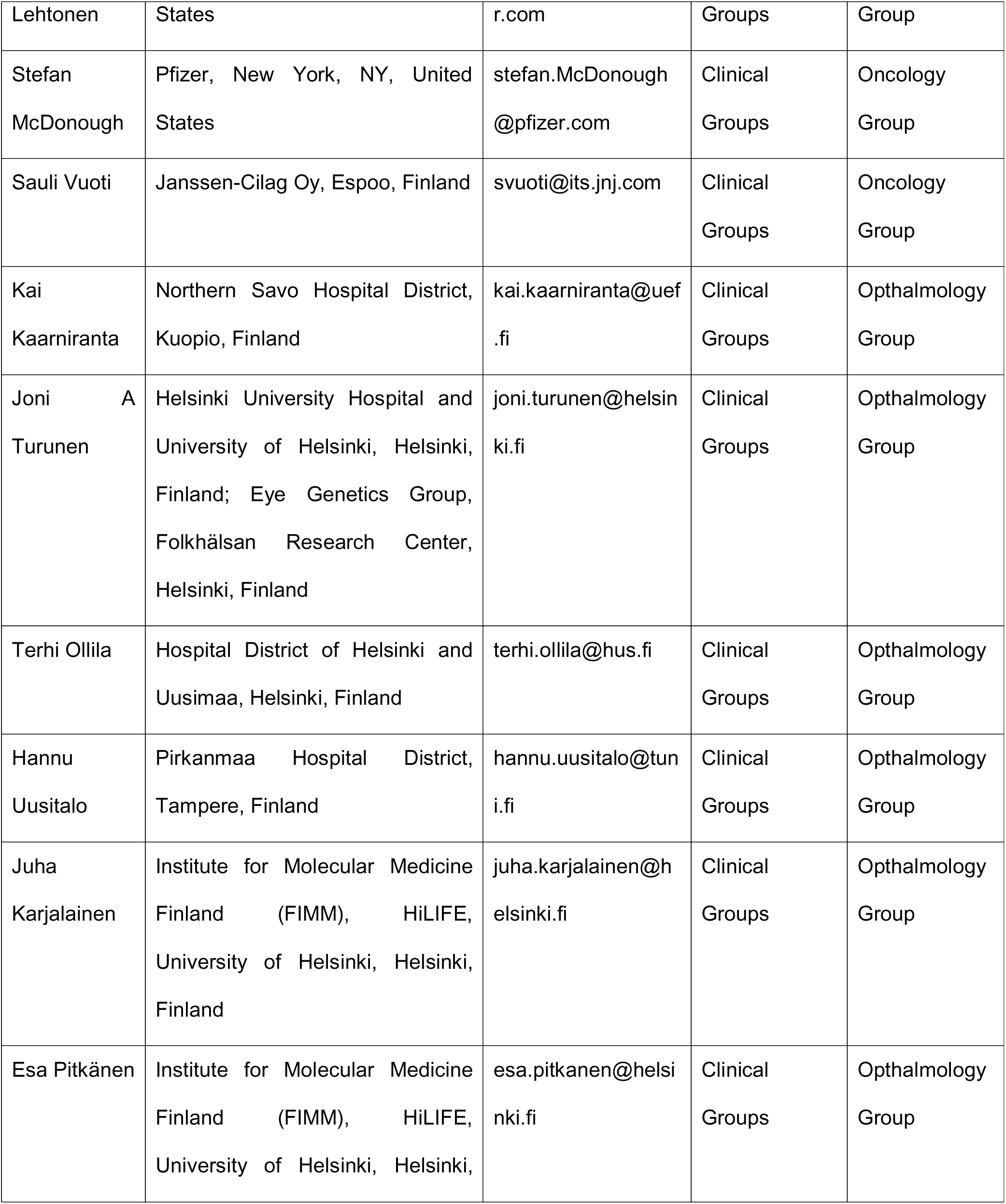
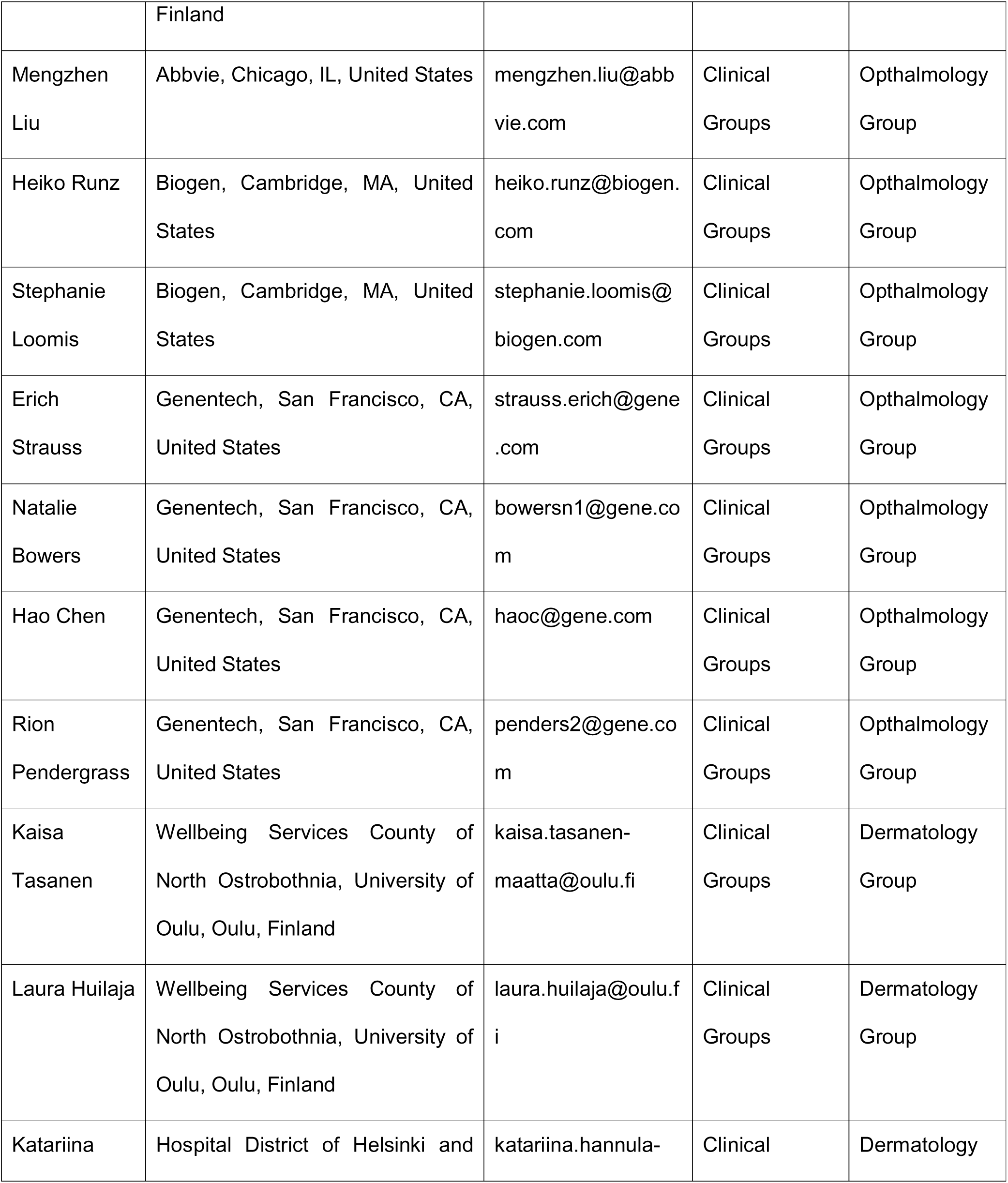
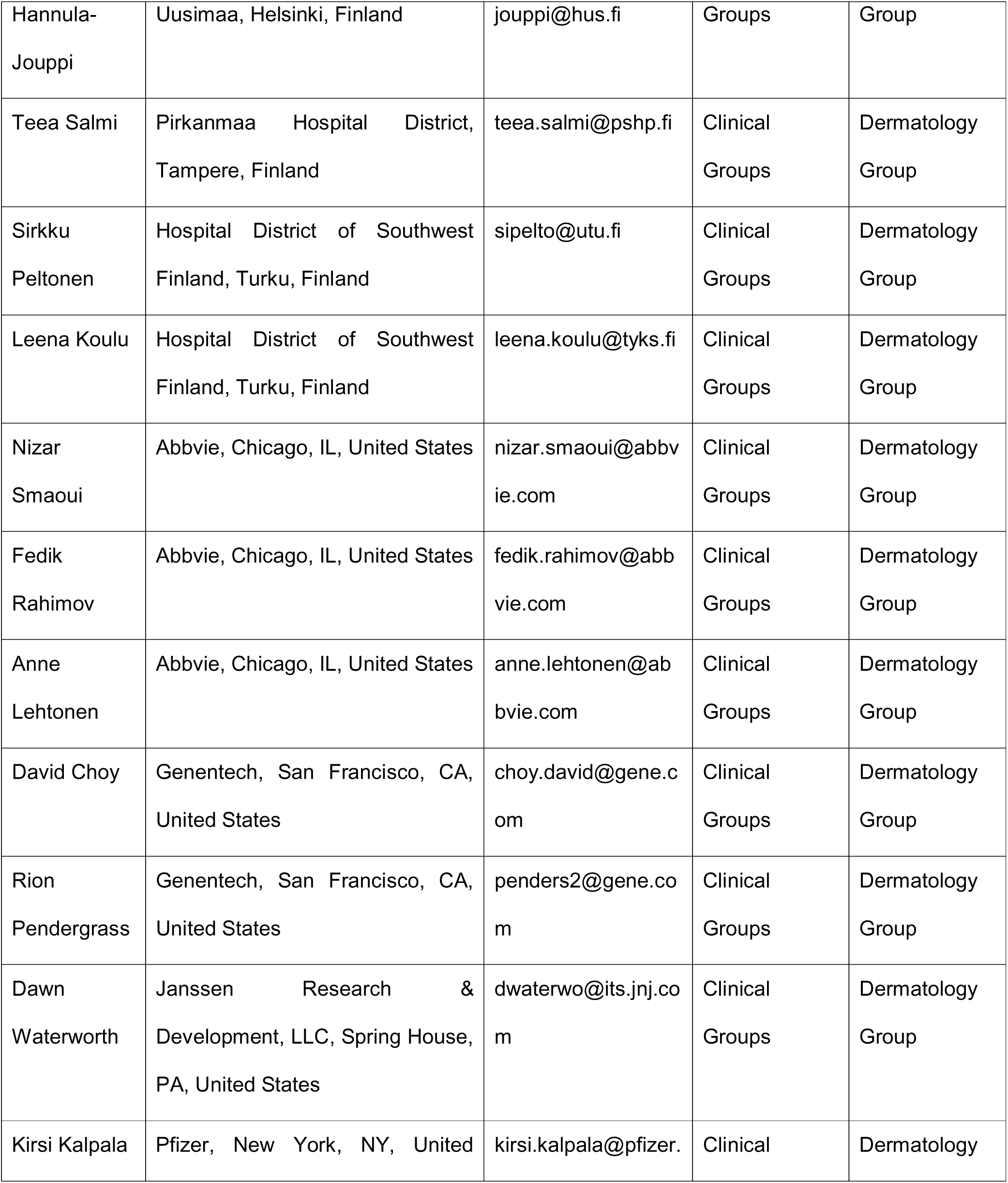
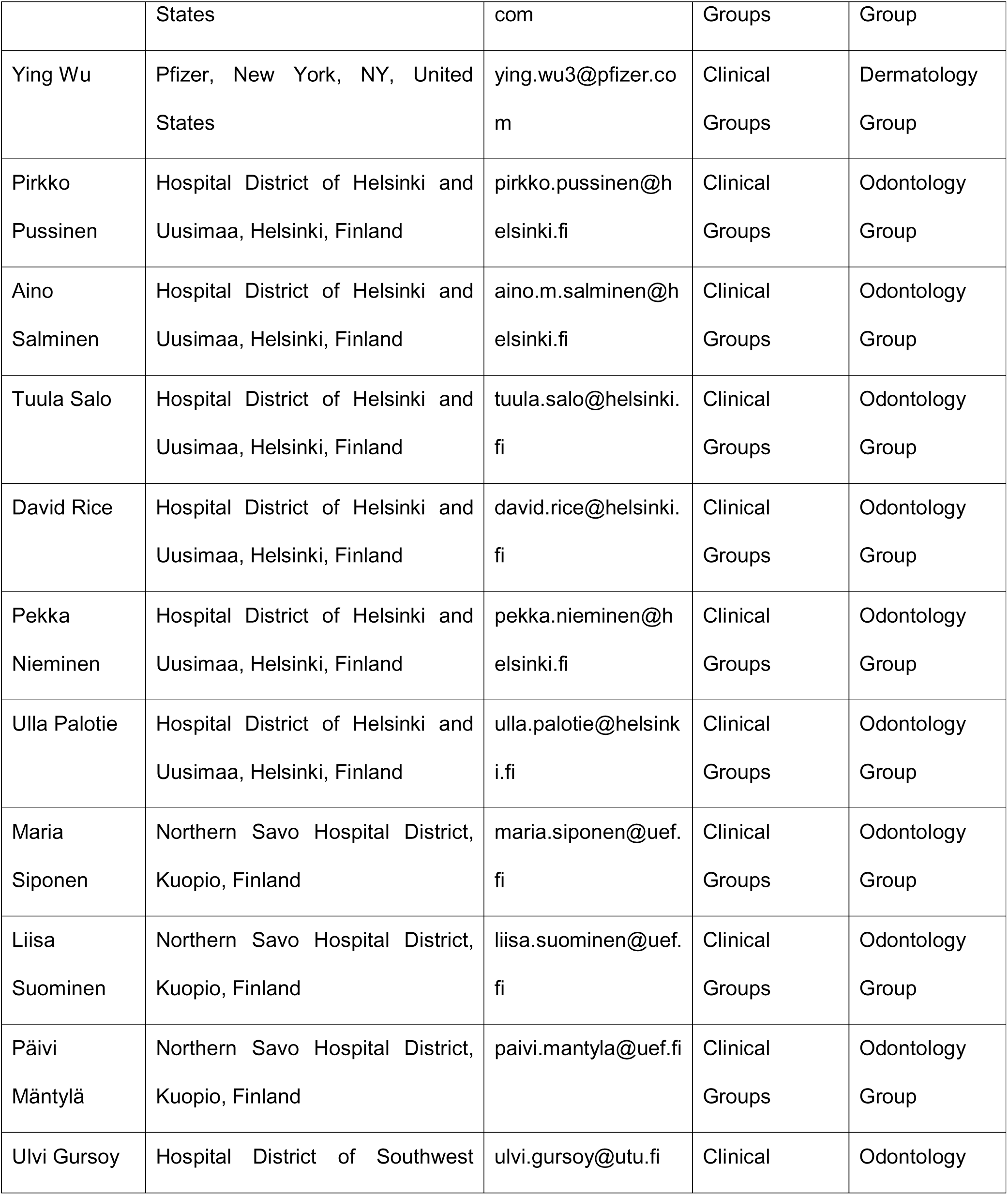
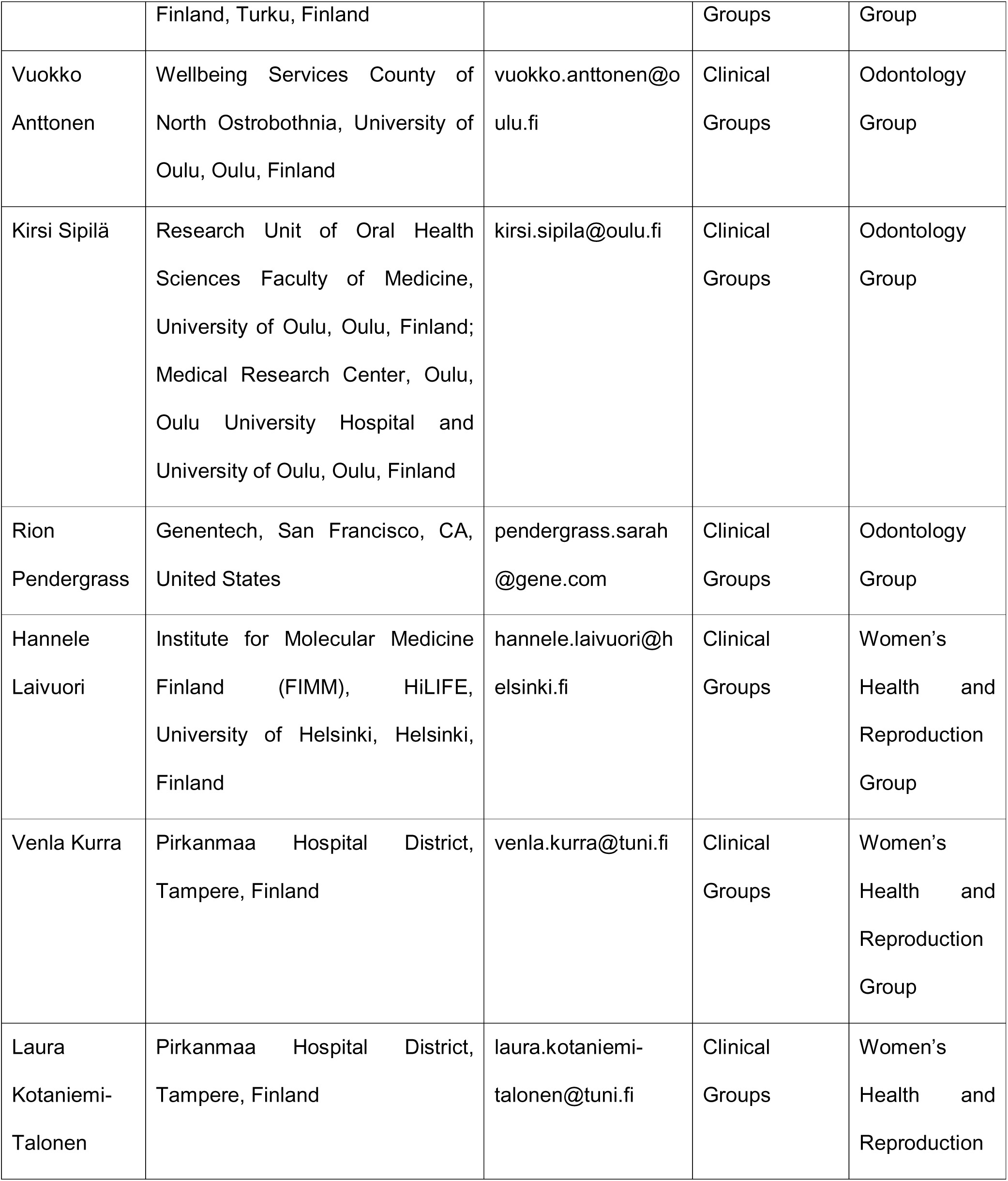
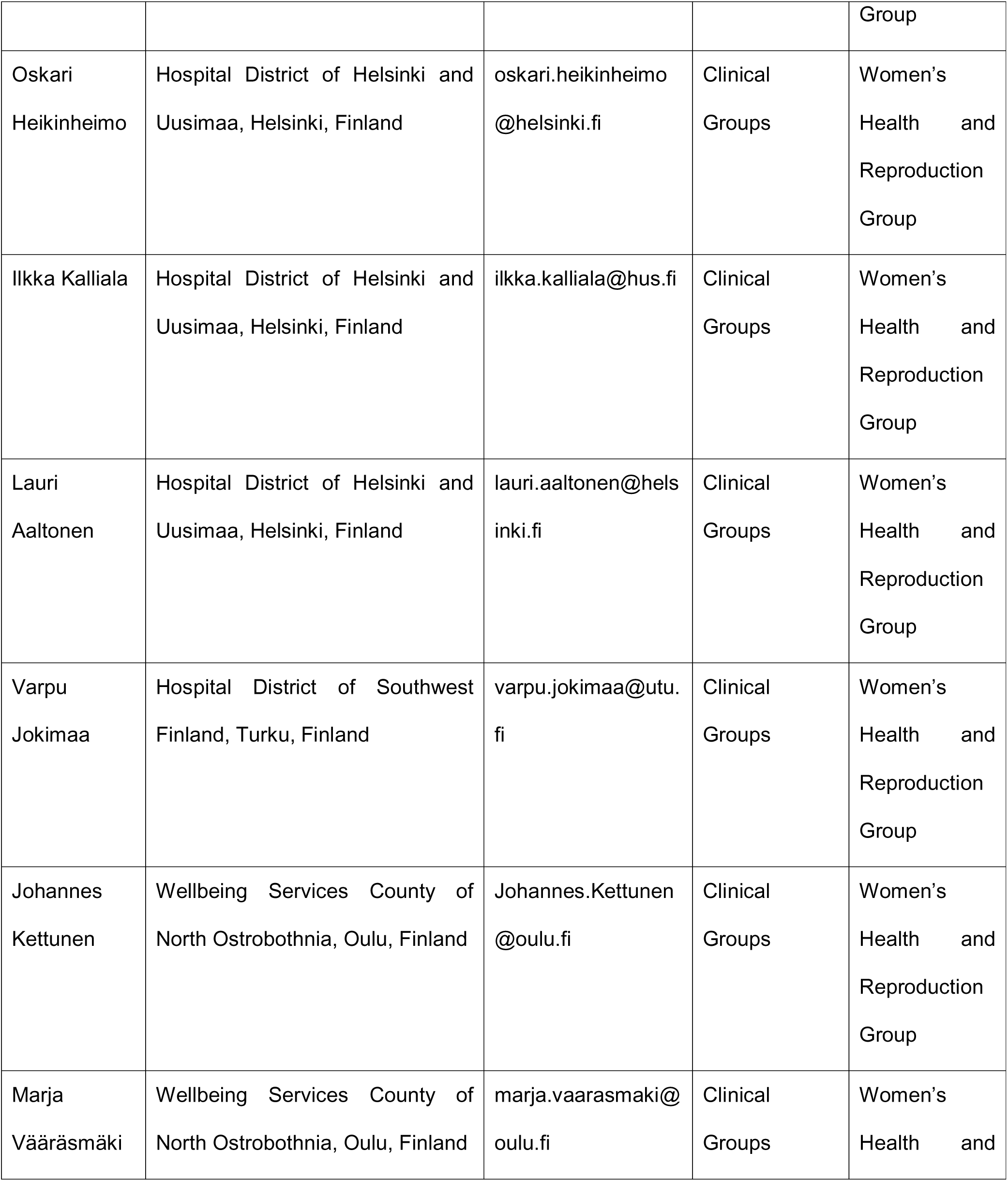
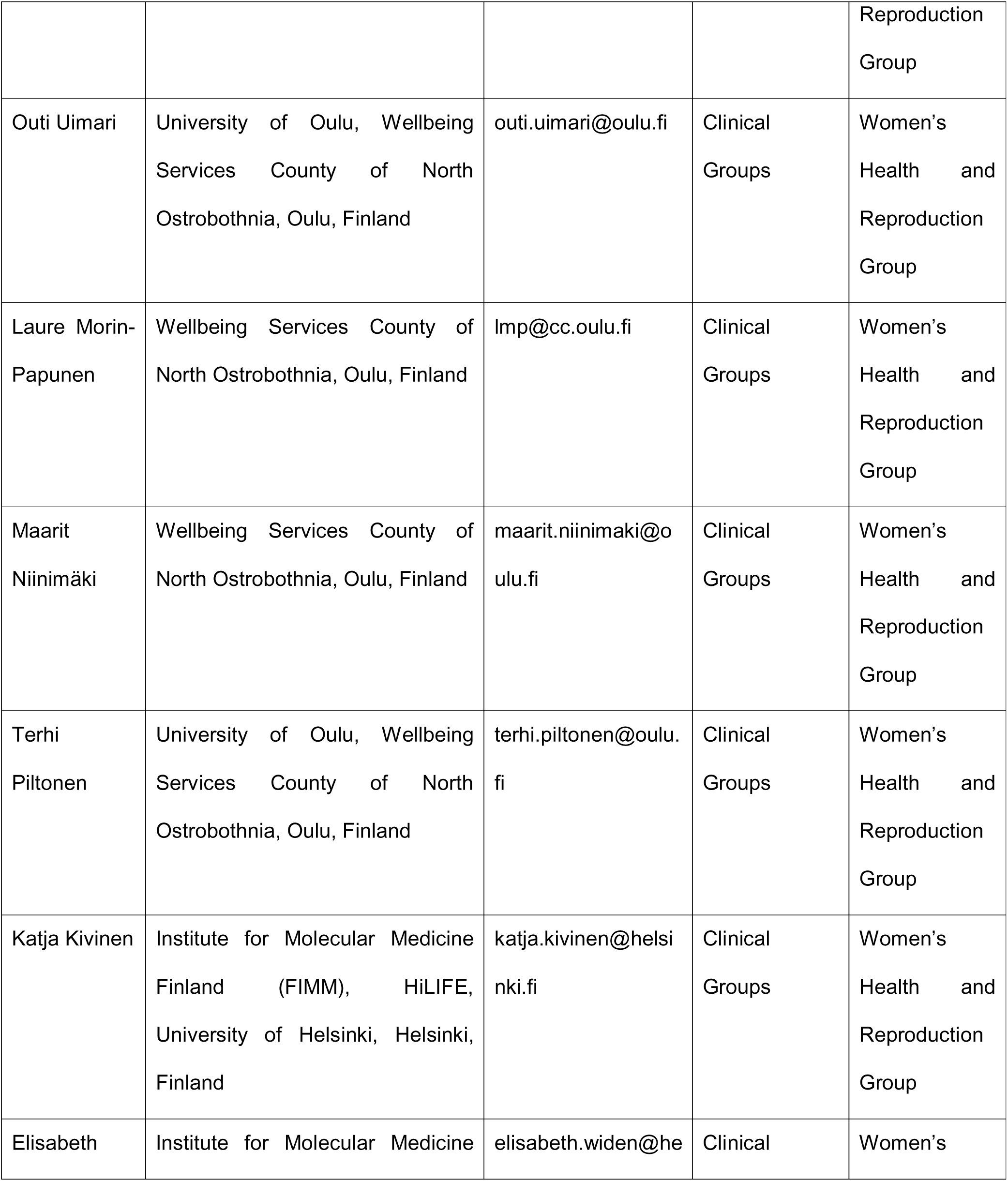
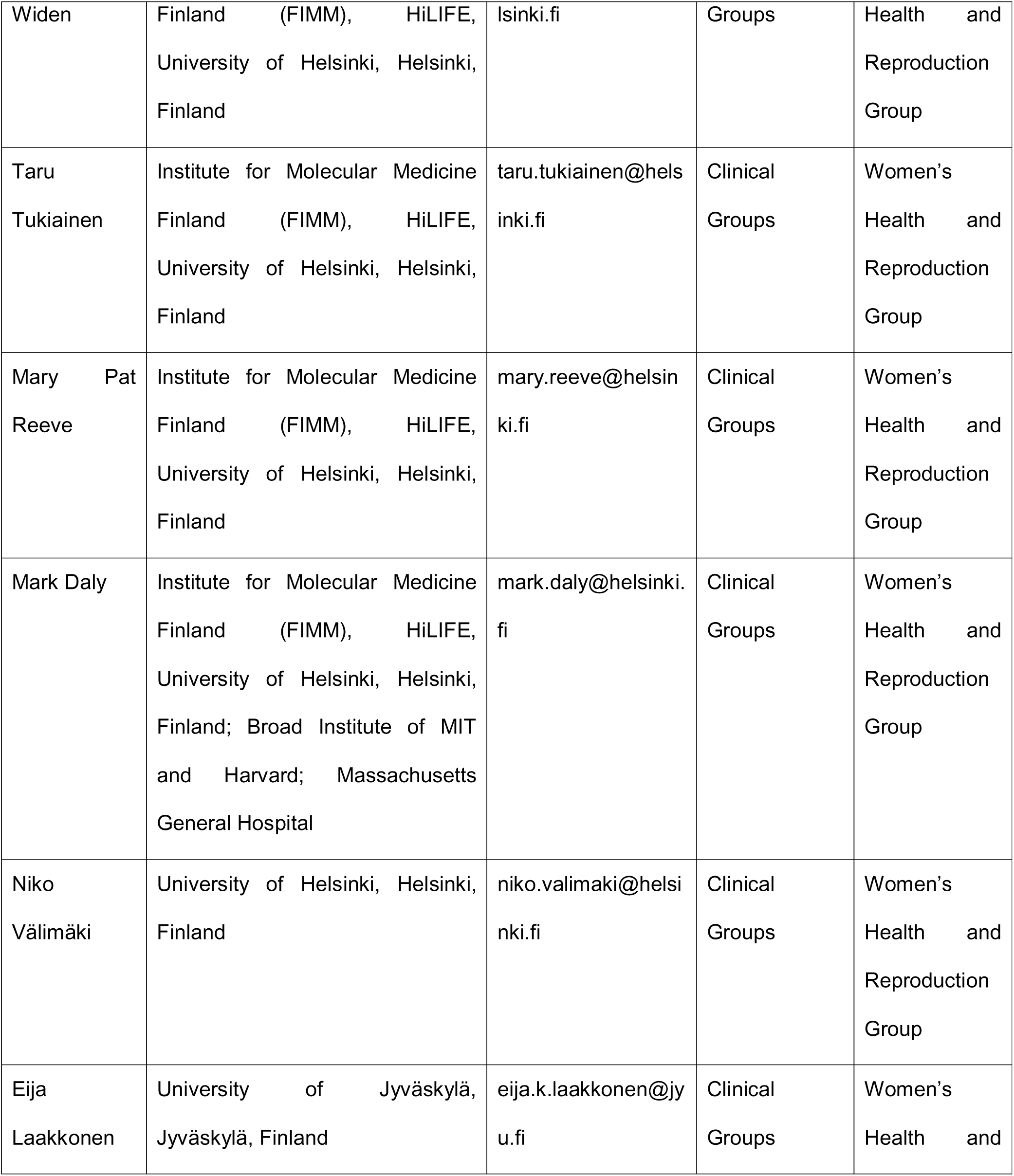
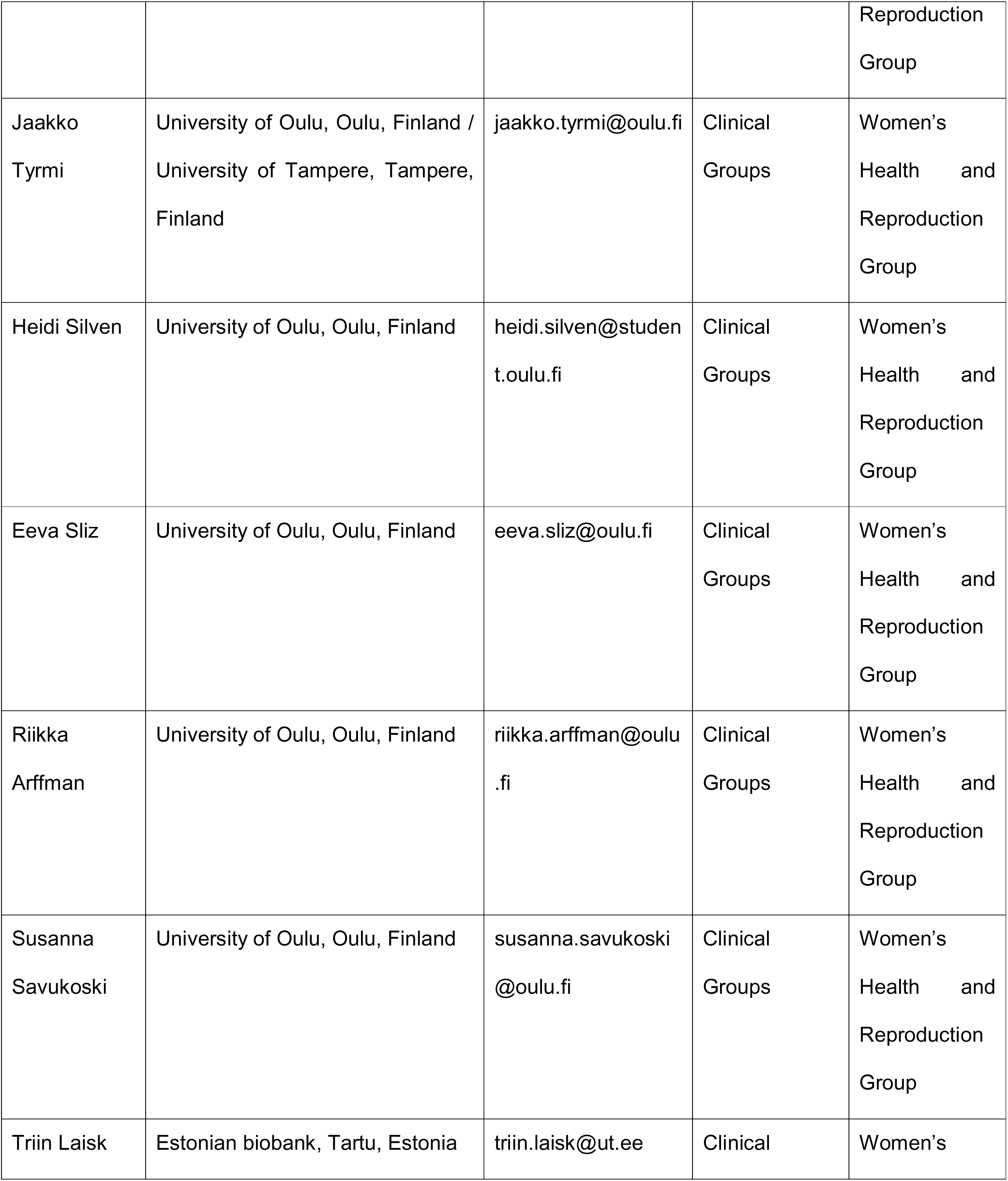
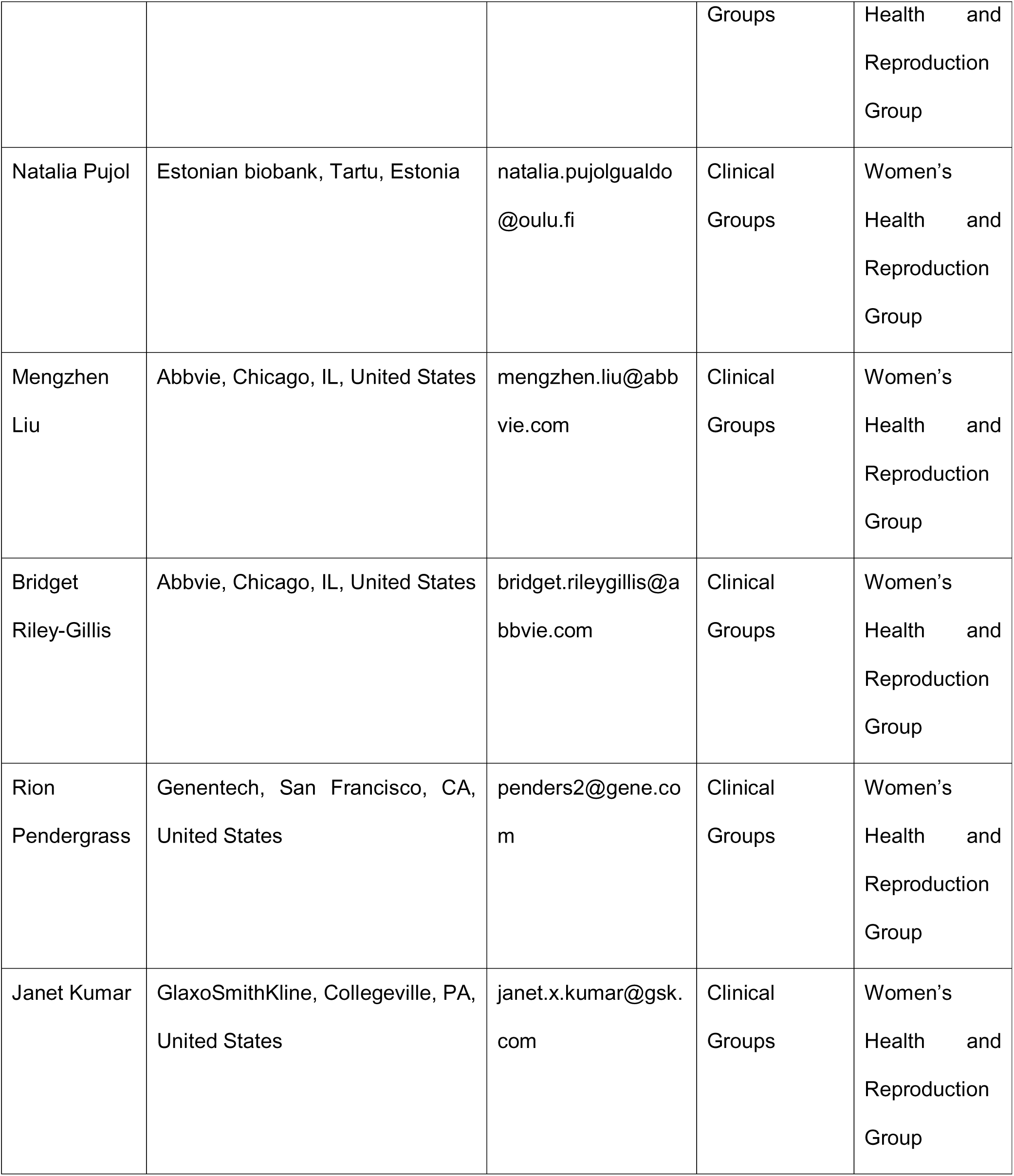
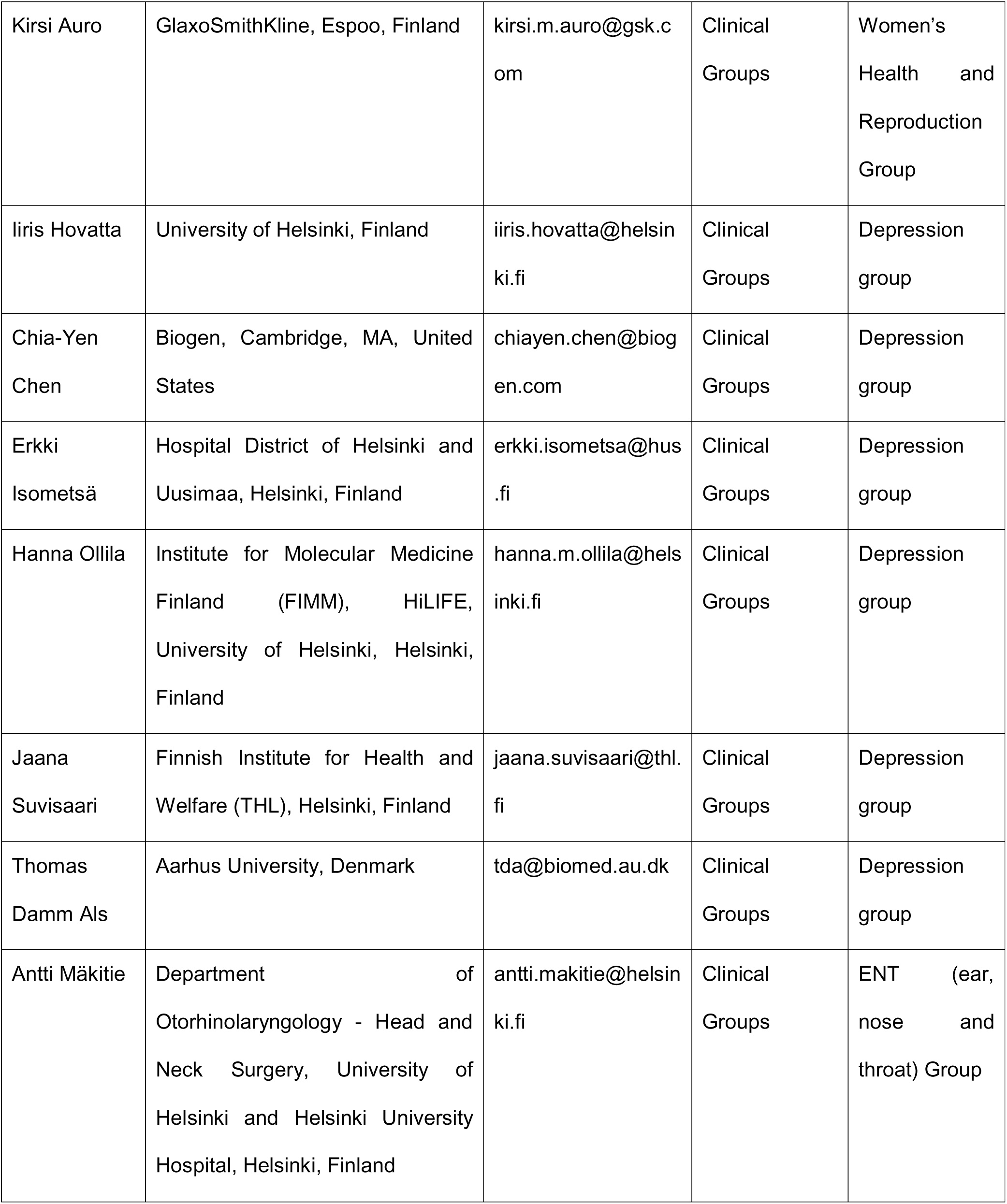
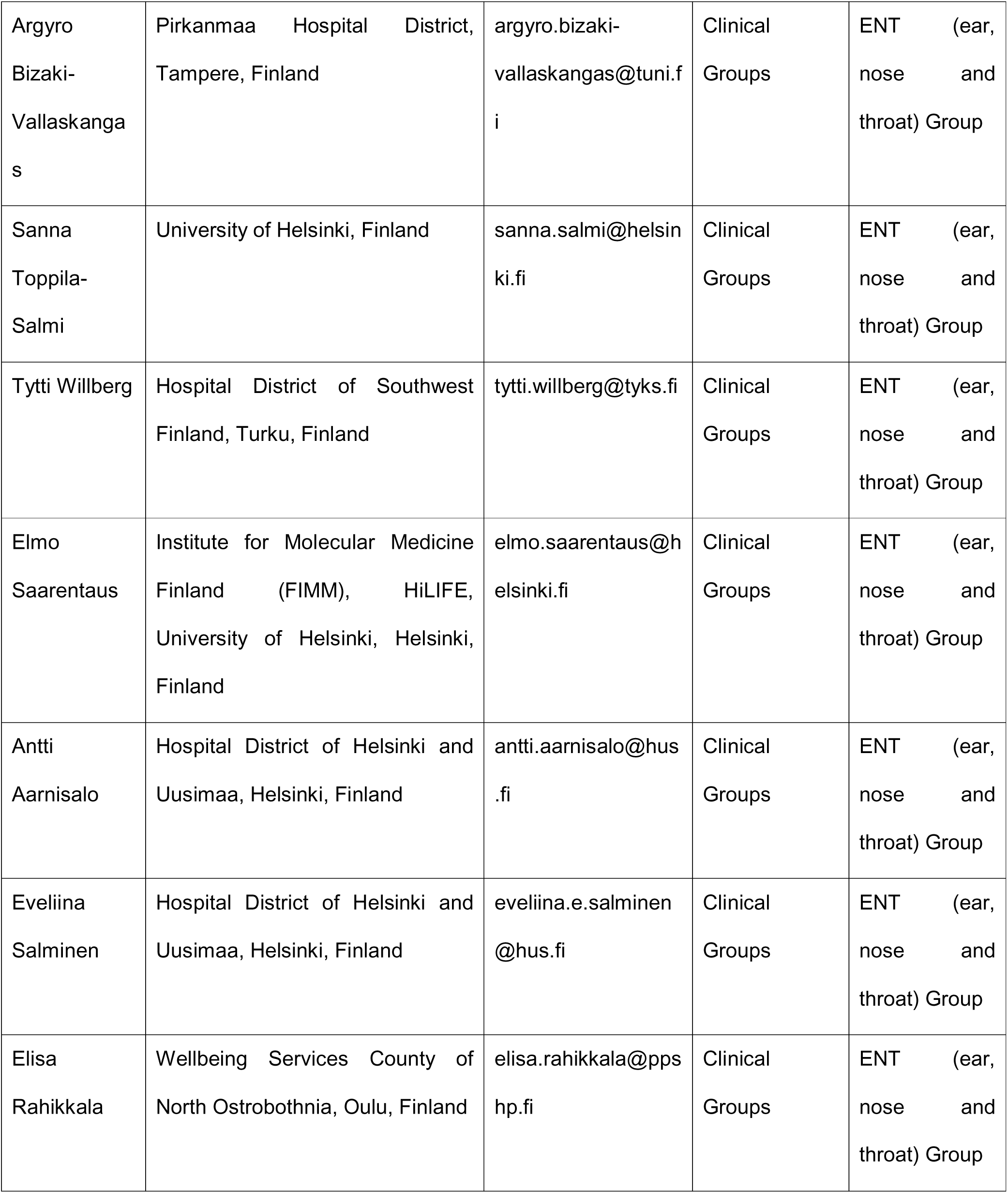
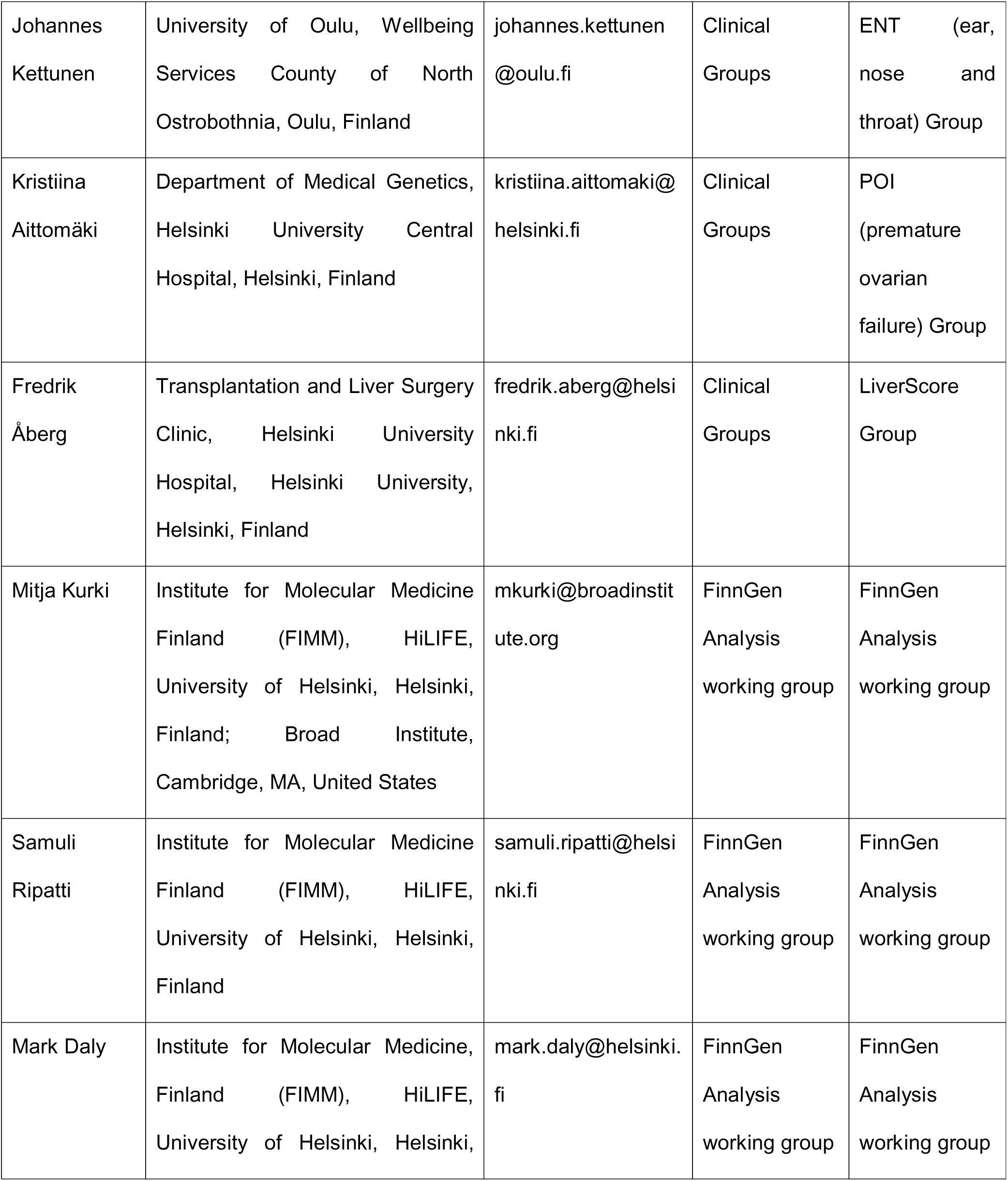
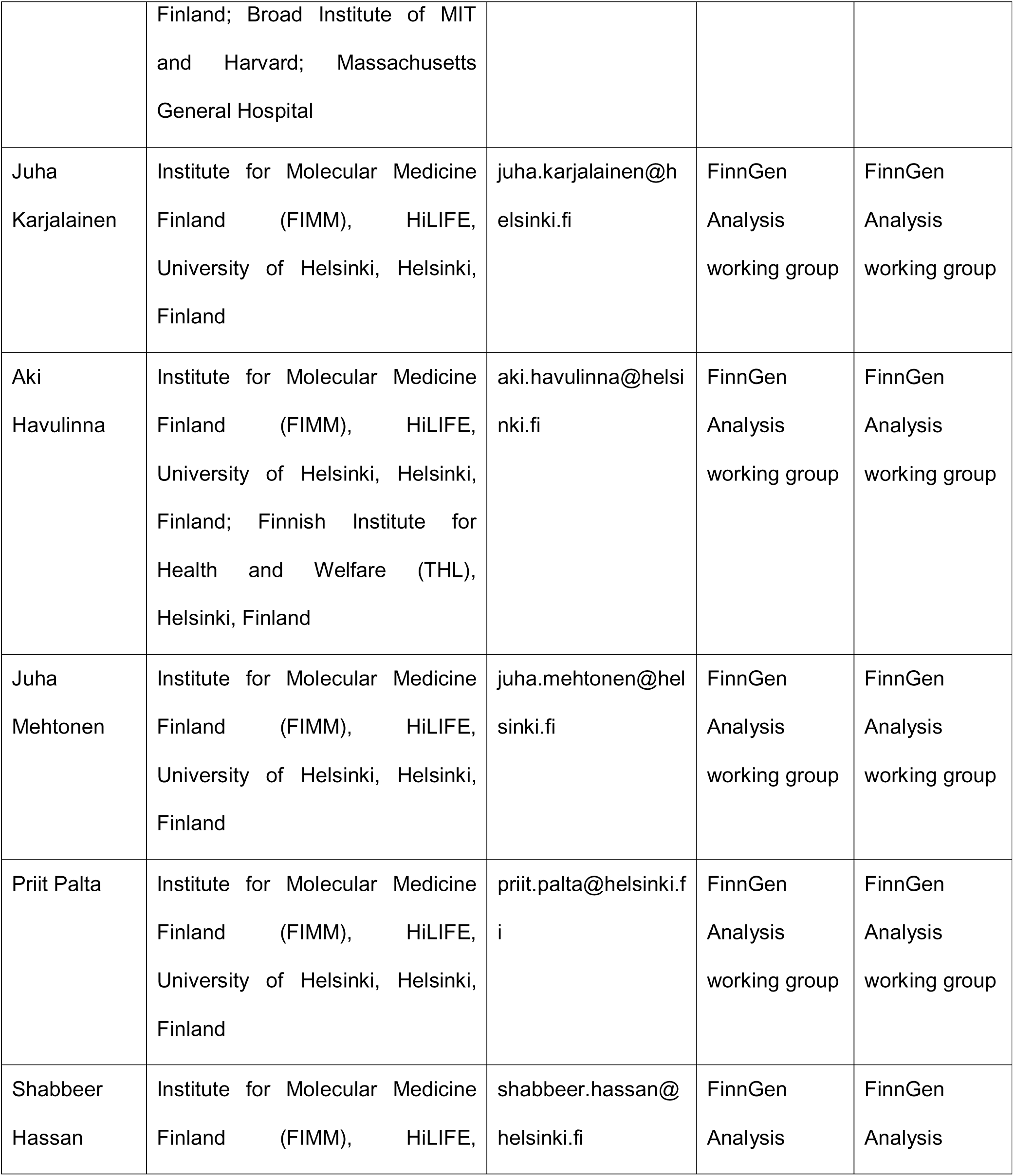
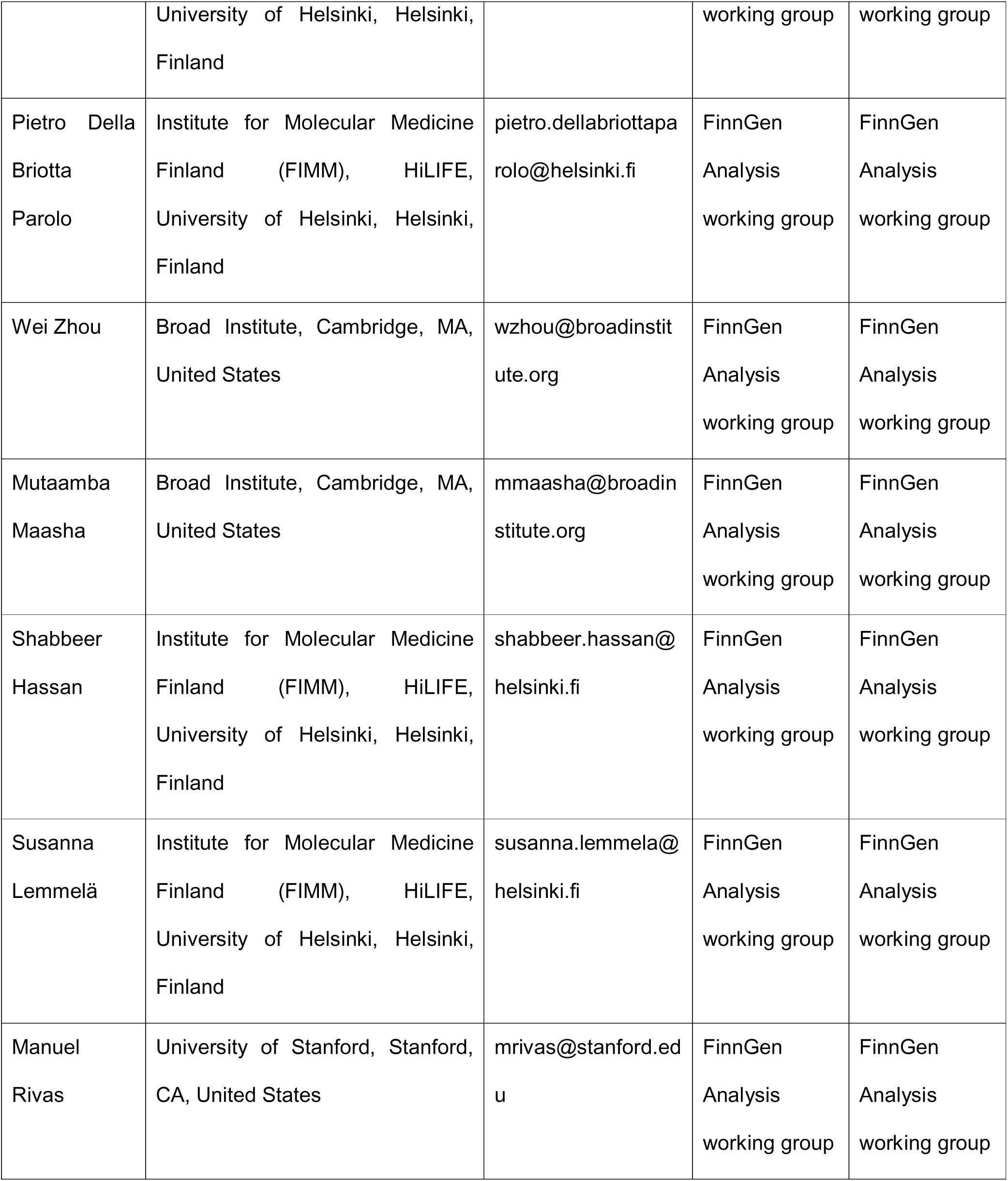
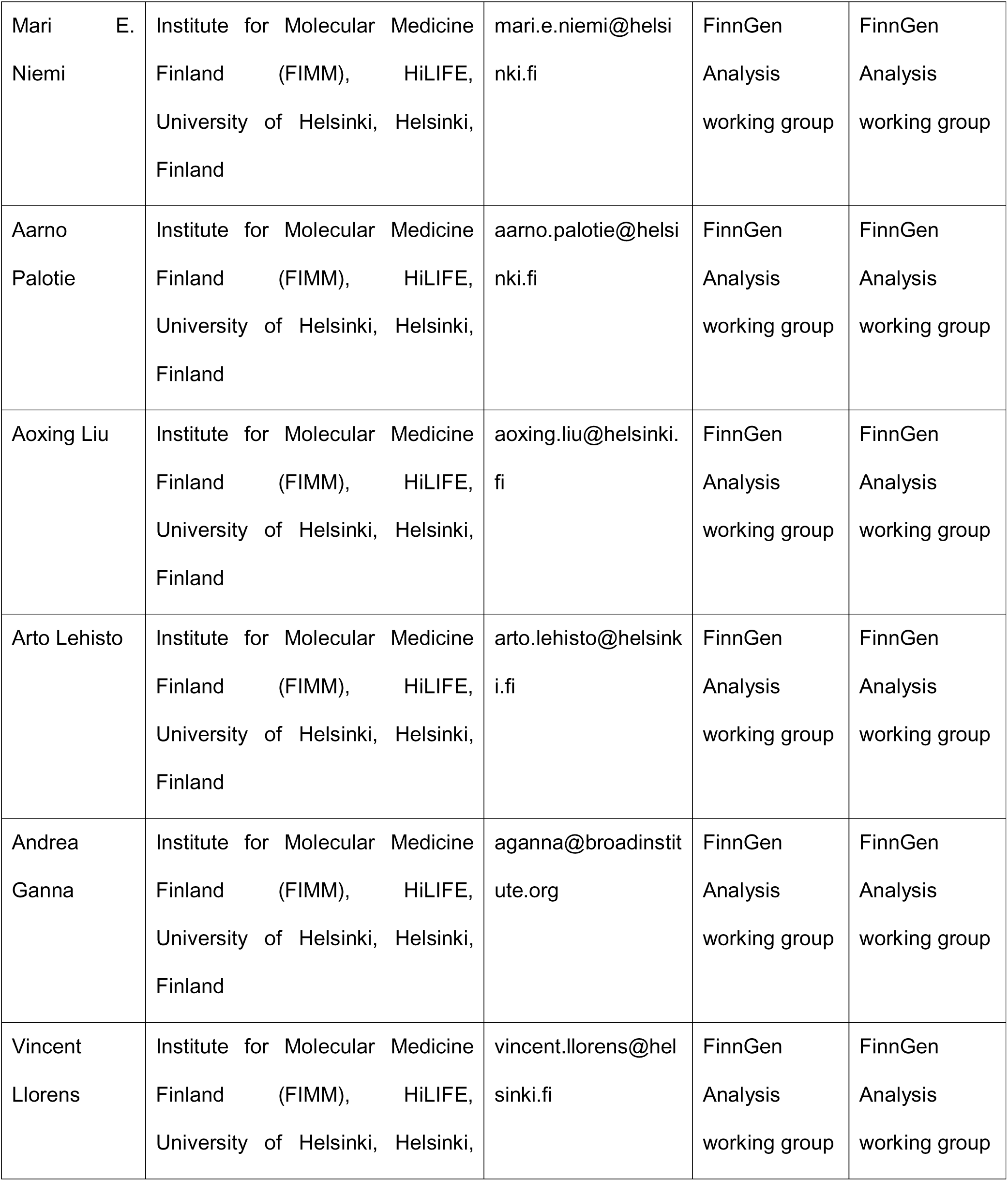
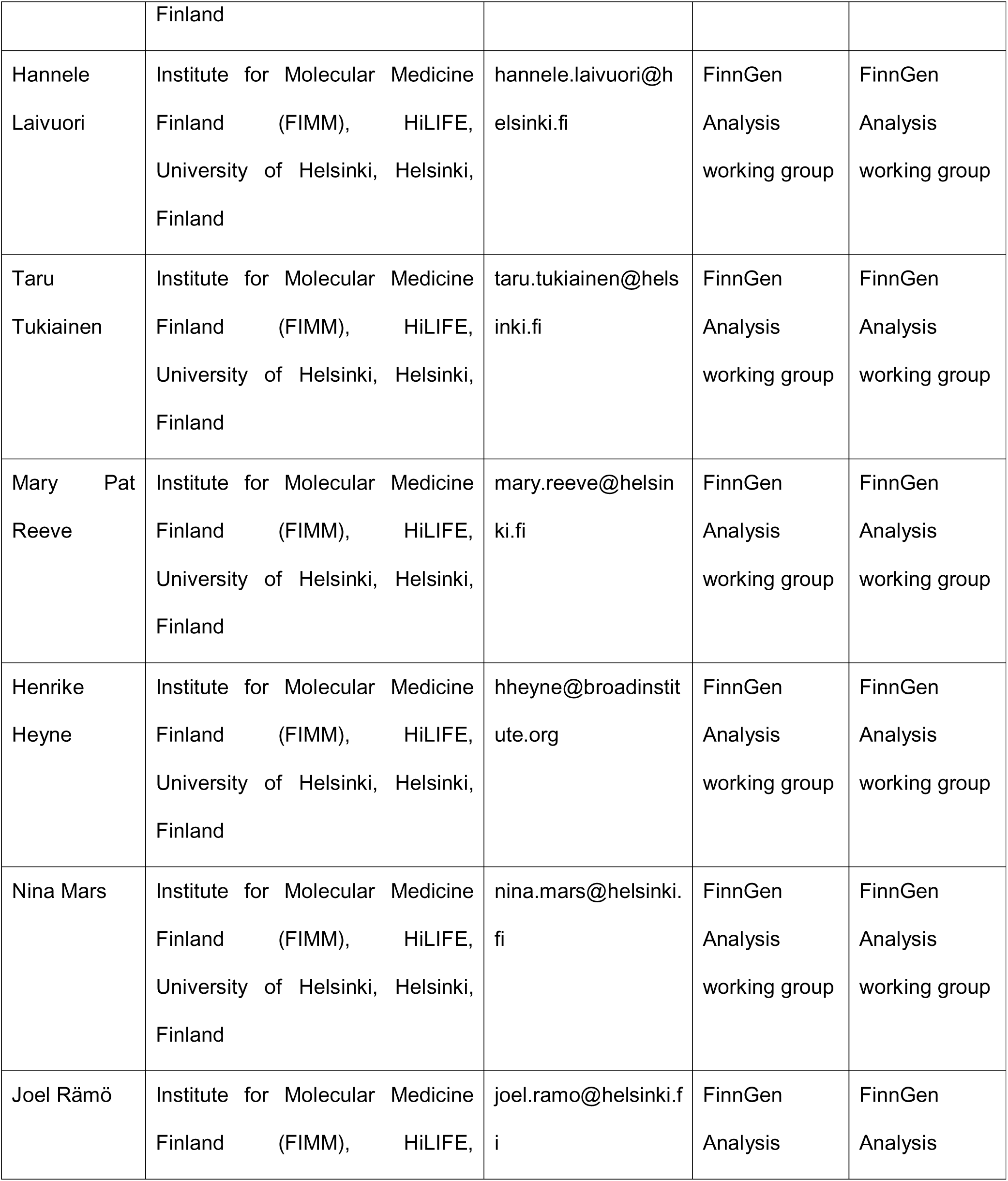
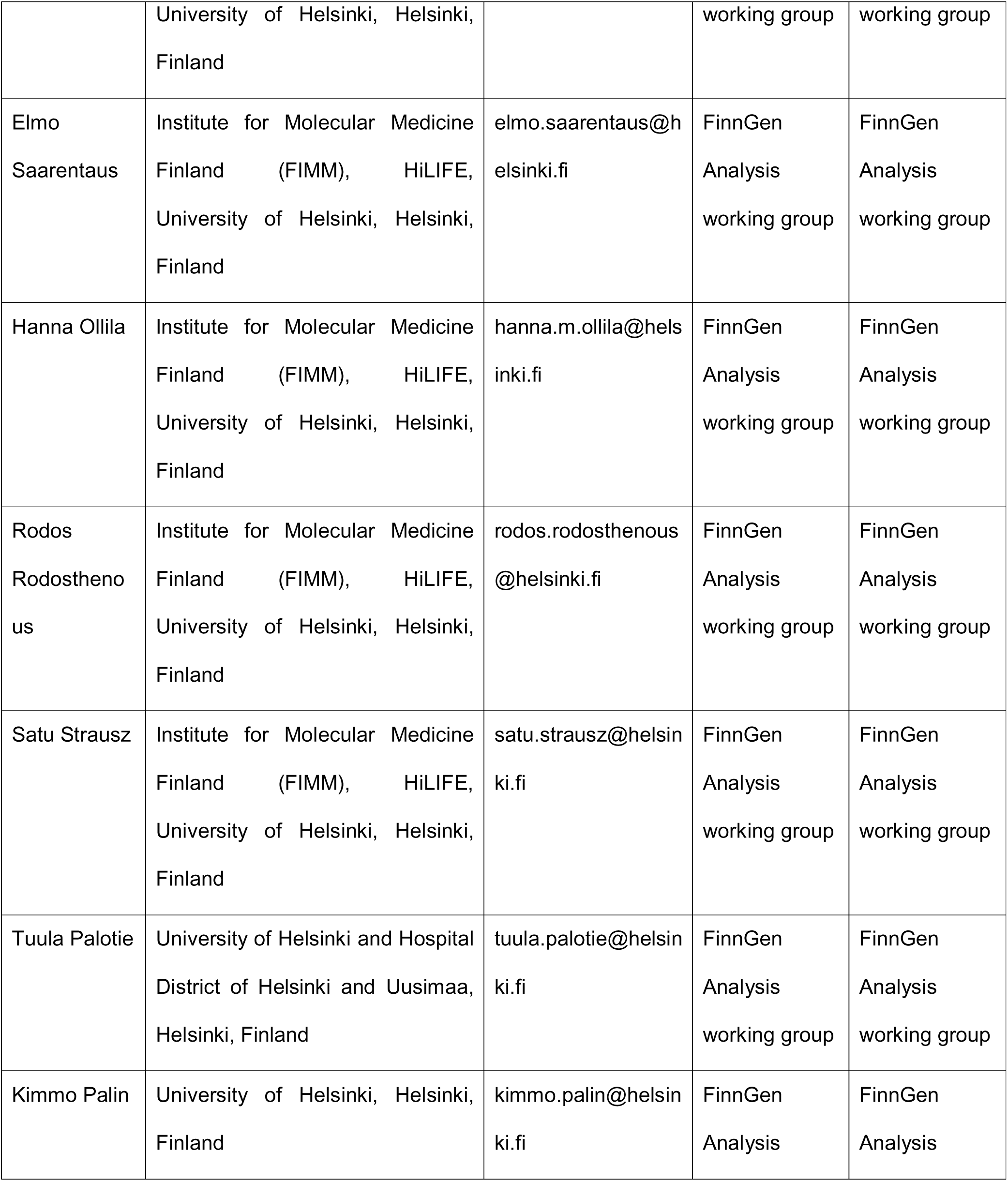
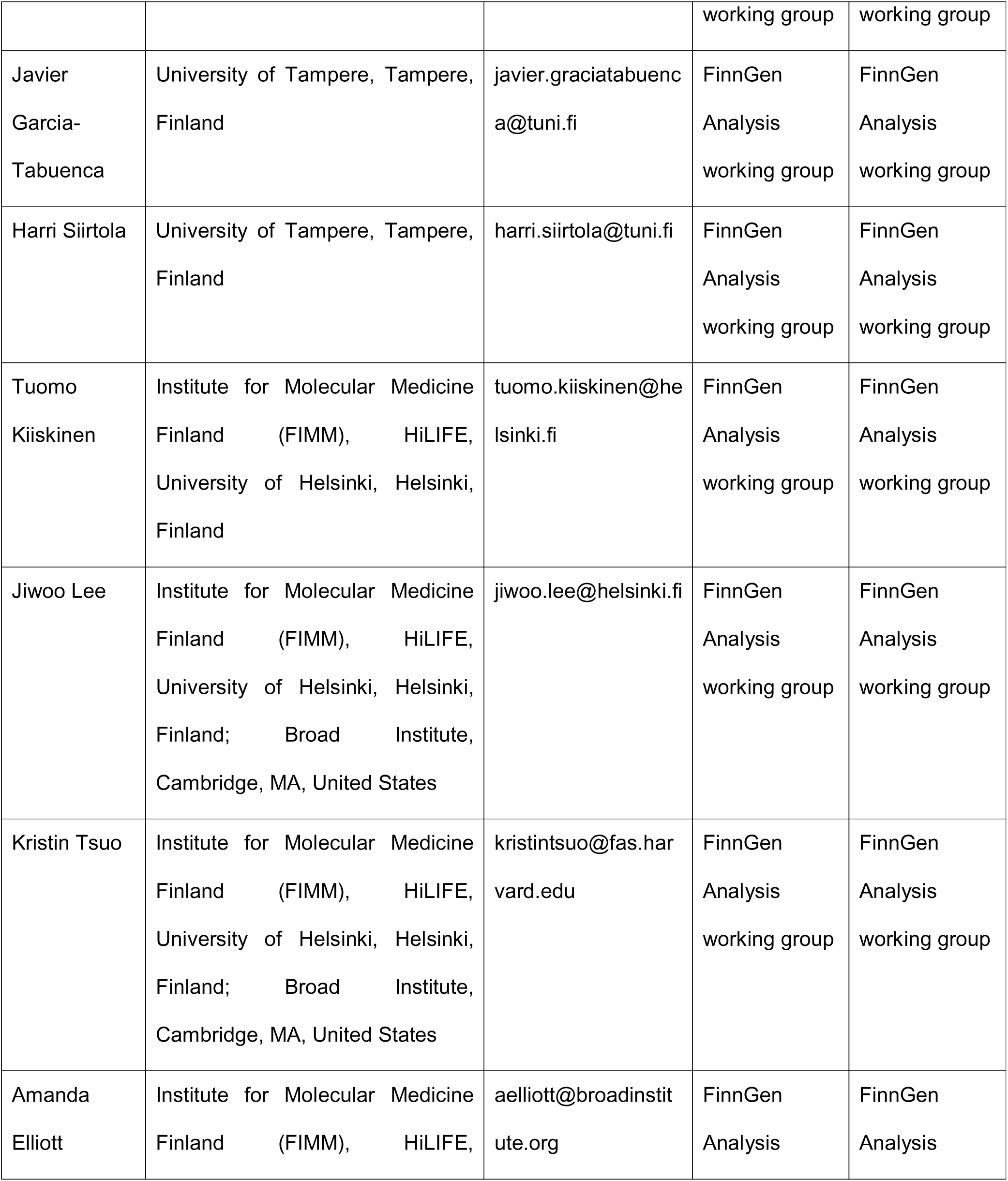
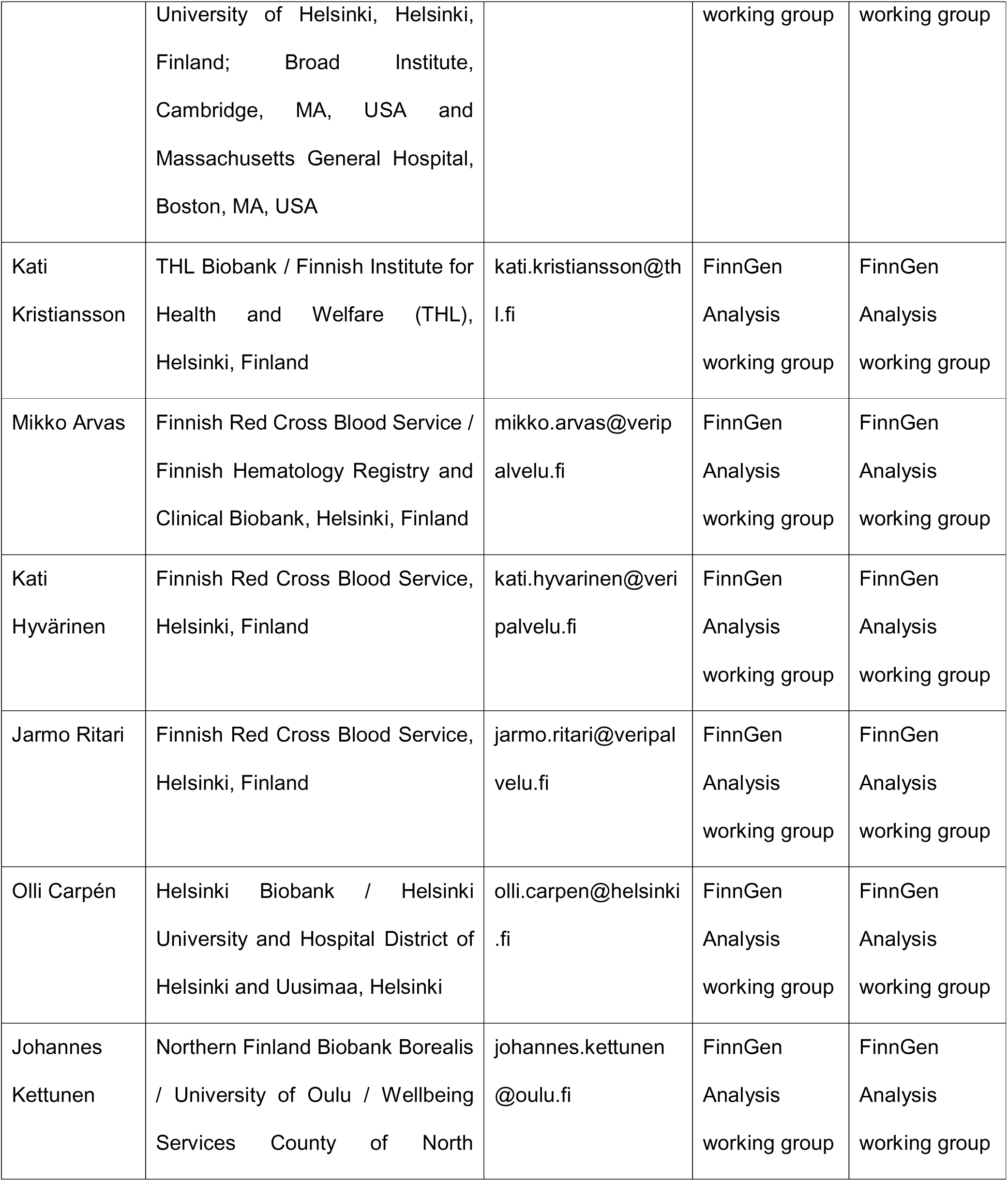
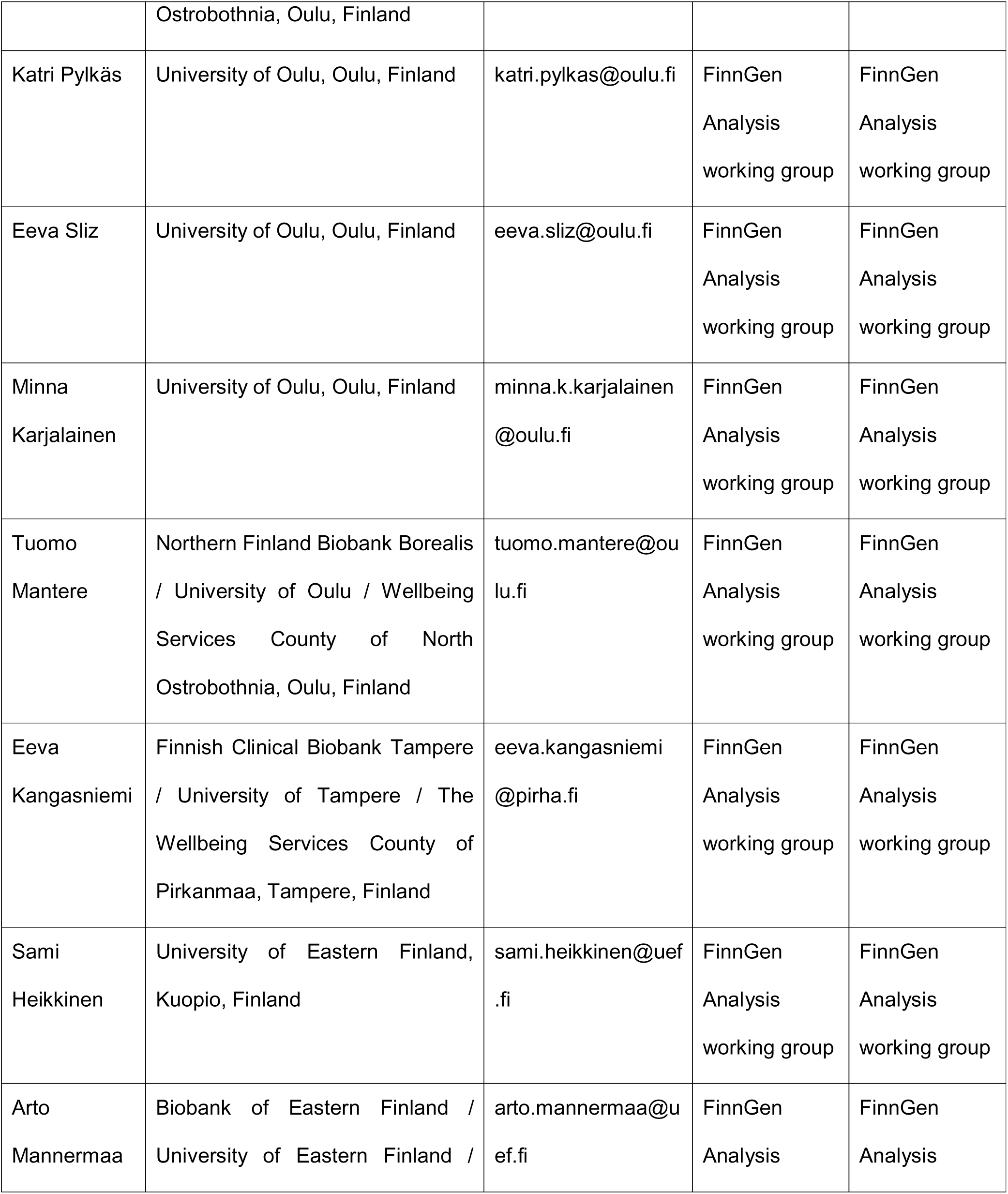
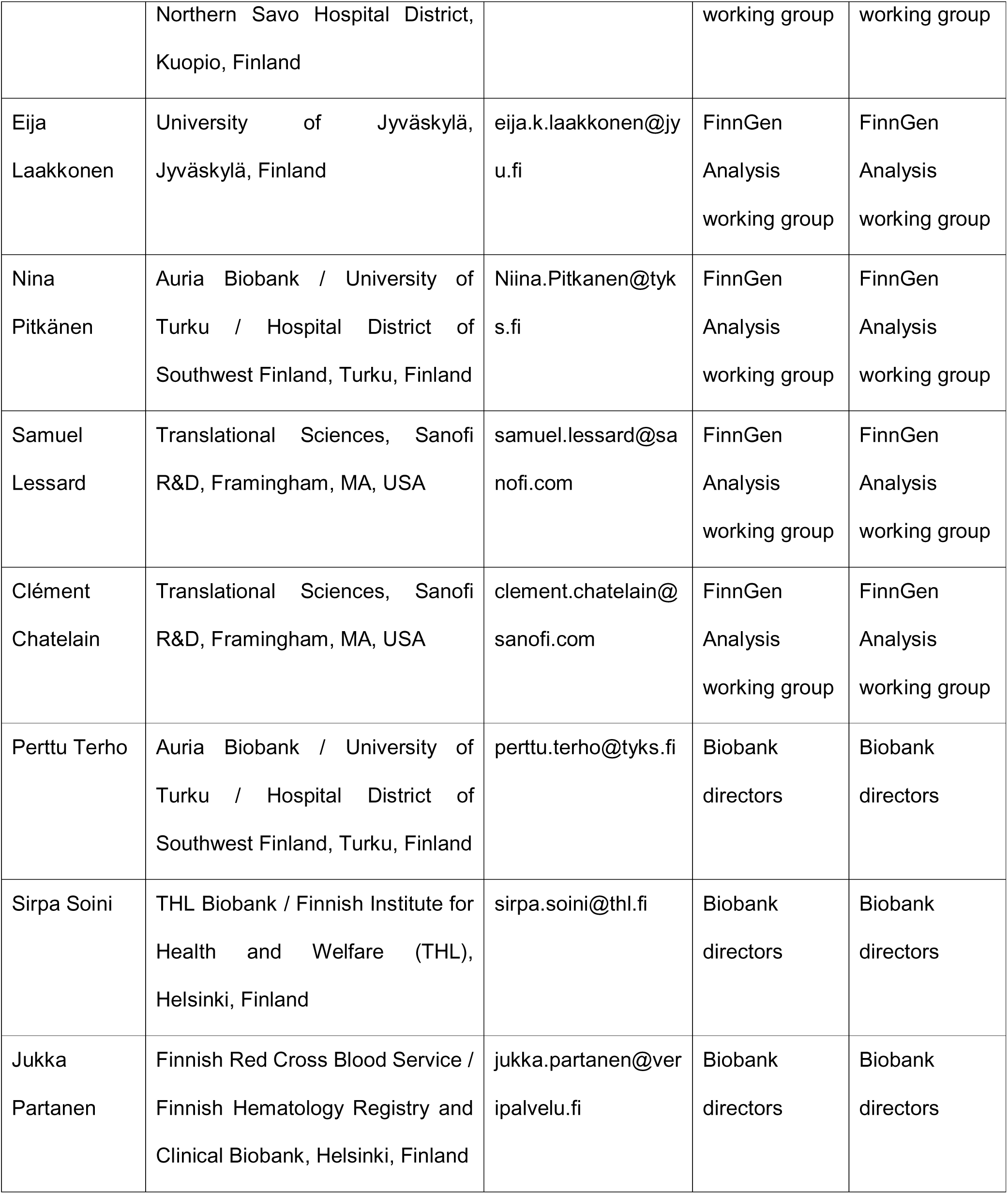
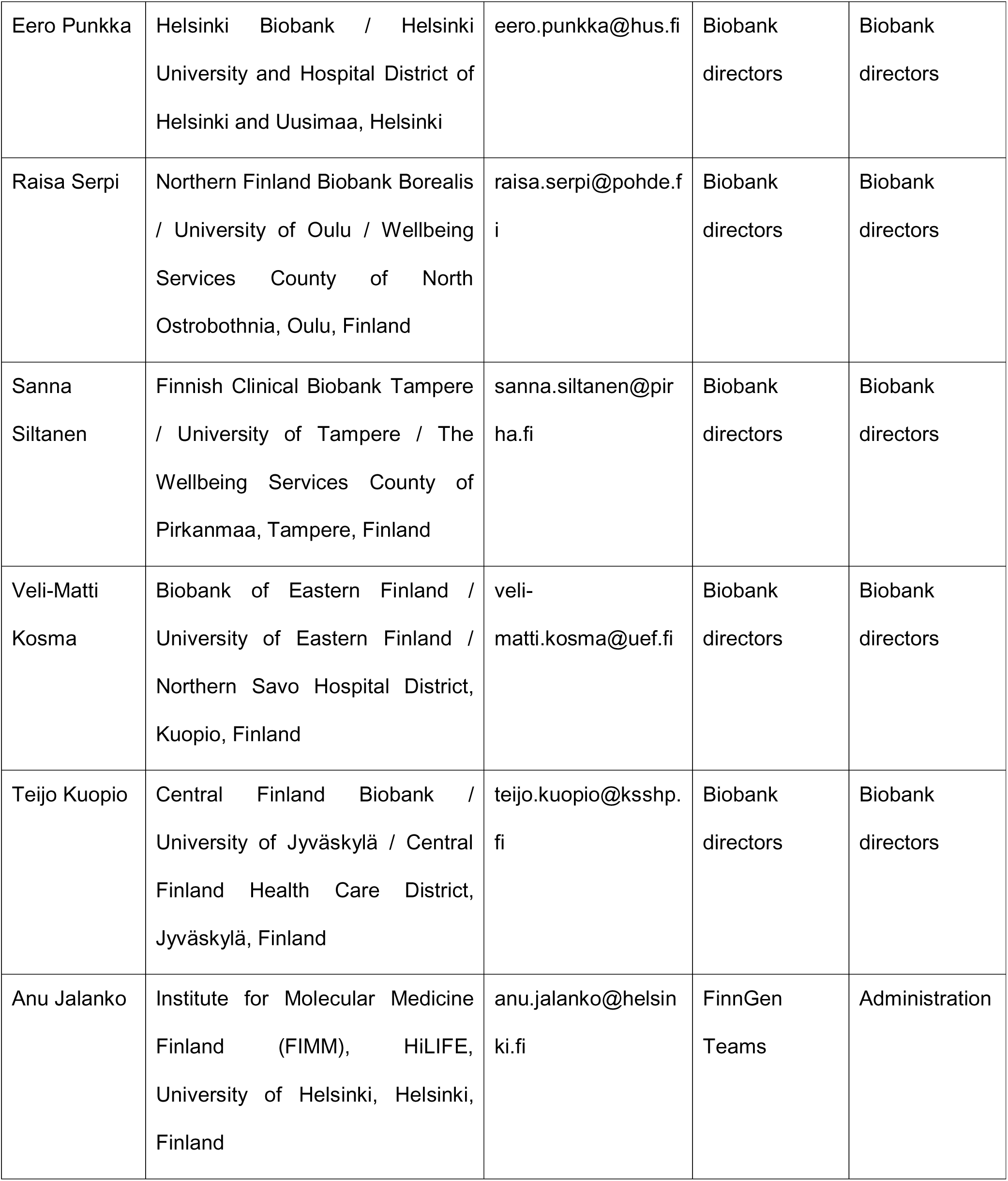
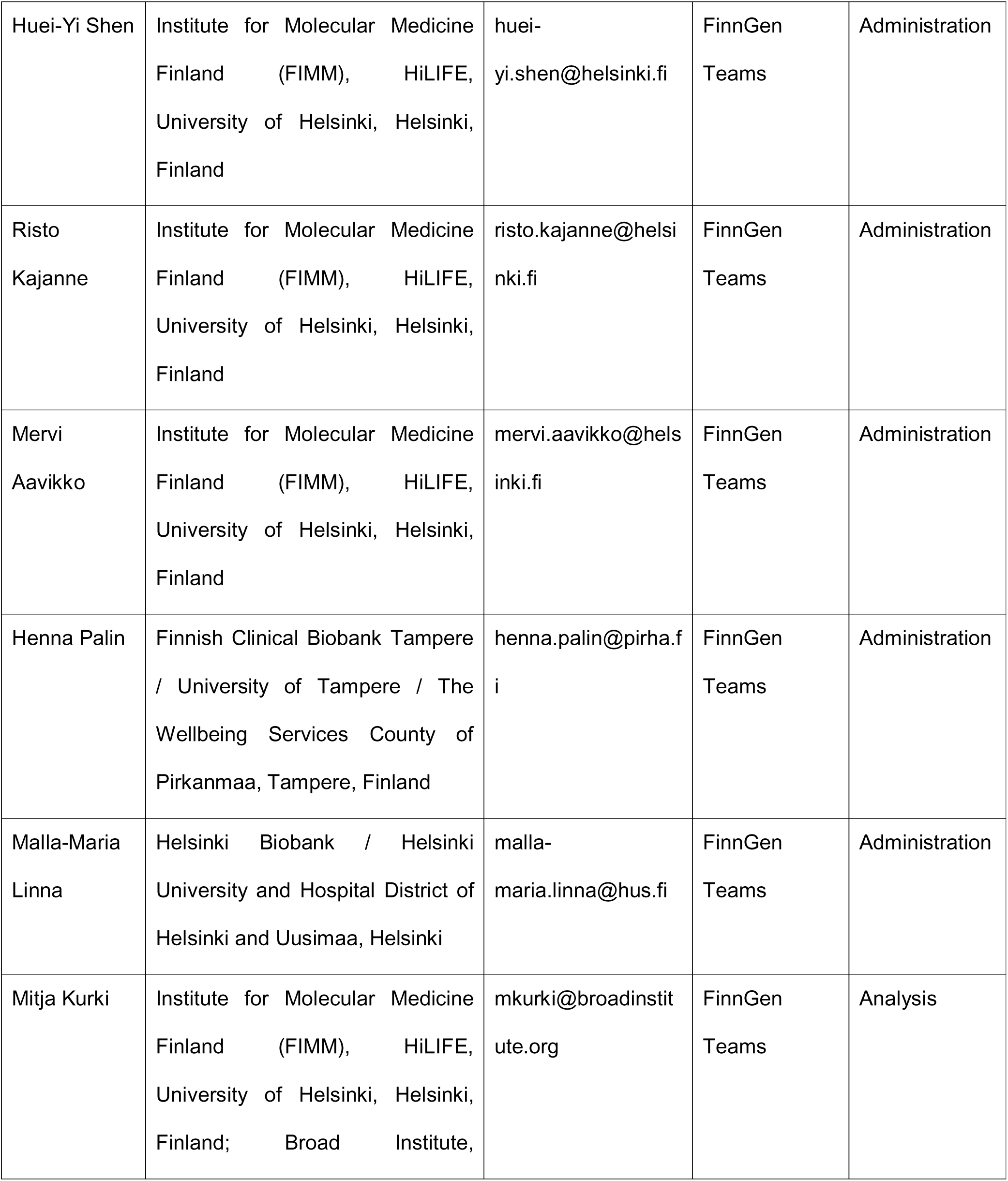
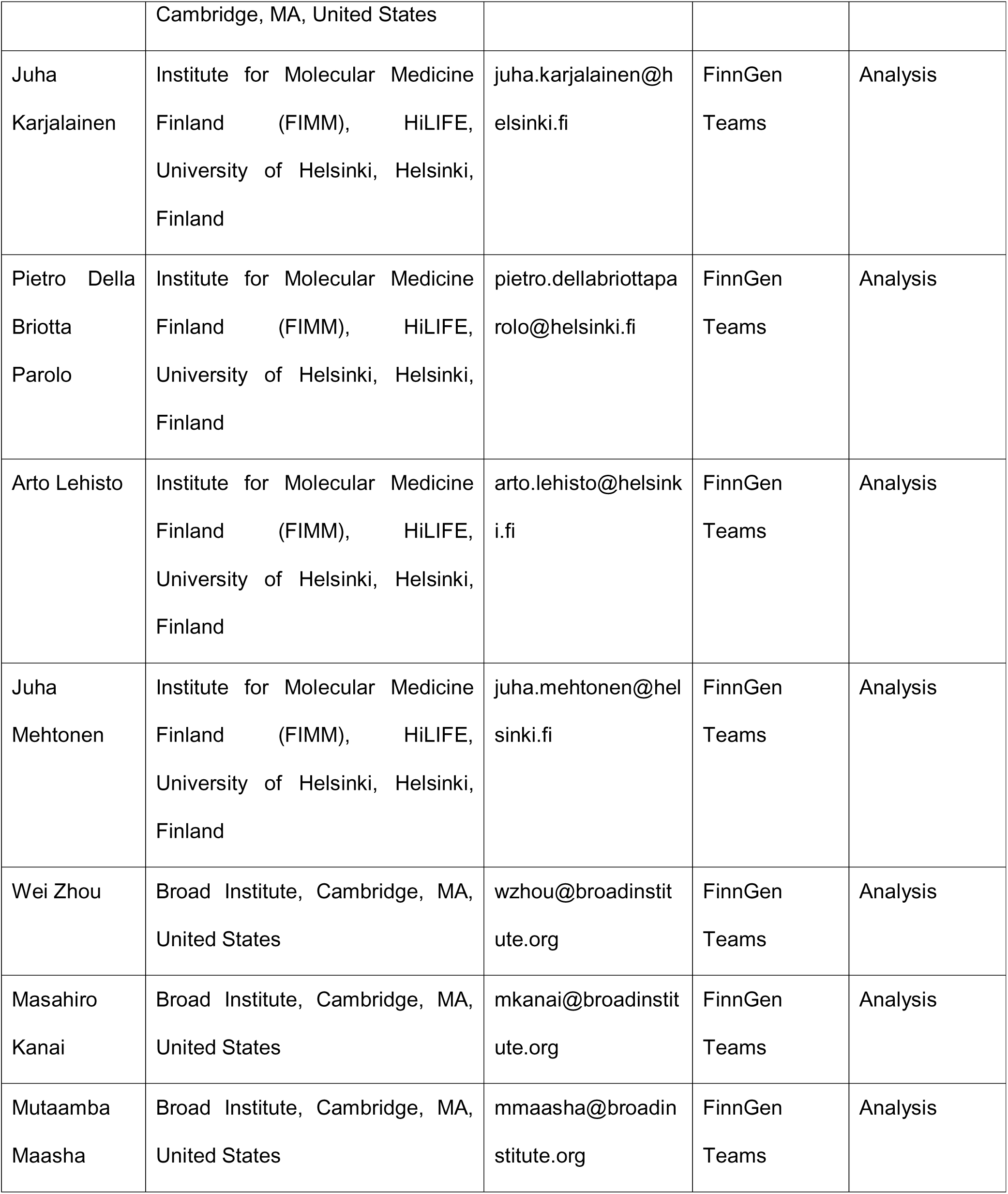
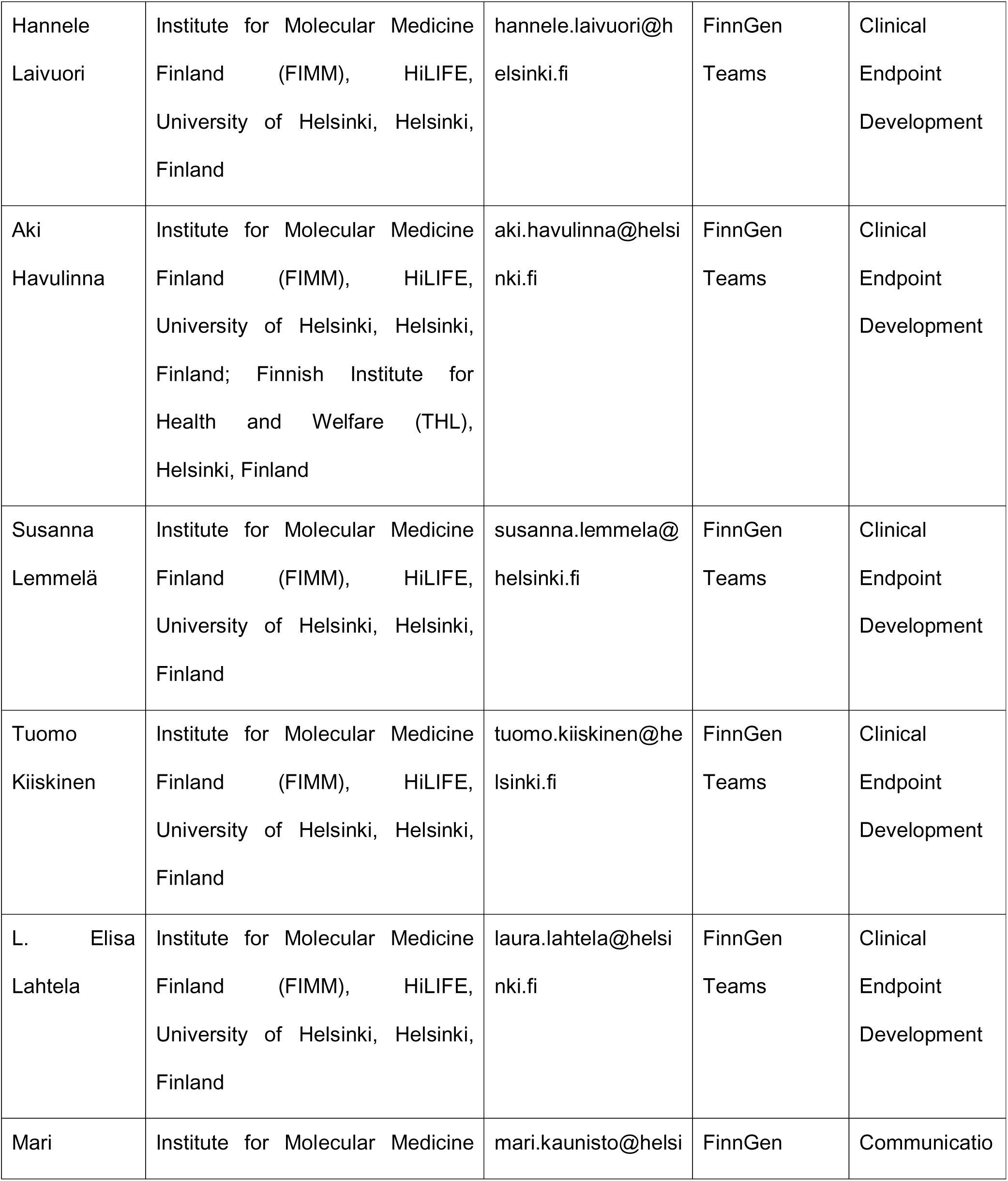
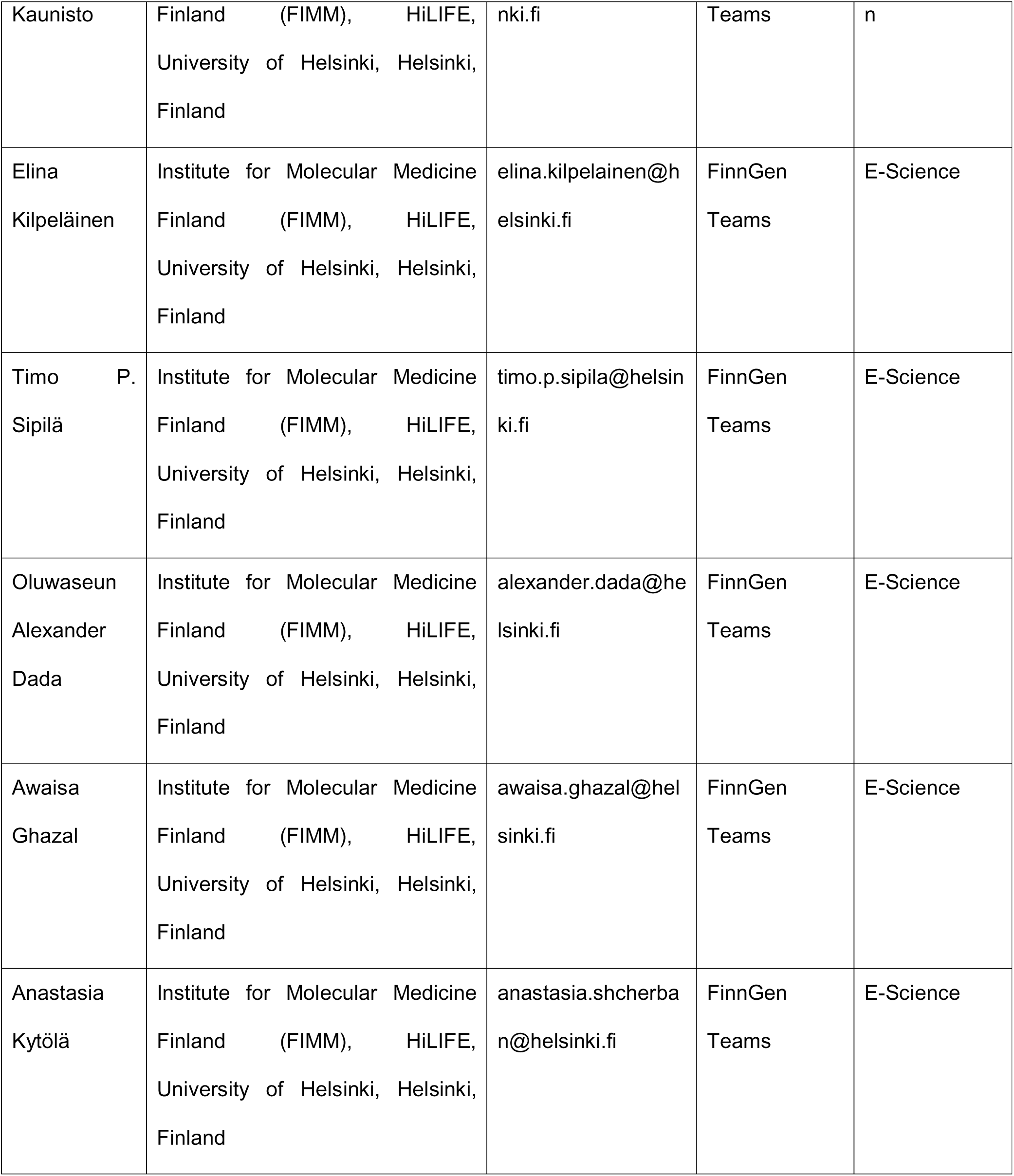
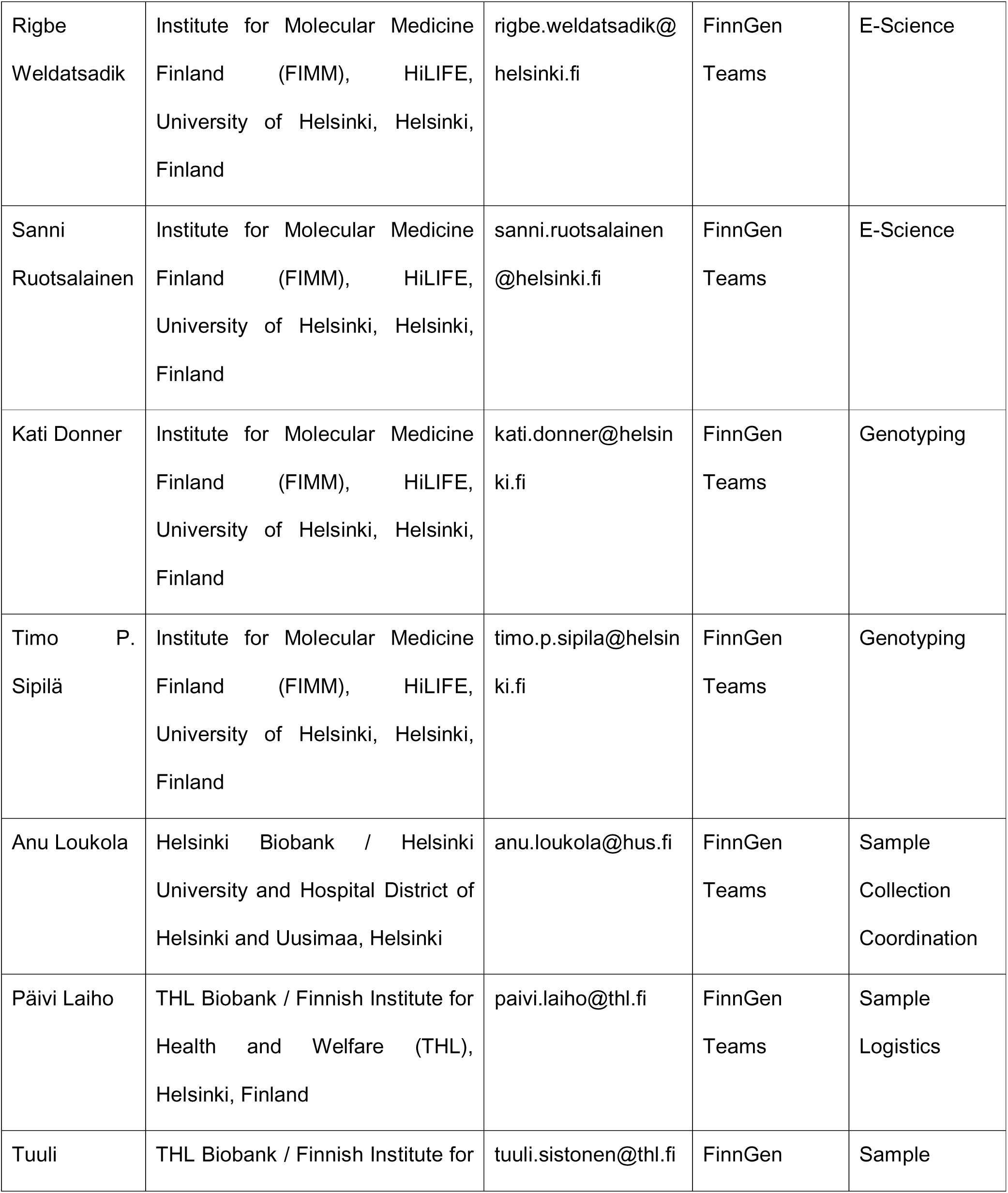
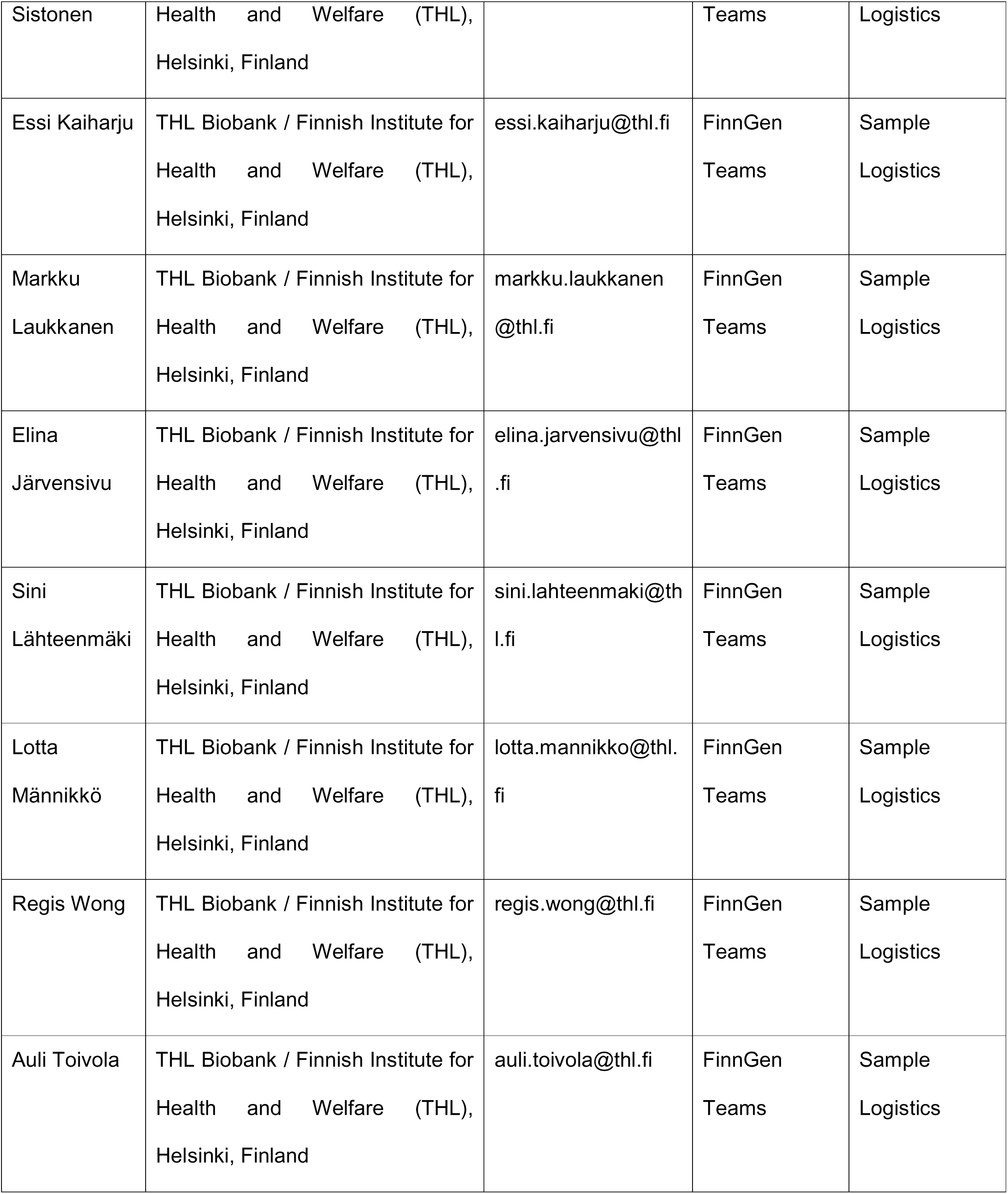
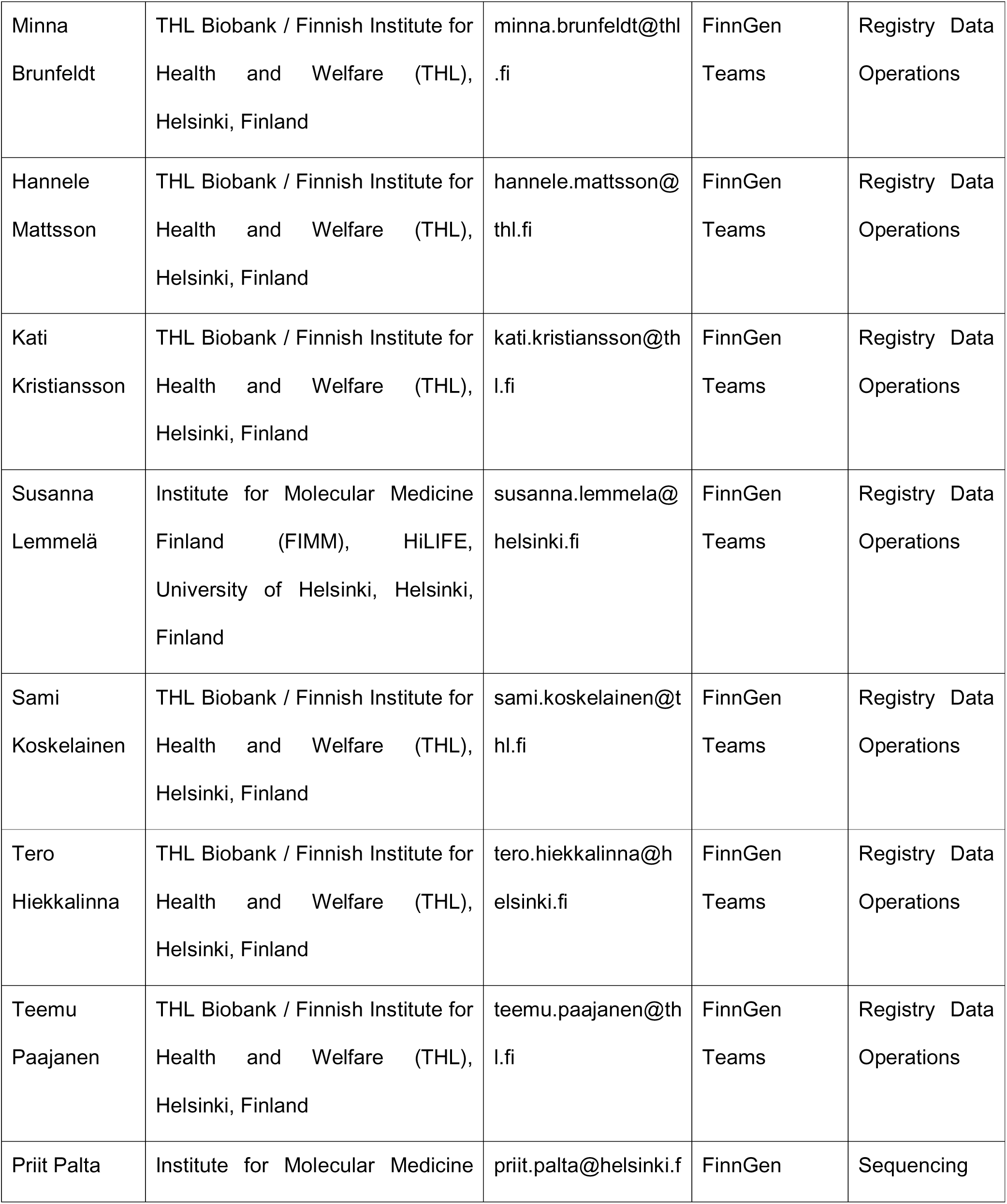
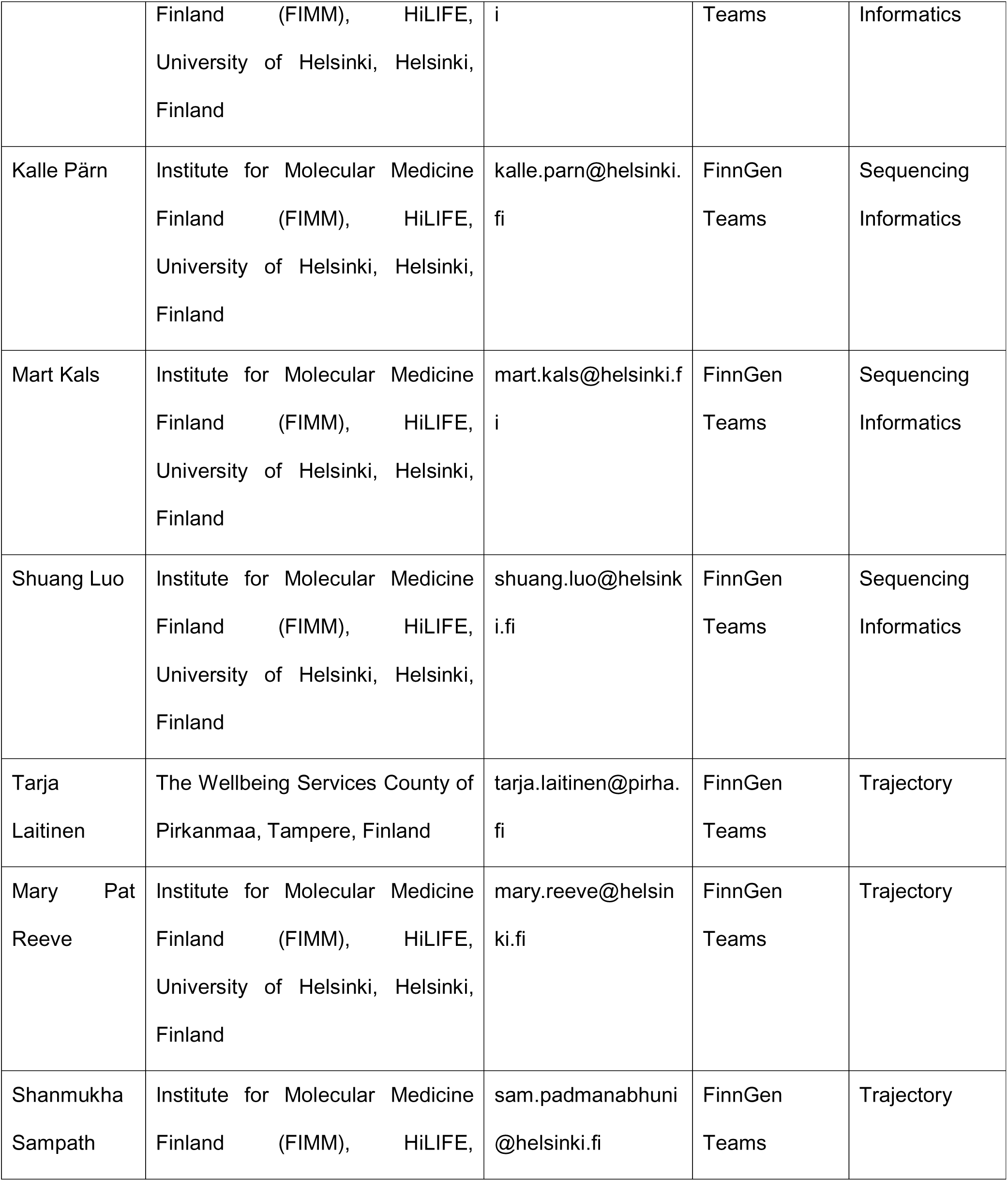
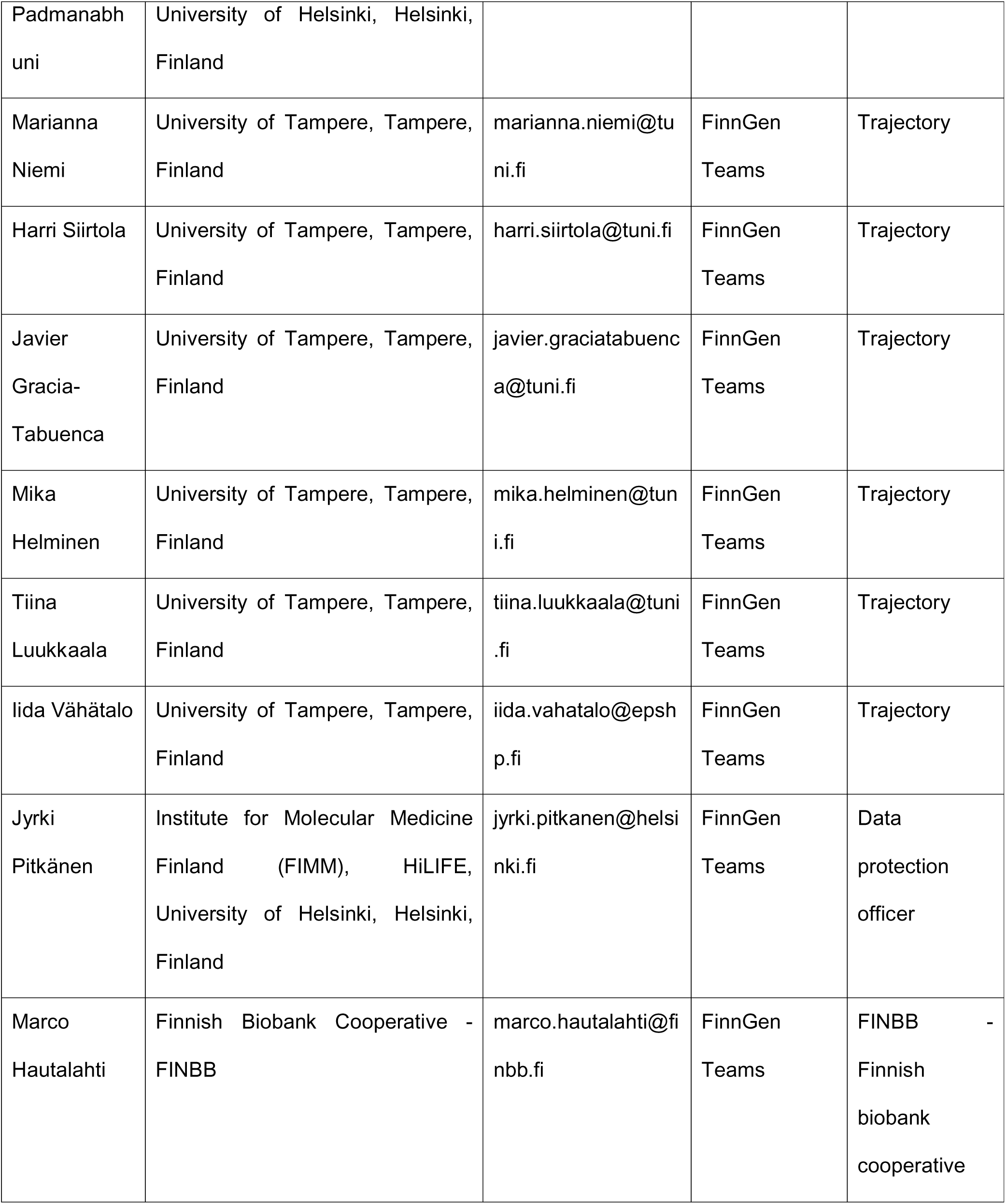
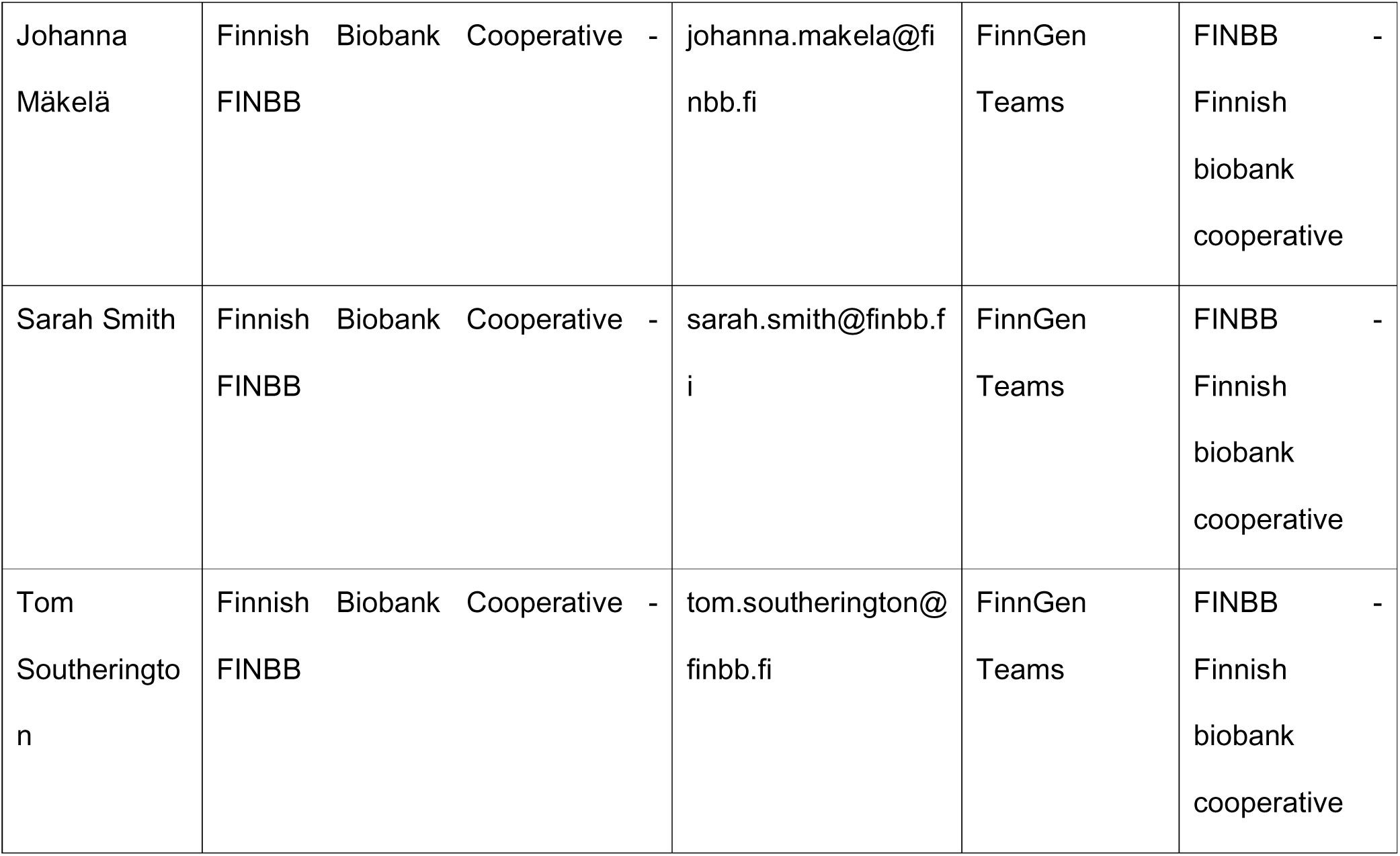

