## Supplementary Figure 1 for "TWINGEN – protocol for an observational clinical biobank recall and biomarker study to identify individuals with high risk of Alzheimer’s disease"

**Supplementary Figure 1. Blood samples collected in TWINGEN**

**5**

**6**

**2**

**4**

**3**

**1**

**RNA**

**SERUM**

**SERUM**

10 ml

10 ml

10 ml

2,5 ml

8,5 ml

8,5 ml

**30’ (max 60’) in RT	1500G 10’	30’ (max 60’) in RT	1500G 10’**

Stored as a whole blood -20 °C, final storage for RNA -80 °C

Twin Study research

**Pooled plasma	Pooled serum**

**EDTA-plasma aliquots x 16**

1 x 0,5 ml UEF for biomarker analysis

8 x 0,5 ml Twin Study research

7 x 0,5 ml THL Biobank

Stored first at -20 °C and final -80 °C

**SERUM aliquots x 14**

7 x 0,5 ml Twin Study research

7 x 0,5 ml THL Biobank

Stored first at -20 °C and final -80 °C

EDTA-plasma 2 ml mikroputki

16 kpl á 0,5 ml

Pakastus -20 °C

Huom! Kaikkia jakoputkia ei välttämättä saada.
